# The prognostic value of mean platelet volume in patients with coronary artery disease: a systematic review with meta-analyses

**DOI:** 10.1101/2024.03.20.24304646

**Authors:** Akhmetzhan Galimzhanov, Han Naung Tun, Yersin Sabitov, Francesco Perone, Tigen Mustafa Kursat, Erhan Tenekecioglu, Mamas A Mamas

**Affiliations:** Department of propedeutics of internal disease, Semey Medical University, Semey, Kazakhstan; Keele Cardiovascular Research Group, Keele University, Stoke on Trent, Keele, United Kingdom; Larner College of Medicine, University of Vermont, Burlington, Vermont, USA; Emergency Hospital, Semey, Kazakhstan; Cardiac Rehabilitation Unit, Rehabilitation Clinic “Villa delle Magnolie”, 81020 Castel Morrone, Caserta, Italy; Faculty of Medicine, Department of Cardiology, Marmara University, Istanbul, Türkiye; Department of Cardiology, Bursa Yuksek İhtisas Training and Research Hospital, Health Sciences University, Bursa, Türkiye; Department of Cardiology, Erasmus MC, Thorax Center, Erasmus University, Rotterdam, the Netherlands; Keele Cardiovascular Research Group, Keele University, Stoke on Trent, Keele, United Kingdom; National Institute for Health and Care Research (NIHR) Birmingham Biomedical Research Centre, UK

**Keywords:** mean platelet volume, coronary artery disease, prognosis, mortality, systematic review

## Abstract

**Background:** Mean platelet volume (MPV) is a widely available laboratory index, however its prognostic significance in patients with coronary artery disease (CAD) is still unclear. We intended to investigate and pool the evidence on the prognostic utility of admission MPV in predicting clinical outcomes in patients with CAD.

**Methods:** PubMed, Web of Science, and Scopus were the major databases used for literature search. The risk of bias was assessed using the quality in prognostic factor studies. We used random-effects pairwise analysis with the Knapp and Hartung approach supported further with permutation tests and prediction intervals (PIs).

**Results:** We identified 52 studies with 47066 patients. A meta-analysis of 9 studies with 14,864 patients demonstrated that 1 femtoliter increase in MPV values was associated with a rise of 29% in the risk of long-term mortality (hazard ratio (HR) 1.29, 95% confidence interval (CI) 1.22-1.37) in CAD as a whole. The results were further supported with PIs, permutation tests and leave-one-out sensitivity analyses. MPV also demonstrated its stable and significant prognostic utility in predicting long-term mortality as a linear variable in patients treated with percutaneous coronary intervention (PCI) and presented with acute coronary syndrome (ACS) (HR 1.29, 95% CI 1.20-1.39, and 1.29, 95% CI 1.19-1.39, respectively).

**Conclusion:** The meta-analysis found robust evidence on the link between admission MPV and the increased risk of long-term mortality in patients with CAD patients, as well as in patients who underwent PCI and patients presented with ACS.

**PROSPERO number:** CRD42023495287

## Introduction

Coronary artery disease (CAD) remains a significant contributor to cardiovascular mortality with the highest age-standardised rate 108.8 deaths per 100,000.^1^ In this regard, it is crucial to improve prediction of adverse clinical events in this population. While numerous studies have investigated potential new biomarkers in cardiovascular medicine, only a few have translated into clinical practice due to factors such as cost, logistical challenges, a lack of understanding of complex atherosclerotic pathophysiology, and patient population heterogeneity.^2^ Conversely, exploring the predictive value of readily available laboratory tests appears more feasible. For example, mean platelet volume (MPV) is a widely used, inexpensive parameter measured by routine automated analyzers.^3^ Higher MPV indicates larger, younger platelets with a greater prothrombotic potential.^4^ While several studies have been conducted to examine the prognostic value of MPV, there are inconsistent data around its utility in the prediction of ischemic outcomes.^5–9^ Previously, we conducted a systematic review focused around the prognostic utility of MPV but only in acute coronary syndrome patients,^10^ meanwhile another meta-analysis focused on only patients treated with percutaneous coronary intervention (PCI).^11^ With new data available, we decided to update the systematic review and meta-analysis and broaden the population of interest. We therefore set out to investigate and compile the data that was currently available regarding the prognostic usefulness of admission MPV in predicting clinical outcomes in patients with CAD as a whole, as well as in the subpopulation of patients who were treated with PCI, had ACS, or had stable CAD.

## Methods

The prospectively registered protocol for the meta-analysis is available in PROSPERO with the number <u>CRD42023495287</u>. The study followed the standards outlined in the Guide to systematic review and meta-analysis of prognostic factor studies and the Preferred Reporting Items for Systematic Reviews and MetaAnalyses (PRISMA) statement.^12,13^ PubMed, Web of Science, and Scopus were the major databases used for literature search. The search approach was developed utilising the following criteria: patient (coronary artery disease) - exposure (mean platelet volume) - outcome (mortality, major adverse cardiovascular events, etc.).^13^ The full search strategy is described in the Supplementary Materials.

We explored clinical trial registries, high-impact journal websites, conference proceedings to retrieve additional publications. The citation-based tracking was based on references from the included publications and prior meta-analyses, as well as the CoCites tool, which ranks articles based on their co-citation rates (Supplementary Table).^14^ We used a specific ShinyApp tool to create a flow-diagram of the systematic review.^15^

### Screening

The inclusion criteria were studies that investigated the prognostic importance of admission MPV in all-comer CAD patients in prediction of clinical endpoints, such as mortality and major adverse cardiovascular events. We excluded articles dedicated to patients younger than 18 years, pregnant women, terminal liver and kidney diseases, and life expectancy less than 3 years. In terms of study design, we omitted case-control and cross-sectional studies, case reports and series of cases, editorials, correspondence, brief reports, systematic and narrative reviews, and meta-analyses. The Rayyan web-based platform provided a machine-learning tool for semi-automated abstract selection to aid in the screening process.^16^

### Data extraction

The checklist for critical appraisal and data extraction for systematic reviews of prediction modeling studies—prognostic factors (CHARMS-PF) was applied to retrieve data from the original investigations.^13,17^ The extracted data included information on study characteristics (publication year, country, study design, follow-up period), study population (inclusion criteria, sample size, mean age, gender, prevalence of risk factors, provided laboratory parameters), and effect estimates. If the original studies did not provide overall mean and standard deviations for the parameter of interest, these statistics were calculated indirectly from the reported data with the use of specific formulae.^18,19^ A portion of the data was acquired directly from the primary investigation’s authors.

The risk of bias was assessed using the quality in prognostic factor studies (QUIPS) tool, which evaluates six major domains: participation, attrition, prognostic factor measurement, confounding bias, outcome measurement, and analysis and reporting.^20^ The screening, data extraction and the risk of bias assessment were conducted by several reviewers with any disagreement resolved via discussion with the whole team.

### Statistical analysis

We used random-effects pairwise analysis with both maximum (ML) and restricted maximum likelihood estimates (REML). To improve the interpretation of the results and reduce heterogeneity, statistical analyses were performed independently for studies with reported time-to-event and dichotomous effect estimates (hazard ratios (HRs) and odds ratios (ORs)), separately for studies that treated MPV as a linear or categorical variable, and separately for events that occurred at different times.^13^ To mitigate the confounding bias, we conducted all statistical analyses only with adjusted summary effect estimates. We opted for the Knapp and Hartung approach in our meta-analyses to provide more reliable estimates.^21–23^ While this method produces wider confidence intervals, it ensures a lower chance of falsely concluding an effect exists.^21^ In order to further decrease type I error rates, we carried out permutation tests to support statistically significant results of the meta-analyses.^24^ Prediction intervals (PIs) offer a valuable tool for meta-analysis, transcending the limitations of confidence intervals (CIs). While CIs focus on the precision of the pooled estimate, PIs capture the expected variability of true treatment effects across diverse settings, including those relevant to future patients encountered by clinicians.^25,26^ Therefore, we also calculated PIs for meta-analyses with more than 5 studies and statistically significant results.^26^

The heterogeneity of the analyses were calculated with chi-squared, I^2^, and tau-squared tests. To investigate how the prognostic significance of MPV varies depending on clinical scenarios, we performed subgroup analyses in different types of CAD. Furthermore, meta-regression analyses were carried out to investigate possible reasons for heterogeneity if the number of available studies was higher than 10. The publication bias was assessed both graphically (funnel plot asymmetry and trim and fill method) and statistically with Begg’s rank and Egger’s regression tests. The validity of results were further approved by leave-one-out sensitivity analyses. All statistical analyses were implemented with the *metafor* statistical R package.^27^ Given the potential influence of evidence certainty on interpretation and decision-making, the authors applied the GRADE approach and categorised the certainty of evidence as high, moderate, low, or very low.^28^

## Results

We identified 52 eligible studies with 47066 patients.^5–9,29–75^ The flow-diagram of the search is presented in Figure 1. The search in additional databases is described in detail in Supplementary Table 1. The list of excluded studies is provided in Supplementary Table 2. The research originated primarily in East Asia (16 studies), followed by the Middle East (15), Europe (12), Latin America (6), South Asia (2) and Australia (1). While a total of 26 studies had a prospective design, another 26 studies were retrospective in nature. The baseline characteristics of the original studies showed a notable degree of heterogeneity. The mean age varied from 36.4 to 78.5 years. The prevalence of male sex ranged from 57% to 82%. Risk factor prevalence varied widely, with smoking ranging from 22 to 58%, diabetes mellitus from 26 to 100%, dyslipidemia from 53 to 90%, and hypertension from 59 to 80%. The detailed overview around study characteristics are found in Supplementary Table 3.

**Figure 1.**
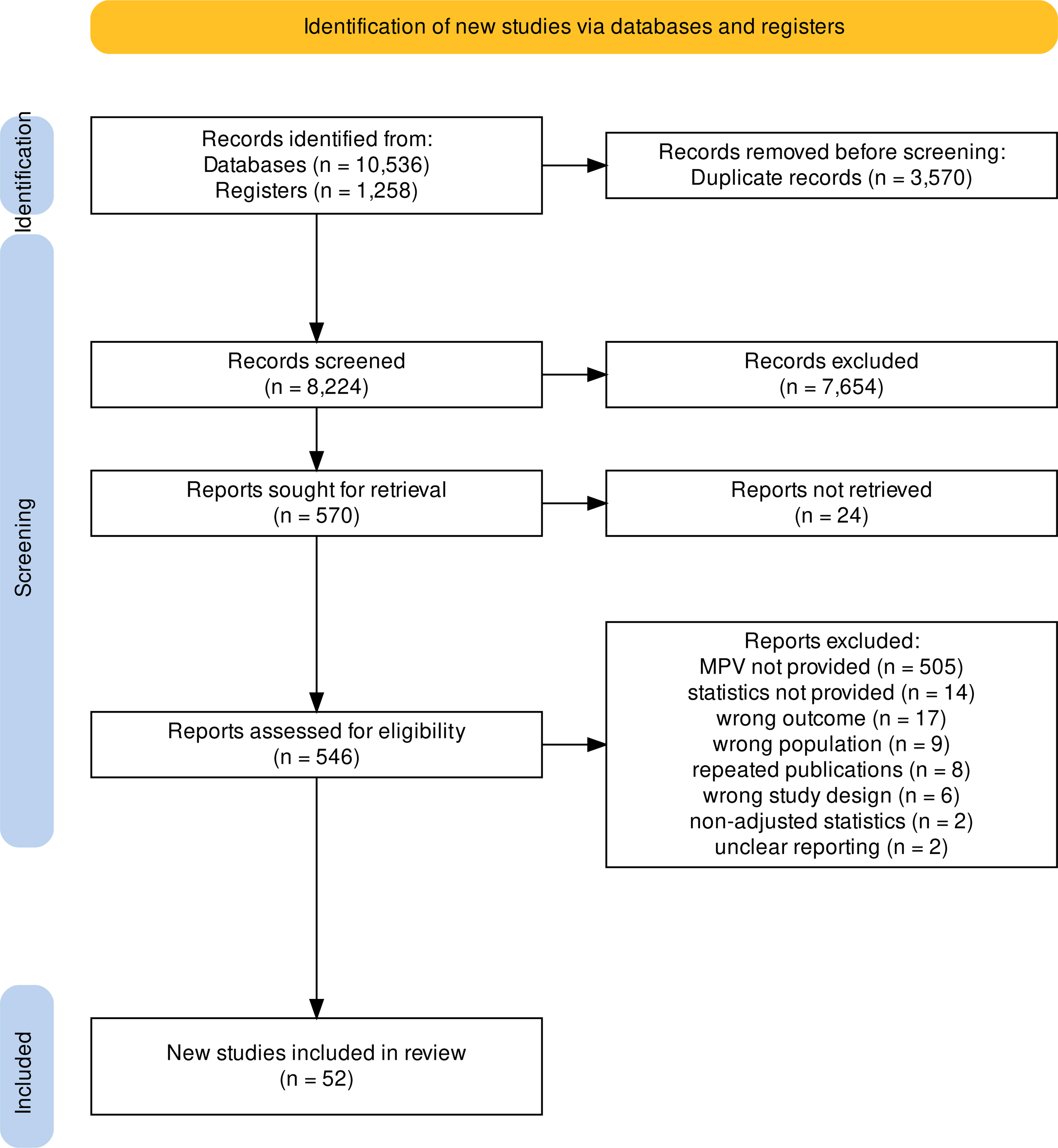
Flow diagram of the systematic search. MPV, mean platelet volume.

### The risk of bias assessment

Detailed information on the risk of bias assessment is presented in Table 1. Generally speaking, a total of 13 studies did not possess high risk of bias features in any of the 6 examined domains.^6,7,31,46,48,49,51,55,61,62,64,68,73^ Three research studies were deemed biased in four out of six categories,^50,65,72^ whereas eighteen papers were deemed high-risk in just one category.^5,8,9,29,30,33,35,38,40,41,45,52,53,56,59,60,67^ Four studies were classified as low-risk in four out of six domains,^5,6,46,62^ meanwhile a total of 18 studies only showed low-risk qualities in one domain.^31,32,34,36,41–43,49,50,52,59,63,66,69–72,75^ The review discovered significant flaws in some included studies. Concerns concerning bias arose from unclear participant selection processes, high dropout rates, and incomplete data reporting. Furthermore, potential measurement bias due to unreported analyzer details, uncontrolled confounding factors, and the possibility of selective reporting based on contradictions between methods and results were observed. Finally, because no studies were pre-registered. Determining the probability of selective reporting bias was challenging.

**Table 1.** The risk of bias assessment.

| **Study, year** | **Study Participati**<br>**on** | **Study Attritio**<br>**n** | **Prognostic**<br>**Factor**<br>**Measurement** | **Outcome**<br>**Measuremen**<br>**t** | **Study Confoundi**<br>**ng** | **Statistical**<br>**Analysis**<br>**and**<br>**Reporting** |
| --- | --- | --- | --- | --- | --- | --- |
| Osuna, 1999 | Low | High | Low | Moderate | Low | Moderate |
| Huzcek, 2005 | Low | Low | Moderate | Low | Low | High |
| Estevez-Loureir<br>o, 2009 | Low | High | Moderate | Low | Low | Moderate |
| Vakili, 2009 | High | Low | Moderate | Moderate | Low | Moderate |
| Akpek, 2011 | Moderate | Low | Moderate | Moderate | Moderate | Moderate |
| Goncalves,<br>2011 | Moderate | High | Moderate | Moderate | Low | High |
| Taglieri, 2011 | Low | Moderate | Low | Low | Low | Moderate |
| Tekbas, 2011 | Low | High | Moderate | Low | Low | Moderate |
| Dogan, 2012 | Low | High | Moderate | Moderate | High | Moderate |
| Lopez-Cuenca,<br>2012 | Moderate | High | Moderate | Moderate | Moderate | Moderate |
| Vrsalovic, 2012 | High | High | Moderate | Low | High | Moderate |
| Eisen, 2013 | Moderate | High | Low | Low | Low | Moderate |
| Akgul, 2013 | Low | High | Moderate | Low | High | High |
| Fabregat<br>Andreas, 2013 | Low | High | Low | Moderate | Low | Moderate |
| Nozari, 2013 | Moderate | Moderate | High | High | High | Moderate |
| Unal, 2013 | Low | Moderate | Moderate | High | Moderate | Moderate |
| Bergoli 2014 | High | Low | High | Moderate | High | Moderate |
| Lekston, 2014 | High | High | Moderate | Low | Moderate | High |
| Choi, 2014 | Moderate | High | Moderate | Moderate | High | Moderate |
| Liu, 2014 | Low | Low | Moderate | Moderate | High | Moderate |
| Seo, 2014 | Low | Low | Low | Low | Moderate | Moderate |
| Seyyed-Moham<br>madzad, 2014 | Low | Low | High | High | Moderate | Moderate |
| Siller-Matula,<br>2014 | Low | Moderate | Moderate | Low | Moderate | Moderate |
| Wan, 2014 | Low | Moderate | Moderate | Moderate | Moderate | Moderate |
| Ghaffari, 2015 | Low | High | High | Moderate | High | High |
| Lai, 2015 | Low | Low | Moderate | Low | Moderate | Moderate |
| Navarta, 2015 | Low | Moderate | Moderate | Moderate | High | Moderate |
| Ranjith, 2015 | Low | Low | Moderate | Moderate | Low | High |
| Lai, 2016 | High | Low | Low | Low | Moderate | Moderate |
| Wasilewski, 2016 | Moderate | Low | Low | Low | Moderate | Moderate |
| Tongtong Yu, 2017 | Low | High | Low | Moderate | Low | Moderate |
| Gina Yu, 2017 | Low | High | Low | Low | High | Moderate |
| Adam, 2018 | Low | High | Moderate | Low | High | Moderate |
| Machado, 2018 | Low | Moderate | Moderate | Moderate | High | Moderate |
| Monteiro Júnior, 2018 | Low | Low | Moderate | Low | High | Moderate |
| Tian, 2018 | Low | Moderate | Low | Moderate | Moderate | Moderate |
| Niu, 2018 | Low | Moderate | Low | Low | Low | Moderate |
| Wada, 2018 | Low | Low | Moderate | Low | Moderate | Moderate |
| Çanga, 2019 | Low | High | Moderate | Moderate | High | Moderate |
| Chang, 2019 | Low | Low | Moderate | Moderate | Low | Moderate |
| Garlobo, 2019 | High | High | High | High | Moderate | Moderate |
| Navarta, 2019 | Low | High | Moderate | Moderate | High | Moderate |
| Nozari, 2019 | High | Moderate | Moderate | Moderate | Moderate | Moderate |
| Şatiroğlu, 2019 | Low | Low | Moderate | Low | High | Moderate |
| Jiang, 2019 | Low | Moderate | Low | Moderate | Moderate | Moderate |
| Vogiatzis, 2019 | High | Low | High | Moderate | High | Moderate |
| Xinsen, 2020 | Low | High | High | Moderate | High | Moderate |
| Chen Xinsen, 2020 | Low | High | High | Moderate | Moderate | Moderate |
| Adali, 2022 | Low | High | High | High | Moderate | High |
| Liang, 2023 | Low | Low | Moderate | Low | Moderate | Moderate |
| Pedersen, 2023 | Low | Moderate | High | Low | Moderate | High |
| Toprak, 2023 | Low | High | High | Moderate | Moderate | Moderate |

The risk of bias assessment was performed according to the QUIPS tool. Hayden JA, van der Windt DA, Cartwright JL, Côté P, Bombardier C. Assessing bias in studies of prognostic factors. *Ann Intern Med.* 2013;158:280-286.

### The prognostic role of MPV in coronary artery disease

The results of the meta-analyses around the prognostic significance of MPV in all-comer CAD patients are presented in Figure 2 and Supplementary Figures S1-13. MPV as a linear variable was found to be an independent predictor of long-term mortality in CAD patients. A meta-analysis of 9 studies with 14,864 patients demonstrated that 1 femtoliter increase in MPV values was associated with a rise of 29% in the risk of follow-up mortality (HR 1.29, 95% CI 1.22-1.37). The results were further supported with calculating PIs (1.16-1.43), conducting permutation tests (1.29, 95% CI 1.21-1.39) and leave-one-out sensitivity analyses (Supplementary Table 4), and using REML as an estimator (1.29, 95% CI 1.22-1.37). The meta-analysis showed homogeneous results with the tau-squared statistics approximating 0. The publication bias was not detected with Begg’s rank and Egger’s regression tests being non-significant (p values 0.11 and 0.26, respectively, Supplementary Figure 1).

**Figure 2.**
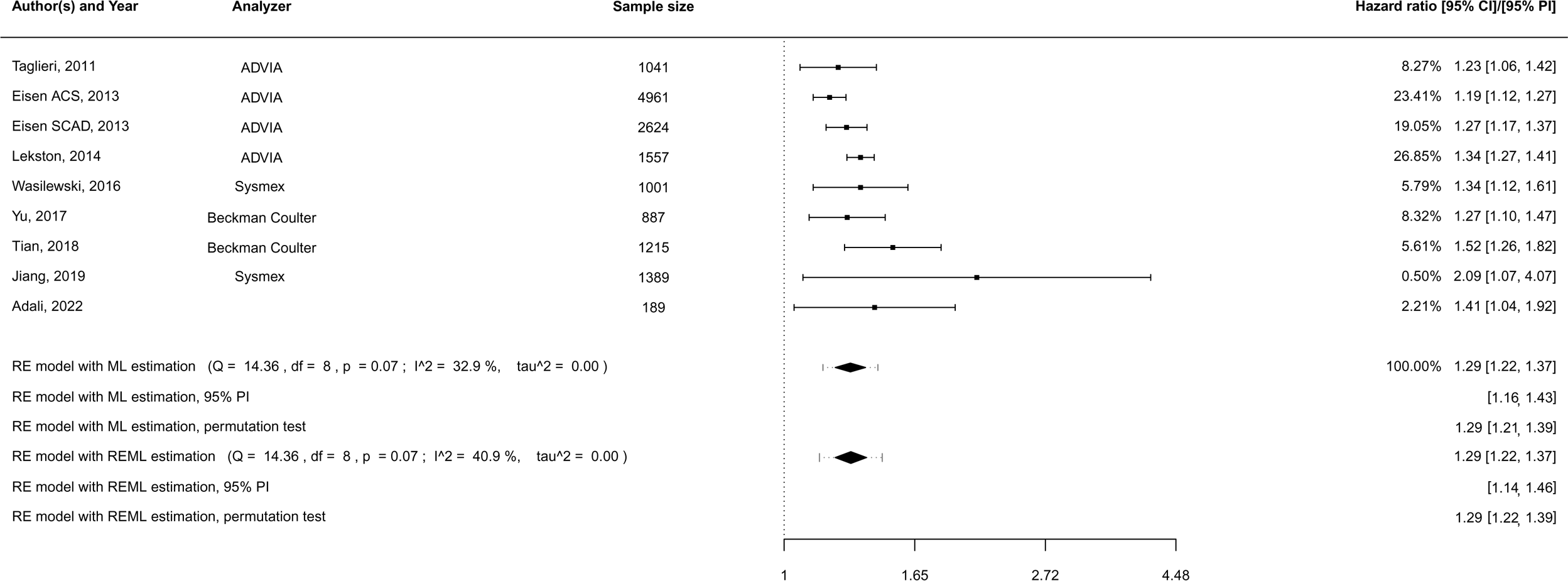
Meta-analysis for long-term mortality with MPV treated as a categorical variable and hazard ratios as effect estimates in patients with coronary artery disease. CI, confidence interval; ML, maximum likelihood; MPV, mean platelet volume; PI, prediction interval; RE, random effects; REML, restricted maximum likelihood.

The subgroup analysis did not detect any inconsistency of the results in the populations of stable CAD and ACS patients (P value for subgroup difference 0.98, Supplementary Table 5). The meta-regression analyses were non-informative due to the limited number of the included studies (Supplementary Table 6). The certainty of the evidence was graded as moderate.

The results for other outcomes were inconsistent. While some analyses showed a statistically significant connection, these results were not supported in the leave-one-out sensitivity analyses (MPV as a categorical variable for long-term mortality and analyses for in-hospital endpoints), calculation of prediction intervals (all in-hospital outcomes and long-term MACE), REML estimation (MPV as a categorical variable for long-term mortality), and publication bias assessment (for long-term and in-hospital MACE with MPV as a categorical variable, Supplementary Table 4 and Figures S1-13).

The subgroup analyses found statistically heterogeneous results in patients with ACS and stable CAD for long-term MACE with MPV being an independent predictor only in ACS subpopulation (p value for subgroup difference 0.03 and <0.001, Supplementary Table 5). Meanwhile, the meta-regression analysis also detected significant results of predictive importance of MPV in ACS patients compared to those in stable CAD patients in the analysis for long-term MACE. In addition, high heterogeneity in the meta-analyses could be explained with the prevalence of hypertension, smoking, and study design (Supplementary Table S6).

### The prognostic role of MPV in patients treated with PCI

Concerning patients who underwent PCI, MPV also demonstrated its stable and significant prognostic utility in predicting long-term mortality as a linear variable with HR 1.29 [95% CI 1.20-1.39] (7 studies with 13,634 participants). The analyses with PIs (1.14-1.46), permutation tests (HR 1.29, 95% CI 1.20-1.39), REML estimators (1.30, 95% CI 1.20-1.40) were also conclusive (Figure 3). The heterogeneity was low with the tau-squared near 0. After eliminating one study at a time, the leave-one-out sensitivity analysis did not show any significant variation (Supplementary Table 4). No publication bias was identified with graphical assessment (Supplementary Figure S14), with Begg’s rank (p value 0.38) and Egger’s regression tests (p value 0.09). The evidence was regarded as moderate. As far as other endpoints, the analyses were nonsignificant or inconclusive with permutation tests, PIs, REML estimators, leave-one-out sensitivity analyses (Supplementary Figures 14-19). While the subgroup analyses did not find any inconsistency of the results in patients with stable CAD and ACS for long-term endpoints and MPV as a linear variable, the difference was significant for long-term MACE and MPV as a categorical variable. The meta-regression analyses were not informative due to the limited number of studies.

**Figure 3.**
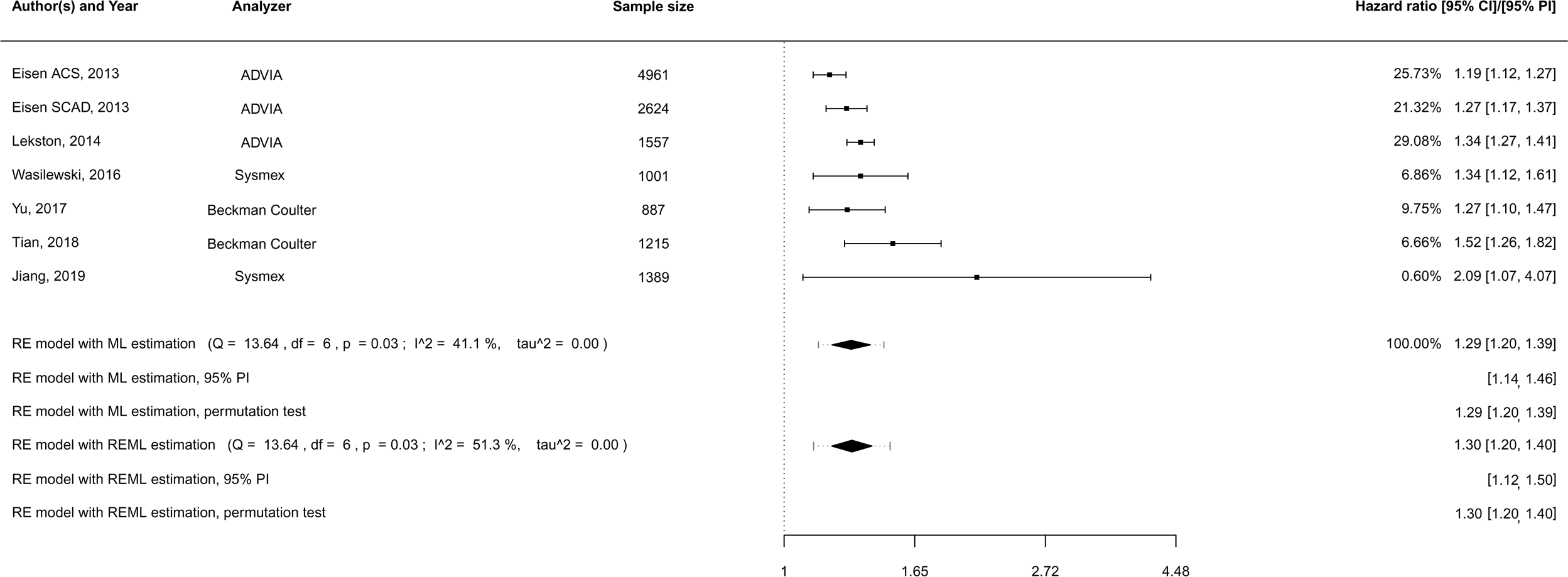
Meta-analysis for long-term mortality with MPV treated as a categorical variable and hazard ratios as effect estimates in patients treated with percutaneous coronary intervention. CI, confidence interval; ML, maximum likelihood; MPV, mean platelet volume; PI, prediction interval; RE, random effects; REML, restricted maximum likelihood.

### The prognostic role of MPV in ACS

The meta-analysis showed significant prognostic value of MPV as a continuous predictor for long-term mortality in ACS patients (HR 1.29, 95% CI 1.19-1.39) with the results being conclusive in leave-one-out sensitivity analyses (Supplementary Table 4), estimation with PIs (1.12-1.48), permutation tests (HR 1.29, 95% CI 1.19-1.39), and REML statistics (1.29, 95% 1.19, 1.40). The results were homogeneous (tau squared statistics approximating 0). Although the number of included studies were low, publication bias was not detected with both graphical and statistical methods (p value 0.71 and 0.48, Supplementary Figure S20-21). The evidence was graded as moderate.

With respect to the other endpoints, the results were either nonsignificant or inconsistent with leave-one-out sensitivity analyses (for inhospital outcomes), estimation of PIs (long-term MACE and all inhospital outcomes), REML statistics (long-term mortality with MPV as a categorical variable). The funnel plots were asymmetrical that suggests the possibility of publication bias in the analyses for long-term and inhospital MACE (all p values <0.001, Supplementary Figures, 22-32). The subgroup and meta-regression analyses were noninformative.

### The prognostic role of MPV in stable CAD

In stable CAD patients, MPV as a linear variable predicted the occurrence of long-term mortality with HR 1.29 (95% CI 1.07-1.55) with consistent results from PIs and REML estimators (Supplementary Figure S33-35). However, these findings became nonsignificant after eliminating one study at a time (Supplementary Table S4). The analyses for long-term MACE did not reveal a prognostic value of MPV, however all analyses for stable CAD patients were limited with a low number of included studies. Hence, we regarded the certainty of the evidence as low.

## Discussion

This systematic review and meta-analysis found robust evidence that MPV, as a linear variable, was linked to the increased risk of long-term mortality in an all-comer population of CAD patients, patients treated with PCI, and patients with ACS. We revealed that one femtoliter increase in admission MPV values was associated with a rise of 29% in the risk of follow-up death in patients with CAD overall, as well as in PCI and ACS subpopulations. It is known that around 25% of statistically significant results from traditional meta-analyses are deemed to be nonsignificant after calculation with Knapp and Hartung adjustment.^21^ However, since all of the CIs in our meta-analyses were determined using Knapp and Hartung adjustment and confirmed by permutation testing, the likelihood that these results are false-positive is relatively negligible. A prediction interval represents the degree of uncertainty we anticipate in the pooled estimates if a new study is added to the meta-analysis.^76^ As a result, it is highly unlikely that the inclusion of new research from the same population will affect the conclusions of our meta-analyses since all of the PIs demonstrated the predictive value of MPV in predicting long-term mortality.

Although there was some data on the prognostic utility of MPV in predicting MACE, these results were not supported with calculation of PIs, REML statistics, and conducting permutation tests. It could be partially explained with the disparities in used definitions for MACE. This emphasises the need of utilising internationally standard terminology for study endpoints to make it easier to compare and pool results in systematic reviews and meta-analyses.^77^ Moreover, we did not reveal conclusive findings regarding analyses with MPV treated as a categorical variable. Categorization of continuous exposures in epidemiological studies are customary, however, it is proven to lead to loss of statistical power, useful information, and efficiency.^78^ This could account for the discrepancy of results in analyses when MPV values were grouped by quantiles and tertiles.

There was a lack of studies investigating the nonlinear dose-response relationships between MPV and clinical outcomes in CAD patients. There is much data supporting the prognostic value of elevated MPV especially in acute thrombotic conditions. The high MPV values indicate the abundance of large-sized platelets known to be younger, more active with more prothrombotic potential.^4^ However, the evidence on the unfavourable role of low MPV values for prognosis is also accumulating. The study led by Wada et al enrolled a large cohort of patients with stable CAD treated with elective PCI and found that a low MPV, but not a high MPV, was associated with the increased risk of ischemic outcomes even after adjustment with traditional risk factors (HR 1.16 per 1 femtoliter decrease, 95% CI 1.04–1.30; lowest versus highest MPV groups HR 1.43, 95% CI 1.10–1.86).^7^ These findings highlight the possibility that distinct associations between MPV and clinical outcomes may result from differences in the pathophysiology of stable CAD and ACS. Our meta-regression and subgroup analyses also support this theory with a more significant prognostic value of MPV in ACS than in stable CAD. A study by Rief et al showed that low values of MPV were linked to the high risk of critical limb ischemia in peripheral artery disease patients (OR 0.84, 95% CI 0.75–0.94).^79^ Another study revealed a positive association between a low MPV and poor survival in pancreatic cancer individuals.^80^ Numerous studies found a negative correlation between MPV and autoimmune disease activity.^81–83^ In Behçet’s disease, for example, MPV decreased during disease exacerbation and rose during infection in same subjects.^81^ These results emphasise the close interaction between thrombosis and inflammation by pointing to a relationship between low MPV values and chronic inflammation. It is believed that an increase in big platelet consumption at the sites of inflammation is the cause of a drop in MPV.^3^ Large epidemiological studies are therefore desperately needed to examine the nonlinear relationship between MPV and clinical outcomes in patients with CAD across the entire MPV value range (Figure 4).

**Figure 4.**
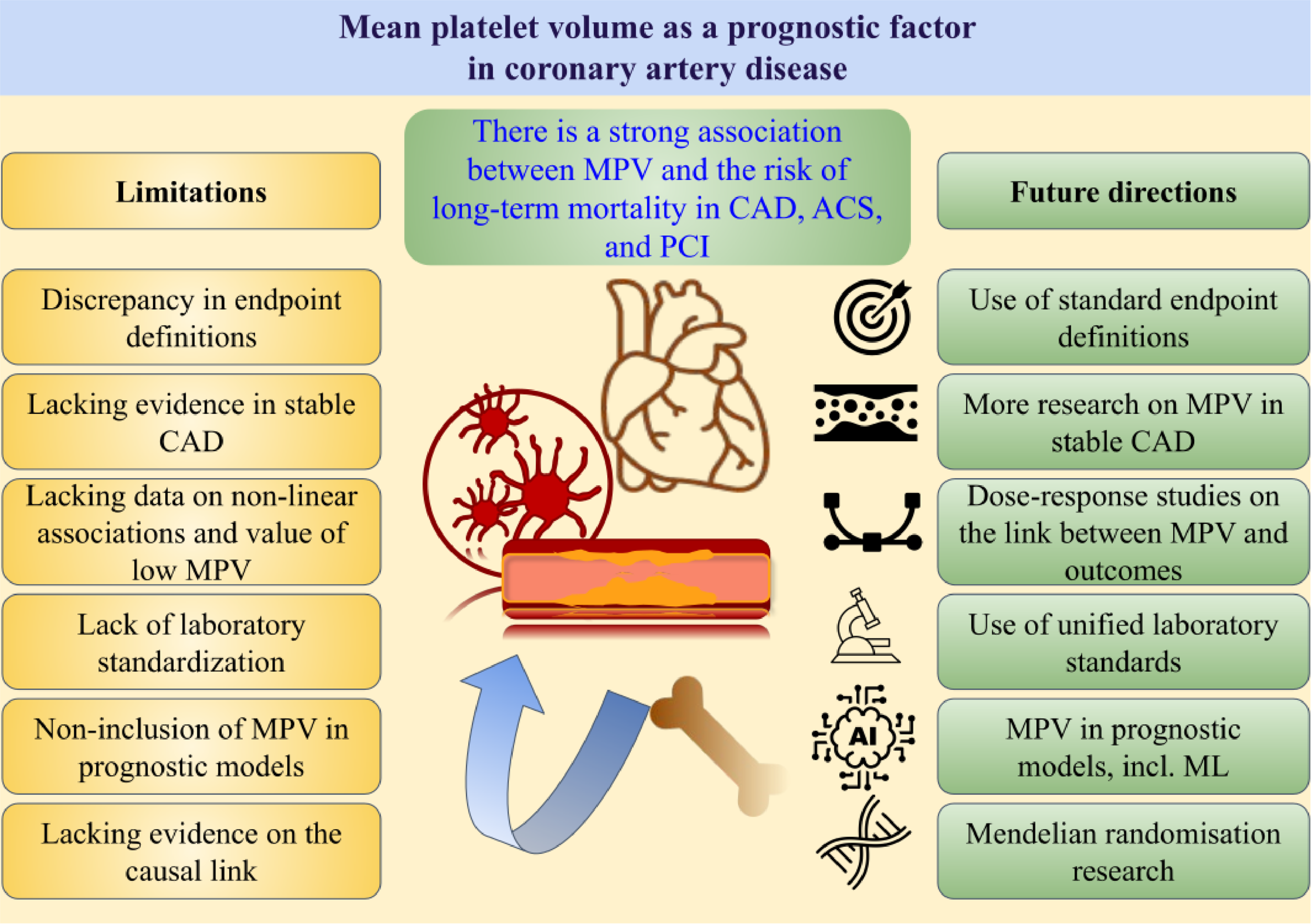
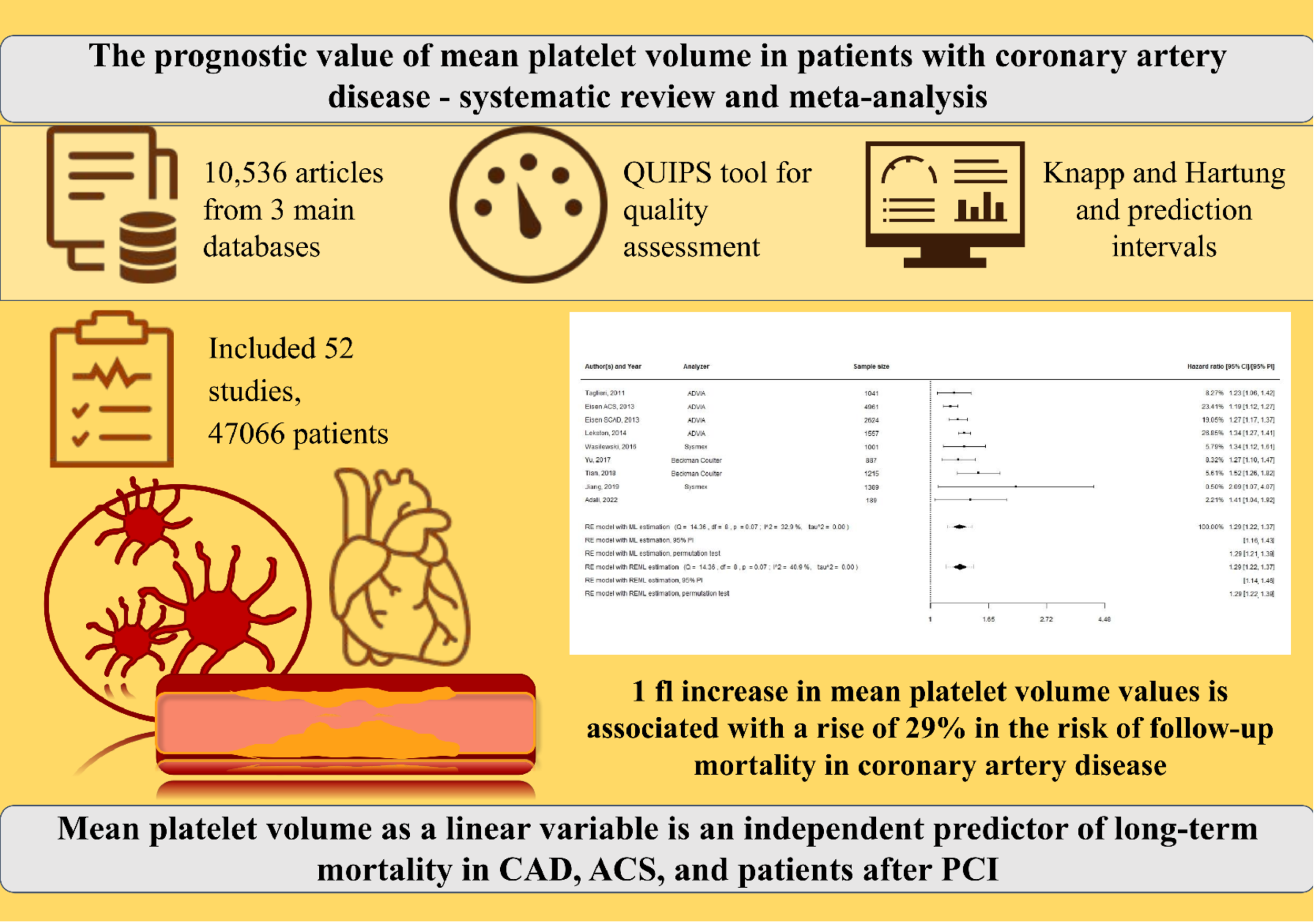
The limitations and future directions of research on mean platelet volume as a prognostic factor in coronary artery disease. ACS, acute coronary syndrome; CAD, coronary artery disease; ML, machine learning; MPV, mean platelet volume; PCI, percutaneous coronary intervention.

There is a lack of studies examining the additional values of MPV over prognostic scores in the prediction of adverse outcomes in patients with CAD. Taking into account the robust data on usefulness of MPV as a predictor and the availability of MPV in routine clinical practice, we believe that future studies should consider MPV as a candidate biomarker in decision-making models for CAD patients. A lack of laboratory standardisation was thought to hinder practical implementation of MPV as a biomarker in everyday clinical settings.^3,84^ Although we did not reveal any impact of vendors of automated analysers on the prognostic utility of MPV for long-term mortality in the meta-regression analyses, it is necessary to improve standardisation in the measurement of MPV. However, modern machine learning techniques can integrate multidimensional data, such as MPV values, analyzer vendor, in vitro anticoagulant used, time lag after sampling, flow cytometry method, and more. These techniques can create sophisticated prediction and decision-making models that are challenging to derive using traditional statistics. This is especially valid in the age of artificial intelligence (Figure 4).

Our systematic review has several limitations that are mainly due to limitations of the primary investigations. The retrospective design of half of the included studies points to the possibility of selection and recall bias in the review. Furthermore, a paucity of published data made it impossible to conduct meta-analyses for myocardial infarction, stent thrombosis, target vessel revascularization, stroke, heart failure, major bleeding. In addition, the observational nature of the primary studies justifies the only associative link between MPV and mortality in CAD patients. Despite extracting only adjusted statistics from the primary studies, the risk of confounding bias coil not be excluded. Future Mendelian randomisation studies would be of great interest to determine the causal relationship between genetically determined MPV and the risk of atherosclerosis.

## Conclusion

The current systematic review and meta-analysis found robust evidence on the link between admission MPV and the increased risk of long-term mortality in overall CAD patients, as well as in patients who underwent PCI and patients presented with ACS. Our meta-analysis showed that one femtoliter increase in MPV was associated with a rise of 29% in the risk of long-term mortality in the CAD population. Further studies are needed to investigate dose-response relationships and value of MPV in clinical decision making.

## Data Availability

This is a systematic review and meta-analysis based on the data from the primary studies

