## Supplementary Materials for "The prognostic value of mean platelet volume in patients with coronary artery disease: a systematic review with meta-analyses"

**Content**

Supplementary search strategy

Supplementary Table 1. Additional sources.

Supplementary Table 2. A list of some excluded studies with reasons for exclusion.

Supplementary Table 3. Baseline features of the included studies

Supplementary Table 4. Leave-one out sensitivity analyses.

Supplementary Table 5. Subgroup analyses.

Supplementary Table 6. Meta-regression analysis.

Supplementary Figures

- Supplementary Figure S1. All ischemic heart disease patients. Additional analyses for long-term mortality with MPV as a continuous variable and hazard ratios as effect estimates.
- Supplementary Figure S2. All ischemic heart disease patients. Analyses for long-term mortality with MPV treated as a categorical variable and hazard ratios as effect estimates.
- Supplementary Figure S3. All ischemic heart disease patients. Analyses for long-term major cardiovascular events with MPV treated as a continuous variable and hazard ratios as effect estimates.
- Supplementary Figure S4. All ischemic heart disease patients. Additional analyses for long-term major cardiovascular events with MPV treated as a continuous variable and hazard ratios as effect estimates.
- Supplementary Figure S5. All ischemic heart disease patients. Analyses for long-term major cardiovascular events with MPV treated as a categorical variable and hazard ratios as effect estimates.
- Supplementary Figure S6. All ischemic heart disease patients. Additional analyses for long-term major cardiovascular events with MPV treated as a categorical variable and hazard ratios as effect estimates.
- Supplementary Figure S7. All ischemic heart disease patients. Analyses for long-term major cardiovascular events with MPV treated as a categorical variable and odds ratios as effect estimates.
- Supplementary Figure S8. All ischemic heart disease patients. Analyses for long-term mortality with MPV treated as a categorical variable and odds ratios as effect estimates.
- Supplementary Figure S9. All ischemic heart disease patients. Analyses for one-month mortality with MPV treated as a continuous variable and hazard ratios as effect estimates.
- Supplementary Figure S10. All ischemic heart disease patients. Analyses for one-month mortality with MPV treated as a categorical variable and odds ratios as effect estimates.
- Supplementary Figure S11. All ischemic heart disease patients. Analyses for one-month major adverse cardiovascular events with MPV treated as a categorical variable and odds ratios as effect estimates.
- Supplementary Figure S12. All ischemic heart disease patients. Analyses for inhospital mortality with MPV treated as a categorical variable and odds ratios as effect estimates.
- Supplementary Figure S13. All ischemic heart disease patients. Analyses for inhospital major adverse cardiovascular events with MPV treated as a categorical variable and odds ratios as effect estimates.
- Supplementary Figure S14. Patients treated with percutaneous coronary intervention.

Additional analyses for long-term mortality with MPV treated as a continuous variable and hazard ratios as effect estimates.
Supplementary Figure S15. Patients treated with percutaneous coronary intervention. Analyses for long-term major adverse cardiovascular events with MPV treated as a continuous variable and hazard ratios as effect estimates.
Supplementary Figure S16. Patients treated with percutaneous coronary intervention. Analyses for long-term major adverse cardiovascular events with MPV treated as a categorical variable and hazard ratios as effect estimates.
Supplementary Figure S17. Patients treated with percutaneous coronary intervention. Analyses for long-term major adverse cardiovascular events with MPV treated as a categorical variable and odds ratios as effect estimates.
Supplementary Figure S18. Patients treated with percutaneous coronary intervention. Analyses for one-month mortality with MPV treated as a continuous variable and hazard ratios as effect estimates.
Supplementary Figure S19. Patients treated with percutaneous coronary intervention. Analyses for inhospital major adverse cardiovascular events with MPV treated as a categorical variable and odds ratios as effect estimates.
Supplementary Figure S20. Patients with acute coronary syndrome. Analyses for long-term mortality with MPV treated as a continuous variable and hazard ratios as effect estimates.
Supplementary Figure S21. Patients with acute coronary syndrome. Additional analyses for long-term mortality with MPV treated as a continuous variable and hazard ratios as effect estimates.
Supplementary Figure S22. Patients with acute coronary syndrome. Analyses for long-term mortality with MPV treated as a categorical variable and hazard ratios as effect estimates.
Supplementary Figure S23. Patients with acute coronary syndrome. Analyses for long-term major adverse cardiovascular events with MPV treated as a continuous variable and hazard ratios as effect estimates.
Supplementary Figure S24. Patients with acute coronary syndrome. Additional analyses for long-term major adverse cardiovascular events with MPV treated as a continuous variable and hazard ratios as effect estimates.
Supplementary Figure S25. Patients with acute coronary syndrome. Analyses for long-term major adverse cardiovascular events with MPV treated as a categorical variable and hazard ratios as effect estimates.
Supplementary Figure S26. Patients with acute coronary syndrome. Additional analyses for long-term major adverse cardiovascular events with MPV treated as a categorical variable and hazard ratios as effect estimates.
Supplementary Figure S27. Patients with acute coronary syndrome. Analyses for long-term major adverse cardiovascular events with MPV treated as a categorical variable and odds ratios as effect estimates.
Supplementary Figure S28. Patients with acute coronary syndrome. Analyses for one-month mortality with MPV treated as a continuous variable and hazard ratios as effect estimates.
Supplementary Figure S29. Patients with acute coronary syndrome. Analyses for one-month mortality with MPV treated as a categorical variable and odds ratios as effect estimates.
Supplementary Figure S30. Patients with acute coronary syndrome. Analyses for inhospital mortality with MPV treated as a categorical variable and odds ratios as effect estimates.
Supplementary Figure S31. Patients with acute coronary syndrome. Analyses for inhospital major adverse cardiovascular events with MPV treated as a categorical variable and odds ratios as effect estimates.

Supplementary Figure S32. Patients with acute coronary syndrome. Additional analyses for inhospital major adverse cardiovascular events with MPV treated as a categorical variable and odds ratios as effect estimates.

Supplementary Figure S33. Patients with stable coronary artery disease. Analyses for long-term mortality with MPV treated as a continuous variable and hazard ratios as effect estimates.

Supplementary Figure S34. Patients with stable coronary artery disease. Analyses for long-term major adverse cardiovascular events with MPV treated as a continuous variable and hazard ratios as effect estimates.

Supplementary Figure S35. Patients with stable coronary artery disease. Analyses for long-term major adverse cardiovascular events with MPV treated as a categorical variable and odds ratios as effect estimates.

**Supplementary search strategy**

**Database: PubMed**
**Last search date: 18 Dec 2023**
**Number of items: 5509**

#1 – Patient - (("Acute Coronary Syndrome"[MeSH Terms] OR "Coronary Artery Disease"[MeSH Terms] OR "Myocardial Infarction"[MeSH Terms] OR "Percutaneous Coronary Intervention"[MeSH Terms] OR "ST Elevation Myocardial Infarction"[MeSH Terms] OR "Non-ST Elevated Myocardial Infarction"[MeSH Terms] OR "angioplasty, balloon, coronary"[MeSH Terms] OR "Drug-Eluting Stents"[MeSH Terms] OR "Coronary Thrombosis"[MeSH Terms] OR "angina, unstable"[MeSH Terms] OR "angina pectoris, variant"[MeSH Terms] OR "Myocardial Ischemia"[MeSH Terms] OR "Coronary Care Units"[MeSH Terms] OR "Coronary Stenosis"[MeSH Terms] OR "acute coronary syndrom*"[Title/Abstract] OR "coronary artery diseas*"[Title/Abstract] OR "Myocardial Infarction"[Title/Abstract] OR "ischemic heart diseas*"[Title/Abstract] OR "ischaemic heart diseas*"[Title/Abstract] OR "percutaneous coronary intervention*"[Title/Abstract] OR "primary angioplasty"[Title/Abstract] OR "Drug-Eluting Stents"[Title/Abstract] OR "Drug-Eluted Stents"[Title/Abstract] OR "STEMI"[Title/Abstract] OR "unstable angina"[Title/Abstract] OR "bare metal stent*"[Title/Abstract] OR "myocardial infarction with st"[Title/Abstract] OR "myocardial infarction without st"[Title/Abstract] OR "coronary angioplast*"[Title/Abstract] OR "coronary stent*"[Title/Abstract])

#2 – Exposure - ("Mean Platelet Volume"[MeSH Terms] OR "Blood Platelets"[MeSH Terms] OR "Platelet Count"[MeSH Terms] OR "Blood Cell Count"[MeSH Terms] OR "Thrombocytopenia"[MeSH Terms] OR "Thrombocytosis"[MeSH Terms] OR "Mean Platelet Volume"[Title/Abstract] OR "blood platelet*"[Title/Abstract] OR "platelet count*"[Title/Abstract] OR "platelet volum*"[Title/Abstract] OR "thrombocytopen*"[Title/Abstract] OR "Thrombocytosis"[Title/Abstract] OR "platelet indic*"[Title/Abstract] OR "platelet parameter*"[Title/Abstract] OR "platelet index*"[Title/Abstract] OR "large platelet*"[Title/Abstract] OR "larger platelet*"[Title/Abstract] OR "platelet size"[Title/Abstract] OR "complete blood count*"[Title/Abstract] OR "hematological parameter*"[Title/Abstract] OR "haematological parameter*"[Title/Abstract] OR "hematological marker*"[Title/Abstract] OR "haematological marker*"[Title/Abstract] OR "platelet marker*"[Title/Abstract] OR "hematological indic*"[Title/Abstract] OR "haematological indic*"[Title/Abstract] OR "hematological index*"[Title/Abstract] OR "haematological index*"[Title/Abstract] OR "platelet number*"[Title/Abstract]))

#3 – Outcome - ("Mortality"[MeSH Terms] OR "Hospital Mortality"[MeSH Terms] OR "Prognosis"[MeSH Terms] OR "Biomarkers"[MeSH Terms] OR "Follow-Up Studies"[MeSH Terms] OR "Kaplan-Meier Estimate"[MeSH Terms] OR "Proportional Hazards Models"[MeSH Terms] OR "Odds Ratio"[MeSH Terms] OR "Predictive Value of Tests"[MeSH Terms] OR "Prospective Studies"[MeSH Terms] OR "Cohort Studies"[MeSH Terms] OR "Risk Assessment"[MeSH Terms] OR "Risk Factors"[MeSH Terms] OR "Heart Disease Risk Factors"[MeSH Terms] OR "Cardiometabolic Risk Factors"[MeSH Terms] OR "ROC Curve"[MeSH Terms] OR "Clinical Decision Rules"[MeSH Terms] OR "Survival Analysis"[MeSH Terms] OR "Heart Arrest"[MeSH Terms] OR "Area Under Curve"[MeSH Terms] OR "Cause of Death"[MeSH Terms] OR "coronary thrombosis/prevention and control"[MeSH Terms] OR "Survival Rate"[MeSH Terms] OR "Coronary Restenosis"[MeSH Terms] OR "long-term prognos*"[Title/Abstract] OR "long-term outcom*"[Title/Abstract] OR "long-term surviv*"[Title/Abstract] OR "long-term adverse outcom*"[Title/Abstract] OR "long term prognos*"[Title/Abstract] OR "long term outcom*"[Title/Abstract] OR "long term adverse outcom*"[Title/Abstract] OR "long term surviv*"[Title/Abstract] OR "short-term prognos*"[Title/Abstract] OR "short-term outcom*"[Title/Abstract] OR "short-term adverse outcom*"[Title/Abstract] OR "short-term surviv*"[Title/Abstract] OR "short term prognos*"[Title/Abstract] OR "short term outcom*"[Title/Abstract] OR "short term adverse outcom*"[Title/Abstract] OR "short term surviv*"[Title/Abstract] OR "cardiovascular outcom*"[Title/Abstract] OR "cardio-vascular outcom*"[Title/Abstract] OR "cardiac outcom*"[Title/Abstract] OR "cerebral outcom*"[Title/Abstract] OR "cerebrovascular outcom*"[Title/Abstract] OR "thrombotic outcom*"[Title/Abstract] OR "ischemic outcom*"[Title/Abstract] OR "ischaemic outcom*"[Title/Abstract] OR "cardiovascular endpoint*"[Title/Abstract] OR "cardiac endpoint*"[Title/Abstract] OR "cerebrovascular endpoint*"[Title/Abstract] OR "thrombotic endpoint*"[Title/Abstract] OR "ischaemic endpoint*"[Title/Abstract] OR "ischemic endpoint*"[Title/Abstract] OR "cardiovascular end point*"[Title/Abstract] OR "cardiac end point*"[Title/Abstract] OR "cerebrovascular endpoint*"[Title/Abstract] OR "cerebrovascular end point*"[Title/Abstract] OR "ischaemic end point*"[Title/Abstract] OR "ischemic end point*"[Title/Abstract] OR "cardiovascular end-point*"[Title/Abstract] OR "cardiac end-point*"[Title/Abstract] OR "cerebrovascular end-point*"[Title/Abstract] OR "ischaemic end-point*"[Title/Abstract] OR "ischemic end-point*"[Title/Abstract] OR "cardiovascular complication*"[Title/Abstract] OR "cardio-vascular complication*"[Title/Abstract] OR "cardiac complication*"[Title/Abstract] OR "cerebral complication*"[Title/Abstract] OR "cerebrovascular complication*"[Title/Abstract] OR "cerebro-vascular complication*"[Title/Abstract] OR "thrombotic complication*"[Title/Abstract] OR "ischaemic complication*"[Title/Abstract] OR "ischemic complication*"[Title/Abstract] OR "adverse cardiac event*"[Title/Abstract] OR "adverse cardiovascular event*"[Title/Abstract] OR "adverse cardio-vascular event*"[Title/Abstract] OR "major adverse cardiovascular event*"[Title/Abstract] OR MACCE[Title/Abstract] OR "cerebral event"[Title/Abstract] OR "cerebrovascular event"[Title/Abstract] OR "reinfarction"[Title/Abstract] OR "re-infarction"[Title/Abstract] OR "Coronary Restenosis"[Title/Abstract] OR "coronary re-stenosis"[Title/Abstract] OR "stent thrombosis"[Title/Abstract] OR "area under curve*"[Title/Abstract] OR "survival analys*"[Title/Abstract] OR "target vessel revascularization"[Title/Abstract] OR "target vessel revascularisation"[Title/Abstract] OR "target lesion revascularization"[Title/Abstract] OR "target lesion revascularisation"[Title/Abstract] OR "kaplan meir curve*"[Title/Abstract] OR "kaplan meir analys*"[Title/Abstract] OR "kaplan meir surviv*"[Title/Abstract] OR "proportional hazard model*"[Title/Abstract] OR "roc curve*"[Title/Abstract] OR "cardiovascular death"[Title/Abstract] OR "cardio-vascular death"[Title/Abstract] OR "cardiac death"[Title/Abstract] OR "cardiovascular risk"[Title/Abstract] OR "cardio-vascular risk"[Title/Abstract] OR "cerebrovascular risk"[Title/Abstract] OR "cerebro-vascular risk"[Title/Abstract] OR "atherothrombotic risk"[Title/Abstract] OR "thrombotic risk"[Title/Abstract] OR "ischemic risk"[Title/Abstract] OR "ischaemic risk"[Title/Abstract] OR "Mortality"[Title/Abstract])

#4 - #1 AND #2 AND #3

**Database: Web of Science**
**Last search date: 18 Dec 2023**
**Number of items: 1670**

### 1 - TOPIC: ((("myocardial infarct*" OR (myocardial NEAR/0 infarction) ) NEAR/2 "st segment*"))
Databases= WOS, DIIDW, KJD, MEDLINE, RSCI, SCIELO Timespan=All years
Search language=Auto

### 2 - TOPIC: (("myocardial infarct*" OR (myocardial NEAR/0 infarction) ) NEAR/2 eleva*)
Databases= WOS, DIIDW, KJD, MEDLINE, RSCI, SCIELO Timespan=All years
Search language=Auto

### 3 - TOPIC: (STEMI)
Databases= WOS, DIIDW, KJD, MEDLINE, RSCI, SCIELO Timespan=All years
Search language=Auto

### 4 - TOPIC: ("myocardial infarct*" OR (myocardial NEAR/0 infarction) )
Databases= WOS, DIIDW, KJD, MEDLINE, RSCI, SCIELO Timespan=All years
Search language=Auto

### 5 - TOPIC: ("acute coronar* syndrom*" OR (acute NEAR/1 coronary NEAR/0 syndrome) )
Databases= WOS, DIIDW, KJD, MEDLINE, RSCI, SCIELO Timespan=All years
Search language=Auto

### 6 - TOPIC: ("coronar* arter* diseas*" OR (coronary NEAR/0 artery NEAR/0 disease) )
Databases= WOS, DIIDW, KJD, MEDLINE, RSCI, SCIELO Timespan=All years
Search language=Auto

### 7 - TOPIC: ("isch$emic* heart diseas*" OR (ischemic NEAR/0 heart NEAR/0 disease) )
Databases= WOS, DIIDW, KJD, MEDLINE, RSCI, SCIELO Timespan=All years
Search language=Auto

### 8 - TOPIC: ("percutan* coronar* intervention*" OR (percutaneous NEAR/0 coronary NEAR/0 intervention) )
Databases= WOS, DIIDW, KJD, MEDLINE, RSCI, SCIELO Timespan=All years
Search language=Auto

### 9 - TOPIC: ((Drug-Eluting NEAR/0 Stent) OR "drug-elut* stent*") OR TOPIC: ((*metal* NEAR/0 stent*) OR "bare-metal stent*")
Databases= WOS, DIIDW, KJD, MEDLINE, RSCI, SCIELO Timespan=All years
Search language=Auto

### 10 - TOPIC: ((angioplast* NEAR/3 coronar*) OR (coronary NEAR/3 angioplasty) )
Databases= WOS, DIIDW, KJD, MEDLINE, RSCI, SCIELO Timespan=All years
Search language=Auto

### 11 - TOPIC: ((stent* NEAR/3 coronar*) OR (coronary NEAR/3 stenting) )
Databases= WOS, DIIDW, KJD, MEDLINE, RSCI, SCIELO Timespan=All years
Search language=Auto

### 12 - TOPIC: ("unstable angina" OR (unstable NEAR/0 angina) )
Databases= WOS, DIIDW, KJD, MEDLINE, RSCI, SCIELO Timespan=All years
Search language=Auto

# 13 - #12 OR #11 OR #10 OR #9 OR #8 OR #7 OR #6 OR #5 OR #4 OR #3 OR #2 OR #1
Databases= WOS, DIIDW, KJD, MEDLINE, RSCI, SCIELO Timespan=All years
Search language=Auto

### 14 - TOPIC: (((platelet* OR thrombocyt*) OR (platelet OR thrombocyte) ) NEAR/2 ((volume OR size OR count* OR number* OR indic* OR parameter* OR index* OR marker*) OR (volume OR size OR count OR number OR indice OR parameter OR index OR marker) ))
Databases= WOS, DIIDW, KJD, MEDLINE, RSCI, SCIELO Timespan=All years
Search language=Auto

### 15 - TOPIC: (((h$ematol* OR h$emogram*) OR (hematology OR hemogram OR hematological) ) NEAR/2 ((indic* OR parameter* OR index* OR marker*) OR (indice OR parameter OR index OR marker) ))
Databases= WOS, DIIDW, KJD, MEDLINE, RSCI, SCIELO Timespan=All years
Search language=Auto

### 16 - TOPIC: ((blood NEAR/2 platelet) OR (blood NEAR/2 platelet*) )
Databases= WOS, DIIDW, KJD, MEDLINE, RSCI, SCIELO Timespan=All years
Search language=Auto

### 17 - TOPIC: ("mean platelet volume" OR (mean NEAR/0 platelet NEAR/0 volume) )
Databases= WOS, DIIDW, KJD, MEDLINE, RSCI, SCIELO Timespan=All years
Search language=Auto

### 18 - TOPIC: ((large NEAR/1 platelet) OR (larg* NEAR/1 platelet*) )
Databases= WOS, DIIDW, KJD, MEDLINE, RSCI, SCIELO Timespan=All years
Search language=Auto

### 19 - TOPIC: ((complet* blood count*) OR (complete NEAR/1 blood NEAR/1 count) )
Databases= WOS, DIIDW, KJD, MEDLINE, RSCI, SCIELO Timespan=All years
Search language=Auto

### 20 - TOPIC: (thrombocytop* OR thrombocytopenia OR thrombocytosis)
Databases= WOS, DIIDW, KJD, MEDLINE, RSCI, SCIELO Timespan=All years
Search language=Auto

# 21 - #20 OR #19 OR #18 OR #17 OR #16 OR #15 OR #14
Databases= WOS, DIIDW, KJD, MEDLINE, RSCI, SCIELO Timespan=All years
Search language=Auto

### 22 - TOPIC: ((long-term OR short-term ) NEAR/1 ((prognos* OR outcom* OR surviv*) OR (prognosis OR outcome OR survival) ))
Databases= WOS, DIIDW, KJD, MEDLINE, RSCI, SCIELO Timespan=All years
Search language=Auto

### 23 - TOPIC: ((cardiovascular OR cardio-vascular OR cardiac OR cerebral OR cerebrovascular OR cerebro-vascular OR atherothromb* OR atherothrombotic OR athero-thromb* OR thromb* OR thrombotic OR isch$emic OR ischemic) NEAR/1 (outcom* OR outcome OR endpoint* OR endpoint OR end-pont* OR end-pont OR complicat* OR complication) )
Databases= WOS, DIIDW, KJD, MEDLINE, RSCI, SCIELO Timespan=All years
Search language=Auto

### 24 - TOPIC: (adverse NEAR/1 (cardi* OR cardiac OR cardiovascular OR cardio-vascular) NEAR/1 (cerebr* OR cerebral OR cerebrovascular OR cerebro-vascular) NEAR/1 (event* OR event OR outcome OR outcom* OR endpoint* OR endpoint OR end-pont* OR end-point) )
Databases= WOS, DIIDW, KJD, MEDLINE, RSCI, SCIELO Timespan=All years
Search language=Auto

### 25 - TOPIC: (adverse NEAR/1 (cardi* OR cardiac OR cardiovascular OR cardio-vascular) NEAR/1 (event* OR event OR outcome OR outcom* OR endpoint* OR endpoint OR end-pont* OR end-point) )

Databases= WOS, DIIDW, KJD, MEDLINE, RSCI, SCIELO Timespan=All years
Search language=Auto

### 26 - TOPIC: (MACCE)
Databases= WOS, DIIDW, KJD, MEDLINE, RSCI, SCIELO Timespan=All years
Search language=Auto

### 27 - TOPIC: ((cardiovascular OR cardio-vascular OR cardiac OR cerebral OR cerebrovascular OR cerebro-vascular OR atherothromb* OR atherothrombotic OR athero-thromb* OR thromb* OR thrombotic OR isch$emic OR ischemic) NEAR/2 risk)
Databases= WOS, DIIDW, KJD, MEDLINE, RSCI, SCIELO Timespan=All years
Search language=Auto

### 28 - TOPIC: (mortality)
Databases= WOS, DIIDW, KJD, MEDLINE, RSCI, SCIELO Timespan=All years
Search language=Auto

### 29 - TOPIC: ((cardiovascular OR cardio-vascular OR cardiac OR cerebral OR cerebrovascular OR cerebro-vascular OR atherothromb* OR atherothrombotic OR athero-thromb* OR thromb* OR thrombotic OR isch$emic OR ischemic) NEAR/2 death)
Databases= WOS, DIIDW, KJD, MEDLINE, RSCI, SCIELO Timespan=All years
Search language=Auto

### 30 - TOPIC: ((proport* OR proportional) NEAR/1 (hazard* OR hazard) NEAR/1 (model* OR model OR analys$s OR analysis) )
Databases= WOS, DIIDW, KJD, MEDLINE, RSCI, SCIELO Timespan=All years
Search language=Auto

### 31 - TOPIC: (reinfarction OR re-infarction)
Databases= WOS, DIIDW, KJD, MEDLINE, RSCI, SCIELO Timespan=All years
Search language=Auto

### 32 - TOPIC: ((coronar* OR coronary) NEAR/1 (restenos* OR re-stenos* OR restenosis OR re-stenosis) )
Databases= WOS, DIIDW, KJD, MEDLINE, RSCI, SCIELO Timespan=All years
Search language=Auto

### 33 - TOPIC: ((stent* OR stent) NEAR/0 (thromb* OR thrombosis) )
Databases= WOS, DIIDW, KJD, MEDLINE, RSCI, SCIELO Timespan=All years
Search language=Auto

### 34 - TOPIC: (kaplan-meir NEAR/2 (curve* OR analys$s OR curve OR analysis) )
Databases= WOS, DIIDW, KJD, MEDLINE, RSCI, SCIELO Timespan=All years
Search language=Auto

### 35 - TOPIC: ((surviv* OR survival) NEAR/0 (analys$s OR rate* OR analysis OR rate) )
Databases= WOS, DIIDW, KJD, MEDLINE, RSCI, SCIELO Timespan=All years
Search language=Auto

### 36 - TOPIC: ("roc curve*" OR (roc near/0 curve) )
Databases= WOS, DIIDW, KJD, MEDLINE, RSCI, SCIELO Timespan=All years
Search language=Auto

### 37 - TOPIC:  ("area under curve*" OR (area NEAR/0 under NEAR/0 curve) )
Databases= WOS, DIIDW, KJD, MEDLINE, RSCI, SCIELO Timespan=All years
Search language=Auto

### 38 - TOPIC:  ((revasculari$ation OR re-vasculari$ation OR revascularization OR re-vascularization)  NEAR  (("target vessel*" OR "target lesion*")  OR  (target NEAR (vessel OR lesion)  )))
Databases= WOS, DIIDW, KJD, MEDLINE, RSCI, SCIELO Timespan=All years
Search language=Auto

# 39 - #38  OR  #37  OR  #36  OR  #35  OR  #34  OR  #33  OR  #32  OR  #31  OR  #30  OR  #29  OR  #28  OR  #27  OR  #26  OR  #25  OR  #24  OR  #23  OR  #22
Databases= WOS, DIIDW, KJD, MEDLINE, RSCI, SCIELO Timespan=All years
Search language=Auto

### 40 - #39  AND  #21  AND  #13
Databases= WOS, DIIDW, KJD, MEDLINE, RSCI, SCIELO Timespan=All years
Search language=Auto

**Database: Scopus**
**Last search date: 18 Dec 2023**
**Number of items: 3357**

#1 – Patient: (( TITLE-ABS-KEY ( "myocardial infarction" W/2 "st segment" ) ) OR ( TITLE-ABS-KEY ( "myocardial infarction" W/2 eleva* ) ) OR ( TITLE-ABS-KEY ( stemi ) ) OR ( TITLE-ABS-KEY ( "myocardial infarction" ) ) OR ( TITLE-ABS-KEY ( "acute coronary syndrome" ) ) OR ( TITLE-ABS-KEY ( "coronary artery disease" ) ) OR ( TITLE-ABS-KEY ( "ischemic heart disease" ) ) OR ( TITLE-ABS-KEY ("ischaemic heart disease" ) ) OR ( TITLE-ABS-KEY ( "Percutaneous Coronary Intervention" ) ) OR ( TITLE-ABS-KEY ( "Drug-Eluting Stents" ) ) OR ( TITLE-ABS-KEY ( "metal stent" ) ) OR ( TITLE-ABS-KEY ( angioplasty W/3 coronary ) ) OR ( TITLE-ABS-KEY ( stent W/3 coronary ) ) OR ( TITLE-ABS-KEY ( "unstable angina" ) ))

#2 – Exposure: (( TITLE-ABS-KEY ( ( platelet OR thrombocyte ) W/2 ( volume OR size OR count OR number OR indice OR parameter OR index OR marker ) ) ) OR ( TITLE-ABS-KEY ( "mean platelet volume" ) ) OR ( TITLE-ABS-KEY ( "blood platelet" ) ) OR ( TITLE-ABS-KEY ( ( hematological OR hemogram OR hematology OR haematological OR haemogram ) W/1 ( indice OR parameter OR index OR marker ) ) ) OR ( TITLE-ABS-KEY ( "large platelet" ) ) OR ( TITLE-ABS-KEY ( "complete blood count" ) ) OR ( TITLE-ABS-KEY ( complete W/1 blood W/1 count ) ) OR ( TITLE-ABS-KEY ( thrombocytopenia OR thrombocytosis OR thrombocytop*) ))

#3 – Outcome: ((TITLE-ABS-KEY ((long-term OR short-term ) PRE/1 ( prognosis OR outcome OR survival))) OR (TITLE-ABS-KEY ((cardiovascular OR cardio-vascular OR cardiac OR cerebral OR cerebrovascular OR cerebro-vascular OR atherothrombotic OR athero-thrombotic OR atherothromb* OR athero-thromb* OR thrombotic OR thrombo* OR ischemic OR ischaemic) PRE/1 (outcome OR endpoint OR complication OR end-point OR "end point"))) OR (TITLE-ABS-KEY (adverse PRE/1 (cardiac OR cardiovascular OR cardio-vascular) PRE/1 (cerebral OR cerebrovascular OR cerebro-vascular) PRE/1 (event OR outcome OR endpoint OR end-point OR "end point"))) OR (TITLE-ABS-KEY (adverse PRE/1 (cardiac OR cardiovascular OR cardio-vascular) PRE/1 (event OR outcome OR endpoint OR end-point))) OR (TITLE-ABS-KEY (MACCE)) OR (TITLE-ABS-KEY ((cardiovascular OR cardio-vascular OR cardiac OR cerebral OR cerebrovascular OR cerebro-vascular OR atherothrombotic OR athero-thrombotic OR atherothromb* OR athero-thromb* OR thrombotic OR thrombo* OR ischemic OR ischaemic) W/2 risk)) OR (TITLE-ABS-KEY (mortality)) OR (TITLE-ABS-KEY ((cardiovascular OR cardio-vascular OR cardiac OR cerebral OR cerebrovascular OR cerebro-vascular OR atherothrombotic OR athero-thrombotic OR atherothromb* OR athero-thromb* OR thrombotic OR thrombo* OR ischemic OR ischaemic) W/2 death)) OR (TITLE-ABS-KEY (proportional PRE/1 hazard PRE/1 (model OR analysis))) OR (TITLE-ABS-KEY (reinfarction)) OR (TITLE-ABS-KEY ("coronary restenosis")) OR (TITLE-ABS-KEY (coronary W/1 restenosis)) OR (TITLE-ABS-KEY (coronary W/1 re-stenosis)) OR (TITLE-ABS-KEY ("stent thrombosis")) OR (TITLE-ABS-KEY (stent W/1 thrombosis)) OR (TITLE-ABS-KEY ("kaplan-meir" PRE/2 (curve OR analysis))) OR (TITLE-ABS-KEY (survival PRE/0 (analysis OR rate))) OR (TITLE-ABS-KEY ("roc curve")) OR (TITLE-ABS-KEY ("area under curve")) OR (TITLE-ABS-KEY (revascularization w/2 ("target vessel" OR "target lesion"))))

#4 - #1 AND #2 AND #3

#### Supplementary Tables.

##### Supplementary Table 1. Additional sources.

| Name | Last search date | Search strategy | Number of results |
| --- | --- | --- | --- |
| Registries | | | |
| ClinicalTrials.gov https://clinicaltrials.gov/ | Mar/5/2024 | ("Mean Platelet Volume" OR "Platelet Size" OR "Large Platelet" OR "Platelet Volume") AND ("Acute Coronary Syndrome" OR "Myocardial Infarction" OR "Unstable angina" OR “Coronary Artery disease”) | 25 |
| The Australian New Zealand Clinical Trials Registry https://www.anzctr.org.au/ | Mar/5/2024 | platelet AND “Coronary Artery disease” | 183 |
| Chinese Clinical Trial Registry http://www.chictr.org.cn/abouten.aspx | Mar/5/2024 | “Platelet”; Filter: cohort studies | 362 |
| The European Union Clinical Trials Register | Mar/9/2024 | platelet AND "coronary artery disease" | 65 |

| | | | |
|---|---|---|---|
| https://www.clinicaltrialsregister.eu/ | | | |
| German Clinical Trial Registry https://www.drks.de/ | Mar/9/2024 | Platelet AND "coronary artery disease" | 24 |
| Japan Primary Registries Network https://rctportal.niph.go.jp/ | Mar/5/2024 | myocardial infarction AND platelet | 1 |
| Clinical Research Information Service, Republic of Korea https://cris.nih.go.kr/cris/ | Mar/5/2024 | Platelet; Filter: disease of the circulatory system | 5 |
| Iranian Registry of Clinical Trials https://www.irct.ir/ | Mar/9/2024 | Platelet AND "coronary artery disease" | 14 |
| Peruvian Clinical Trial Registry https://ensayosclinicos-repec.ins.gob.pe/en/ | Mar/5/2024 | Filter: cardiology<br>“myocardial infarction”<br>“acute coronary syndrome” | 1 |
| Brazilian Registry of Clinical Trials http://www.ensaiosclinicos.gov.br/ | Mar/9/2024 | platelet AND coronary artery disease | 10 |
| Clinical Trials Registry-India https://ctri.icmr.org.in/ | Mar/9/2024 | coronary artery disease | 426 |
| International Standard Randomised Controlled Trial Number registry https://www.isrctn.com/ | Mar/5/2024 | Platelet<br>Filter: Circulatory System | 142 |
| Journal websites | | | |
| Taylor and Francis journals https://www.tandfonline.com/ | Mar/9/2024 | "mean platelet volume" AND "coronary artery disease" | 374 |

| | | | |
|---|---|---|---|
| Nature journals https://www.nature.com/ | Mar/9/2024 | "mean platelet volume" AND "coronary artery disease" | 97 |
| PLOS ONE journal https://journals.plos.org/plosone/ | Mar/9/2024 | "mean platelet volume" AND "coronary artery disease" | 74 |
| SAGE publisher https://journals.sagepub.com/ | Mar/9/2024 | "mean platelet volume" AND "coronary artery disease" | 337 |
| The American Journal of Cardiology https://www.ajconline.org/ | Mar/9/2024 | "mean platelet volume"<br>Filters: Research Article | 1195 |
| Atherosclerosis https://www.atherosclerosis-journal.com/ | Mar/9/2024 | "mean platelet volume"<br>Filters: Research Article | 1017 |
| The International Journal of Cardiology https://www.internationaljournalofcardiology.com/ | Mar/9/2024 | "mean platelet volume"<br>Filters: Research Article | 635 |
| Blood Coagulation and Fibrinolysis https://journals.lww.com/bloodcoagulation/ | Mar/9/2024 | "mean platelet volume" AND "coronary artery disease"<br>Filters: Articles | 57 |
| Journals of American College of Cardiology https://www.jacc.org | Mar/9/2024 | "mean platelet volume"<br>Filters: Original Research | 2329 |
| Springer journals https://link.springer.com/ | Mar/9/2024 | "mean platelet volume" AND "coronary artery disease"<br>Filters: Article, Medicine and Public Health, Cardiology | 137 |
| Thrombosis Research https://www.thrombosisresearch.com | Mar/9/2024 | "mean platelet volume"<br>Filters: Research Article | 3056 |

| Other sources | | | |
|---|---|---|---|
| Biomed Explorer https://sites.research.google/biomedexplorer/ | Mar/9/2024 | “What is the prognostic role of mean platelet volume in coronary artery disease?” | 100 |
| Conferences from the European Society of Cardiology https://esc365.escardio.org/ | Mar/5/2024 | "mean platelet volume"<br>Filters: Topic — coronary artery disease, acute coronary syndromes, acute cardiac care, interventional cardiology and cardiovascular surgery, preventive cardiology | 3393 |
| TCT portal https://www.tctmd.com/see-all/conference | Mar/5/2024 | "mean platelet volume"<br>Filter: all conferences | 1 |
| CoCites tool https://www.cocites.com/ | Mar/9/2024 | CoCites plugin for web browsers were used for PubMed search. | 1520 |

**Supplementary Table 2. A list of some excluded studies with reasons for exclusion.**

| First author, year | Citation | Reasons for exclusion |
|---|---|---|
| Adam, 2018 | Adam AM, Rizvi AH, Haq A, Naseem R, Rehan A, Shaikh AT, Abbas AH, Godil A, Ali A, Mallick MSA, Khan MS, Lashari MN. Prognostic value of blood count parameters in patients with acute coronary syndrome. Indian Heart J. 2018 Mar-Apr;70(2):233-240. doi: 10.1016/j.ihj.2017.06.017. Epub 2017 Jun 30. PMID: 29716700; PMCID: PMC5993917. | It is a second publication related to the study cohort already included in the meta-analysis |
| Ayça B, 2015 | Ayça B, Akin F, Çelik Ö, Yüksel Y, Öztürk D, Tekiner F, Çetin Ş, Okuyan E, Dinçkal M H. Platelet to lymphocyte ratio as a prognostic marker in primary percutaneous coronary intervention. Platelets. 2015;26(7):638-44. doi: 10.3109/09537104.2014.968117. Epub 2014 Oct 28. PMID: 25350375. | The study provided an odds ratio for inhospital MACE with MPV treated as a continuous variable. There were only 2 studies for this analysis, therefore we excluded these studies |

| | | |
|---|---|---|
| Celik T, 2013 | Celik T, Kaya MG, Akpek M, Gunebakmaz O, Balta S, Sarli B, Duran M, Demirkol S, Uysal OK, Oguzhan A, Gibson CM. Predictive value of admission platelet volume indices for in-hospital major adverse cardiovascular events in acute ST-segment elevation myocardial infarction. Angiology. 2015 Feb;66(2):155-62. doi: 10.1177/0003319713513493. Epub 2013 Dec 3. PMID: 24301422. | The study provided an odds ratio for inhospital MACE with MPV treated as a continuous variable. There were only 2 studies for this analysis, therefore we excluded these studies |
| Ceylan, 2023 | Ceylan US, Yaman AE. Evaluation of the inflammatory parameters for predicting stent thrombosis. Bratisl Lek Listy. 2023;124(6):475-479. doi: 10.4149/BLL_2023_073. PMID: 36876384. | Wrong study design |
| Ebina T, 2021 | Ebina T, Tochihara S, Okazaki M, Koike K, Tsuto Y, Tayama M, Takanami Y, Hirose H, Horii M, Okada K, Matsuzawa Y, Maejima N, Iwahashi N, Hibi K, Kosuge M, Tamura K, Kimura K. Impact of red blood cell distribution width and mean platelet volume in patients with ST-segment elevation myocardial infarction. Heart Vessels. 2022 Mar;37(3):392-399. doi: 10.1007/s00380-021-01936-6. Epub 2021 Sep 13. PMID: 34518907. | The adjusted statistics for MPV were not provided |
| Ki, 2013 | Ki YJ, Park S, Ha SI, Choi DH, Song H. Usefulness of mean platelet volume as a biomarker for long-term clinical outcomes after percutaneous coronary intervention in Korean cohort: a comparable and additive predictive value to high-sensitivity cardiac troponin T and N-terminal pro-B type natriuretic peptide. Platelets. 2014;25(6):427-32. doi: 10.3109/09537104.2013.835393. Epub 2013 Oct 8. PMID: 24102424. | It is a publication related to a cohort of patients already included in the meta-analysis |
| Małyszczak, 2020 | Małyszczak A, Łukawska A, Dyląg I, Lis W, Mysiak A, Kuliczkowski W. Blood Platelet Count at Hospital Admission Impacts Long-Term Mortality in Patients with Acute Coronary Syndrome. Cardiology. 2020;145(3):148-154. doi: 10.1159/000505640. Epub 2020 Feb 4. PMID: 32018251. | The adjusted statistics for MPV were not provided |

| | | |
|---|---|---|
| Moon, 2016 | Moon AR, Choi DH, Jahng SY, Kim BB, Seo HJ, Kim SH, Ryu SW, Song H, Kim TH. High-sensitivity C-reactive protein and mean platelet volume as predictive values after percutaneous coronary intervention for long-term clinical outcomes: a comparable and additive study. Blood Coagul Fibrinolysis. 2016 Jan;27(1):70-6. doi: 10.1097/MBC.0000000000000398. PMID: 26340462. | It is a publication related to a cohort of patients already included in the meta-analysis |
| Ndrepepa G, 2022 | Ndrepepa G, Holdenrieder S, Kastrati A. Prognostic value of De Ritis ratio in patients with acute myocardial infarction. Clin Chim Acta. 2022 Oct 1;535:75-81. doi: 10.1016/j.cca.2022.08.016. Epub 2022 Aug 17. PMID: 35985502. | The adjusted statistics for MPV were not provided |
| Odeberg J, 2016 | Odeberg J, Freitag M, Forssell H, Vaara I, Persson ML, Odeberg H, Halling A, Råstam L, Lindblad U. Influence of pre-existing inflammation on the outcome of acute coronary syndrome: a cross-sectional study. BMJ Open. 2016 Jan 12;6(1):e009968. doi: 10.1136/bmjopen-2015-009968. PMID: 26758266; PMCID: PMC4716249. | Wrong study design |
| Sun XP, 2016 | Sun XP, Li J, Zhu WW, Li DB, Chen H, Li HW, Chen WM, Hua Q. Impact of Platelet-to-Lymphocyte Ratio on Clinical Outcomes in Patients With ST-Segment Elevation Myocardial Infarction. Angiology. 2017 Apr;68(4):346-353. doi: 10.1177/0003319716657258. Epub 2016 Jul 11. PMID: 27381032. | The study provided an odds ratio for long-term MACE with MPV treated as a continuous variable. There were only 2 studies for this analysis, therefore we excluded these studies |
| Turen S, 2023 | Turen S, Memic Sancar K. Predictive Value of the Prognostic Nutritional Index for Long-Term Mortality in Patients with Advanced Heart Failure. Acta Cardiol Sin. 2023 Jul;39(4):599-609. doi: 10.6515/ACS.202307_39(4).20221223A. PMID: 37456943; PMCID: PMC10346059. | Wrong population |
| Yang L, 2018 | Yang L, Wang H, Zhang Y, Han T, Wang W. The Prognostic Value of Lipoprotein-Associated | The study provided an odds ratio for long-term MACE with |

| | | |
|---|---|---|
| | Phospholipase A2 in the Long-Term Care of Patients With Acute Coronary Syndrome Undergoing Percutaneous Coronary Intervention. Clin Appl Thromb Hemost. 2018 Jul;24(5):822-827. doi: 10.1177/1076029617737837. Epub 2017 Nov 9. PMID: 29121808; PMCID: PMC6714881. | MPV treated as a continuous variable. There were only 2 studies for this analysis, therefore we excluded these studies |
| Zhao Y, 2022 | Zhao Y, Zhou P, Gao W, Zhong H, Chen Y, Chen W, Waresi M, Xie K, Shi H, Gong H, He G, Qiu Z, Luo X, Li J. Cilostazol combined with P2Y12 receptor inhibitors: A substitute antiplatelet regimen for aspirin-intolerant patients undergoing percutaneous coronary stent implantation. Clin Cardiol. 2022 Feb;45(2):189-197. doi: 10.1002/clc.23787. Epub 2022 Feb 4. PMID: 35120275; PMCID: PMC8860475. | The adjusted statistics for MPV were not provided |

**Supplementary Table 3. Baseline features of the included studies**

| Study, year | Osuna, 1998 | Huczek, 2005 | Estevez-Loureir, 2009 | Vakili, 2009 | Akpek, 2011 | Goncalves, 2011 | Taglieri, 2011 | Tekbas, 2011 | Dogan, 2012 | Lopez-Cuenca, 2012 | Vrsalovic, 2012 | Eisen, 2013 | Akgul, 2013 | Fabregat-Andreas, 2013 | Nozari, 2013 | Unal, 2013 |
|---|---|---|---|---|---|---|---|---|---|---|---|---|---|---|---|---|
| Country | Spain | Poland | Spain | Iran | Turkey | Australia | Italy | Turkey | Turkey | Spain | Croatia | Israel | Turkey | Spain | Iran | Turkey |
| Study design | - | Prospective cohort | Prospective cohort | Retrospective cohort | Prospective cohort | Prospective cohort | Retrospective cohort | Retrospective cohort | Prospective cohort | Prospective cohort | Prospective cohort | Retrospective cohort | Prospective cohort | Retrospective cohort | Retrospective cohort | Prospective cohort |
| Follow-up, months | inhospital | 6 | 1 | inhospital | inhospital | 12 | 12 | 24.2 | 12 | 6 | 1 | 48 | 6 | 12 | 12 | 2 |
| Sample size, n | 1082 | 388 | 617 | 203 | 289 | 1102 | 1041 | 429 | 344 | 329 | 543 | 4961 | 495 | 128 | 2539 | 205 |
| Diagnosis | STEMI, 100% | STEMI, 100% | STEMI, 100% | STEMI, 100% | STEMI, 100% | ACS, 100% | NST-ACS, 100% | STEMI 65%; NSTEMI 35% | NSTEMI 37%, UA 63% | NST-ACS, 100% | STEMI, 100% | ACS, 100% | STEMI, 100% | STEMI, 100% | Stable CAD patients treated with PCI | Stable CAD patients |

| | | | | | | | | | | | | | | | | |
|---|---|---|---|---|---|---|---|---|---|---|---|---|---|---|---|---|
| Mean age, years | 68±13 | 60±11.3 | 63±12 | 56±11.2 | 58±12 | 62.8±11.5 | 76 (67-82) | 61.9±12.4 | 61.9±7.7 | 67.3±12.3 | 67.8 | 67.1±12.5 | 55.6±12.4 | 59.5±14.1 | 58±10.6 | 60.9±10.3 |
| Males, % | 78 | 72.2 | 81 | 78.8 | 80 | 72.8 | 74.2 | 70.4 | 69.6 | 64 | 63.4 | 75.5 | 80.2 | 80.2 | 69.8 | 82 |
| Hypertension, % | 43.5 | 62.6 | 37.5 | 31.5 | 44.7 | 60.2 | 79.7 | 43.4 | 49.1 | 69.4 | 58.4 | 70.5 | 34.3 | 59.2 | 50.1 | 62,9 |
| Diabetes mellitus, % | 19.8 | 16.5 | 17.5 | 22.7 | 28.3 | 27.9 | 25.4 | 52.2 | 34.1 | 40.6 | 27.4 | 38.7 | 20.2 | 37.9 | 27.5 | 35,6 |
| Dyslipidemia, % | 18.9 | 42 | 33.5 | 36.9 | - | 71.5 | 56.7 | 18.9 | 44.1 | 55.7 | 31.1 | - | - | 53 | 67.4 | 54.6 |
| Smoking, % | 55.2 | 55.2 | 35 | 47.8 | 60.6 | 15.9 | 43.5 | 43.4 | 44.9 | 58 | 52.3 | 40.5 | - | 55.5 | 28.3 | - |
| Previous MI, % | 36.3 | 20.1 | - | 12.3 | - | 24.7 | 38.5 | 10.5 | - | 17.2 | - | - | 16.6 | - | 49.6 | - |
| Family history of CAD, % | - | - | 6.5 | - | - | - | 16.3 | 20.7 | 21.1 | - | - | - | - | - | 20 | - |

| | | | | | | | | | | | | | | | | |
|---|---|---|---|---|---|---|---|---|---|---|---|---|---|---|---|---|
| History of PCI, % | - | - | - | - | - | - | 23.5 | - | - | 17 | - | - | - | - | 0 | - |
| BMI, kg/m$^2$ | - | - | - | - | 26.1±2.6 | - | - | - | - | - | 27.8 | - | - | - | - | - |
| Baseline SBP, mmHg | - | 129.3±19.8 | - | 119.7±27 | - | - | 140 (125-160) | 124.9±21.6 | - | - | - | - | 134.8±58.5 | - | - | - |
| Glucose, mg/dl | - | - | - | - | 176.5±97.1 | - | - | 147.7±77.7 | - | - | - | - | 168.9±83.3 | 112.5±36 | - | - |
| Cholesterol, mg/dl | - | - | 190.8±46 | - | 179.5±43 | - | - | - | - | - | - | - | 196.4±46.8 | 166.2±50 | - | - |
| LDL-C, mg/dl | - | - | 119.8±39.1 | - | 117.9±32.8 | - | - | - | - | - | - | - | 131.6±36.7 | 113.9±36 | - | - |
| HDL-C, mg/dl | - | - | 47.7±12,7 | - | 39.0±113.3 | - | - | - | - | - | - | - | 40.9±10.3 | 37.2± | - | - |
| Fasting triglycerides, mg/dl | - | - | - | - | 115.1±60.8 | - | - | - | - | - | - | - | 128.3±89 | - | - | - |
| Creatinine, mg/dl | - | - | 0.98 | - | - | 1.14±0.73 | 1.2 (1.02-1.52) | 1.16±0.4 | 1.1 | 0.95 (0.80-1.13) | 1.25 | 1.05±0.7 | 0.9±0.4 | 0.9±0.2 | 1.06±0.4 | 0.93 [0.80-1.07] |

| | | | | | | | | | | | | | | | | |
|---|---|---|---|---|---|---|---|---|---|---|---|---|---|---|---|---|
| WBC, $10^9$/l | - | - | 11.0 | - | 12.3±4.7 | - | 8.6 (6.9-10.9) | 11.8±4.2 | 8.0 | 8.1 (6.7-9.9) | 10.5 | - | 12.3±4.2 | - | - | 8.27±1.93 |
| Hemoglobin, g/l | - | - | - | - | 144.1±17.6 | 138.2±16.3 | 134 (120-148) | 13.3±1.8 | 134.3±1.3 | 138±18 | - | 135±17 | - | 137±16 | 139.8±1.7 | 14.4±1.6 |
| Platelet count, $10^9$/l | - | - | 257.5±101.1 | - | 243.7±65.8 | 247±74.1 | 241 (199-299) | 277.7±85.3 | 265 | 209 (174-252) | 246.5 | 243.7±71.1 | 257±71.4 | 233±62 | 226.3±63.3 | 268±67 |
| MPV, fl | 8.8±1.03 | 10.0±0.9 | 8.8 | 9.55±0.88 | 10.4±0.9 | 8.7±1.3 | - | 10.4±2 | | 11 (10.3-11.8) | - | 8.55±1.1 | 8.4±1.1 | 9.1±1.1 | - | - |
| EF, % | - | - | 57±14 | - | 48.4±10.6 | - | - | 50.3±9.4 | 53.4 | - | 53.7 | - | - | 52.1±10.8 | - | 55.0 [50-60] |
| PCI, % | - | 100 | 100 | 100 | 100 | - | 68.4 | - | 61.3 | - | 100 | 100 | 100 | 100 | 100 | - |
| Anterior MI, % | - | 43.8 | 41 | 65.1 | - | - | - | - | - | - | 40.7 | - | 44.4 | 40.3 | - | - |
| MVD, % | - | 22.7 | 48 | - | 51.9 | - | 56.1 | - | 45.6 | - | 51.8 | 77 | - | - | - | - |
| GPI use, % | - | 52.1 | 61 | 10.9 | 15.6 | - | - | - | - | - | - | - | 33.3 | - | - | - |

**(Continued) Supplementary Table 3.** Baseline features of the included studies.

| Study, year | Bergoli ,2014 | **Lekston, 2014** | Choi, 2014 | Liu, 2014 | Seo, 2014 | Seyyed-Mohammadza, 2014 | Siller-Matula , 2014 | Wan, 2014 | Ghaffari, 2015 | Lai, 2015 | Navarta, 2015 | Ranjith, 2015 | Lai, 2016 | Wasilewski, 2016 | Tongtong Yu, 2017 | Gina Yu, 2017 |
|---|---|---|---|---|---|---|---|---|---|---|---|---|---|---|---|---|
| Country | Brazil | Poland | Korea | China | South Korea | Iran | Austria | China | Iran | China | Argentina | India | China | Poland | China | China |
| Study design | Prospective cohort | Retrospective cohort | Retrospective cohort | Prospective cohort | Retrospective cohort | Retrospective cohort | Prospective cohort | Prospective cohort | Prospective cohort | Retrospective cohort | Prospective cohort | Prospective cohort | Prospective cohort | Retrospective cohort | Retrospective cohort | Retrospective cohort |
| Follow-up, months | 1 | 12 | 12 | inhospital | 25.8 | 12 | 24 | 52 | 6 | 1 | 10.1 | 12 | 1 | 12 | 28 | 1 |
| Sample size, n | 168 | 1557 | 122 | 190 | 372 | 181 | 404 | 286 | 191 | 649 | 250 | 1206 | 453 | 1001 | 887 | 608 |
| Diagnosis | STEMI ,100% | STEMI , 100% | ACS, 100% | NSTEMI, 100% | All CAD patients treate | Stable CAD patients treate | All CAD patients treated | ACS | STEMI, 100% | STEMI, 100% | STEMI, 22%; NSTE | STEMI, 66.09; NSTE | STEMI, 100% | NSTEMI, 100% | NSTEMI, 100% | STEMI, 100% |

| | | | | | d with PCI | d with PCI | with PCI | | | | MI, 26.8 %; UA 51.2 | MI, 33.91 % | | | | |
|---|---|---|---|---|---|---|---|---|---|---|---|---|---|---|---|---|
| Mean age, years | 60.7±12.7 | 61.9±9.9 | 65.4±11.6 | 66.8±12.7 | 65.2±10.7 | 58.2±10.9 | 64.4±12 | 59.4±12 | 60±11 | 59.1±11.8 | 74±7 | 56±11 | 56.2±8.4 | 64.7 ± 10.7 | 62.5±11.6 | 62.7±13.1 |
| Males, % | 66.3 | 67.6 | 66.8 | 65.8 | 63.2 | 68.5 | 51.5 | 61.2 | 82.2 | 82.3 | 56.4 | 77.4 | 84.5 | 66.6 | 67.3 | 79.1 |
| Hypertension, % | 61.3 | 63.8 | 54.8 | 70 | 57.3 | 49.2 | 84.4 | 41.9 | 37.7 | 45.8 | 81.6 | 34.7 | 41.5 | 65.3 | 60.6 | 49.7 |
| Diabetes mellitus, % | 15.6 | 34.6 | 32.7 | 32.1 | 33.3 | 16.6 | 31.9 | 21.3 | 22 | 32.5 | 26 | 30.0 | 25.2 | 30.3 | 35.8 | - |
| Dyslipidemia, % | - | - | 7.7 | 17.4 | 25.8 | 48.1 | 76.2 | - | 16.2 | 74 | 58.8 | 23.6 | - | 31.2 | 71 | 13 |
| Smoking, % | 68.4 | - | 49 | 36.9 | 48.4 | 44.8 | 55.2 | 46.8 | 40.8 | 30.7 | 26 | 41.3 | 29.8 | 21.1 | 48.8 | - |
| Previous MI, % | 7.1 | 28.3 | - | 12.1 | - | 39.2 | 32.2 | - | 0 | - | 20.8 | - | - | 34.7 | 9.1 | - |

| | | | | | | | | | | | | | | | | |
|---|---|---|---|---|---|---|---|---|---|---|---|---|---|---|---|---|
| Family history of CAD, % | - | - | 1.4 | - | 2.2 | 29.8 | - | - | 7.9 | - | - | - | 15.5 | 24.6 | - | - |
| History of PCI, % | 9.6 | - | - | 11.6 | - | 18.2 | - | - | - | - | 18.4 | - | 4.2 | 26.8 | 11 | 11.5 |
| BMI, kg/m$^2$ | - | 26±1.9 | - | - | 24.2±3.4 | 27.4±4.4 | 28±5 | - | - | 25.8±3.6 | - | - | 26±3.5 | 28.6±4.8 | - | - |
| Baseline SBP, mmHg | - | - | - | - | 133.9±22.2 | - | - | 120.4±17.6 | - | 121.3±20.4 | 146±29 | - | 121.1±19.7 | 150.8±28.7 | 137.3±23.4 | 127±38.8 |
| Glucose, mg/dl | - | 156.8±71 | - | - | 119.8±37.6 | - | - | 102.7±39.6 | 171±91 | 113.5±42.3 | - | - | 100.9±31.5 | 116.4 | - | - |
| Cholesterol, mg/dl | - | 231.7 | - | - | 151.8±36.7 | 157.5±34.2 | - | 150.2±27.6 | - | 183.4±37.1 | - | 185.5±45.4 | 184.6±32.8 | - | - | - |
| LDL-C, mg/dl | - | 146.7 | - | - | 88.8±30 | 86.7±25 | - | 107.48±39.9 | - | 101.2±33.2 | - | 119.7±54.3 | 102.3±18.2 | - | - | - |
| HDL-C, mg/dl | - | 54.1 | - | - | 44.3±11 | 40.6±9.1 | - | 33.6±8 | - | 43.2±22.4 | - | - | 41.3±8.9 | - | - | - |
| Fasting triglyce | - | 123.9 | - | - | 135.8±78.1 | 172.6±87.7 | - | 131.9±71.3 | - | 144.3±80.5 | - | 116.0±44.8 | 150.4±64.6 | - | - | 115.9±132.3 |

| | | | | | | | | | | | | | | | | |
|---|---|---|---|---|---|---|---|---|---|---|---|---|---|---|---|---|
| rides, mg/dl | | | | | | | | | | | | | | | | |
| Creatinine, mg/dl | 1.0±0.8 | 1.24 | 1.2±1.0 | - | 1.17±0.58 | 0.9±0.5 | - | - | 1.2±1.2 | - | 1.0±0.5 | 1.1±0.4 | 1±0.2 | 1.05 | 1.04±0.4 | 1.2±1.0 |
| WBC, $10^9$/l | - | 11.9±5 | - | - | - | - | - | - | 12.3±2.1 | 9.7±2.8 | 8.85±3.8 | - | 10.3±3.7 | 9.2 | 7.6±2.4 | 11.6±4 |
| Hemoglobin, g/l | - | 144.6±13.9 | 133±2 | - | - | - | 141.7±9 | - | - | 138±13.6 | - | - | 138.2±13.4 | 136.6±17.5 | 133±17.7 | 143.6±2 |
| Platelet count, $10^9$/l | - | 207±66.9 | 252±72 | 194.8±56.3 | - | 209±56.9 | 227.6±72 | - | 242.7±56.8 | 211.7±52.1 | 210±75 | 238.5±65.7 | 220.8±65.5 | - | 202.3±56.3 | 246±77 |
| MPV, fl | - | 9.7 | 8.61±0.9 | 9.5±1.6 | - | - | - | 11.5±1.1 | 8.2±1.8 | 10.5±1.2 | 10.6±0.6 | 9.2±1.0 | 9.8±0.9 | 11±1.1 | 9.4±1.5 | - |
| EF, % | - | 44.6 | 58.8±11.2 | 58.6±7.6 | 59.8±11.8 | - | - | 59.7±13 | - | 50.8±5.4 | - | 57.5±10.5 | 49.6±6 | 43.9±10 | 57.1±10 | 44.6±12.2 |
| PCI, % | 100 | - | 100 | - | 100 | 100 | 100 | - | 48.7 | 100 | 47.2 | - | 100 | 100 | 100 | - |
| Anterior MI, % | 47.3 | | - | 23.7 | - | - | - | - | 71.7 | 52.7 | - | - | 47.2 | - | - | - |
| MVD, % | - | 44.4 | - | - | - | - | - | - | 47.4 | 71.3 | - | - | 41.9 | 68.4 | - | - |

| | | | | | | | | | | | | | | | | |
|---|---|---|---|---|---|---|---|---|---|---|---|---|---|---|---|---|
| GPI use, % | 58 | - | - | - | - | - | - | - | - | 42.7 | - | - | 43.7 | 8.7 | 25.5 | - |

**(Continued) Supplementary Table 3.** Baseline features of the included studies.

| Study, year | Adam, 2018 | Mach ado, 2018 | Monte iro Júnior , 2018 | Tian, 2018 | Niu, 2018 | Wada 2018 | Cang a, 2019 | Chan g, 2019 | Garlob o, 2019 | Navart a, 2019 | Nozari , 2019 | Satıro glu, 2019 | Jiang, 2019 | Vogia tzis, 2019 | Xinsen , 2020 |
|---|---|---|---|---|---|---|---|---|---|---|---|---|---|---|---|
| Country | Pakist an | Brazi l | Brazil | China | China | Japan | Turke y | Taiwa n | Cuba | Argent ina | Iran | Turke y | China | Greec e | China |
| Study design | Prospe ctive cohort | Prosp ective cohor t | Prosp ective cohort | Retro specti ve cohor t | Retros pectiv e cohort | Prosp ective cohor t | Retro specti ve cohor t | Prosp ective cohor t | - | Prospe ctive cohort | Retros pectiv e cohort | Retro specti ve cohor t | Prosp ective cohor t | Retro specti ve cohor t | Retros pective cohort |
| Follow-up, months | 1 | in-ho spital | in-hos pital | 30 | 12 | 67.2 | 1 | 28.8 | in-hos pital | 8.6 | 12 | in-ho spital | 24 | in-ho spital | in-hosp ital |
| Sample size, n | 250 | 625 | 466 | 1215 | 2693 | 2872 | 349 | 1094 | 188 | 195 | 4199 | 194 | 1389 | 104 | 516 |

| Diagnosis | STEMI, 39.6%; NSTEMI, 26.4%; UA 34 | STEMI, 100% | STEMI, 70; NSTEMI, 30% | STEMI, 100% | STEMI, 49.43; NST-ACS, 50.57 % | Stable CAD patients treated with PCI | STEMI, 100% | STEMI, 27.25 %; NSTEMI, 44.52 %; UA, 28.23 % | Acute MI, 100% | STEMI, 20.51 %; NST-ACS, 79.49 % | NST-ACS, 100% | STEMI , 100% | Stable CAD patients treated with PCI | STEMI, 32.7 %; NSTEMI, 29.8 %; UA, 37.5 % | STEMI, 100% |
|---|---|---|---|---|---|---|---|---|---|---|---|---|---|---|---|
| Mean age, years | 55.1±10.7 | 60.7±12.1 | 64.2±12.8 | 61.5±12.4 | 61.1±11.8 | 67.0±10.0 | 36.4±3.6 | 69±13 | - | 74±7 | 59.9±10.3 | 78.5±4.7 | 59±9.5 | 64.2±11.1 | 65.4±8.6 |
| Males, % | 65.2 | 67.5 | 61.6 | 73.9 | 75.8 | 83.1 | 90 | 71.1 | 69.7 | 57 | 65.7 | 53 | 76.2 | 76 | 57.6 |
| Hypertension, % | 70.4 | 59 | 71.9 | 48.6 | 47.4 | 74.3 | 14 | 68.3 | 80.9 | 80 | 59.8 | 55.2 | 69 | - | 61.8 |
| Diabetes mellitus, % | 34 | 23.7 | 37.1 | 32.3 | 20.8 | 46.2 | 13.2 | 51.4 | 27.7 | 26.7 | 32.6 | 38.7 | 100 | - | 49.6 |
| Dyslipidemia, % | - | - | 38.2 | 66 | 70.8 | 74.9 | 12.3 | 53.4 | 4.2 | 56.9 | 68.7 | 44.8 | 74,7 | - | 19.4 |

| | | | | | | | | | | | | | | | |
|---|---|---|---|---|---|---|---|---|---|---|---|---|---|---|---|
| Smoking, % | 29.2 | 63.1 | 41.6 | 56.6 | 50.4 | 22.9 | 81.7 | 28.7 | 58.5 | 20.5 | 22.4 | 8.8 | 53.9 | - | 52.7 |
| Previous MI, % | - | 9.0 | - | 4.6 | 10.8 | - | - | - | - | 11.8 | - | - | 26.5 | - | - |
| Family history of CAD, % | 40 | - | 47.2 | - | - | 28.2 | 14 | - | - | - | 18.1 | - | - | - | |
| History of PCI, % | - | 10.9 | - | 4 | - | - | - | - | - | 11.8 | 15 | - | 30.6 | - | - |
| BMI, kg/m$^2$ | - | 27.32±8.68 | - | - | 24.1±2.8 | 24.3±3.4 | - | 25±4 | - | - | 28±4.5 | - | 26.3±3.2 | - | - |
| Baseline SBP, mmHg | 129.4±29.4 | - | - | 128±22 | 124.1±17.4 | - | 127.6±17.7 | - | - | - | - | - | 129.4±16.8 | - | 125.2±19.1 |
| Glucose, mg/dl | - | - | - | - | 113.8±39.2 | 108.4±33.2 | 128.8 ± 60.4 | - | - | 129.6±36.6 | 103.0 (91.0, 132.0) | 117.3±48.4 | 135.14±43.24 | - | 98.2±39.1 |
| Cholesterol, mg/dl | - | - | - | - | - | - | 184.4±51.8 | - | - | - | 173.9 ± 47.1 | 176.5±41.4 | 152.51±38.61 | - | 162.21±44.08 |

| | | | | | | | | | | | | | | | |
|---|---|---|---|---|---|---|---|---|---|---|---|---|---|---|---|
| LDL-C, mg/dl | - | - | - | - | 97.7±33.7 | 103.5±30.5 | 114.9 ± 42.8 | 112±41 | - | - | 102.0 (78.5, 131.0) | 111±35.6 | 87.64±30.12 | - | 96.53 |
| HDL-C, mg/dl | - | - | - | - | - | 43.9±12.7 | 34.5 ± 8.5 | - | - | - | 40.9±10.8 | 39.2±9.3 | 37.84±8.88 | - | 37.45 |
| Fasting triglycerides, mg/dl | - | - | - | - | - | 134.9±81.4 | 141±106.0 | - | - | - | 147.0 (106.5, 209.0) | 111.2±50.6 | 139.82±65.49 | - | 162.57±106.85 |
| Creatinine, mg/dl | - | - | - | 0.96±0.54 | 0.99±0.2 | - | 0.83 ± 0.29 | 2.13±2.5 | - | 1.06±0.6 | 0.9 (0.8, 1.1) | 0.9±0.3 | - | 1.0±0.3 | 0.83±0.46 |
| WBC, $10^9$/l | 10.5±3.2 | - | 10.5 (8.4, 12.8) | 9.5±3.2 | 7.7±2.2 | 5.8±1.9 | 12.0 ± 3.7 | 10.5±4.8 | - | 8.3±3.1 | - | 9.9±3.7 | 6.72±1.73 | - | - |
| Hemoglobin, g/l | 124.8±1.6 | - | 130±2 | 135.2±17.8 | 147.8±19.7 | 132±18 | - | 128±25 | - | - | 140 ± 17 | 131.5±16.9 | 142.7±15.6 | - | - |
| Platelet count, $10^9$/l | 259.7±65.6 | - | 231 (195.7, 278) | 203.1±56.3 | 177.7±60.2 | 204±58 | 244.3±65.1 | 217±75 | - | 207±45.7 | - | 241±65 | - | - | |
| MPV, fl | 11.1±1.4 | 10.7 (10–11.3) | 10.9±0.9 | 9.5±1.6 | 11.6±1.3 | - | 8.0±1.2 | 8.6±1.1 | - | 10.5±0.5 | - | 8.2±1.3 | - | 10.7±1.2 | 10.7±0.91 |
| EF, % | 46±11.2 | - | - | 54.9±9.4 | 55.4±6.1 | 61.7±12.0 | 47.4±9.9 | - | - | - | 50.6±9.2 | - | 63.3±7.3 | - | 49.8±9.5 |

| | | | | | | | | | | | | | | | |
|---|---|---|---|---|---|---|---|---|---|---|---|---|---|---|---|
| PCI, % | - | 100 | 48.5 | 100 | - | 100 | 100 | - | - | - | 100 | 100 | 100 | - | - |
| Anterior MI, % | - | 44.8 | - | 50.5 | - | - | 57.9 | - | - | - | - | 53.1 | - | - | |
| MVD, % | 67.6 | - | - | - | - | - | 29.2 | - | - | - | - | 46.4 | - | - | - |
| GPI use, % | - | - | - | 46 | - | - | 44.7 | - | - | - | - | - | - | - | |

**(Continued) Supplementary Table 3.** Baseline features of the included studies.

| Study, year | Chen Xinsen, 2020 | Adali, 2022 | Liang, 2023 | Pedersen, 2023 | Toprak 2023 |
|---|---|---|---|---|---|
| Country | China | Turkey | China | Denmark | Turkey |
| Study design | Retrospective cohort | Retrospective cohort | Retrospective cohort | Prospective cohort | Retrospective cohort |
| Follow-up, months | - | 31.5 | 12 | 37.2 | 1 |
| Sample size, n | 524 | 189 | 296 | 900 | 1202 |

| Diagnosis | STEMI patients treated with PCI | Stable CAD patients | NST-ACS patients | Stable CAD patients | STEMI patients treated with PCI |
|---|---|---|---|---|---|
| Mean age, years | 58.9±9.1 | 68.2±10.7 | 61.4±6.9 | 65±4 | 59.9±11.5 |
| Males, % | 57.1 | 81.4 | 69.9 | 78 | 69.2 |
| Hypertension, % | 64.9 | - | 48.3 | - | 66.3 |
| Diabetes mellitus, % | 44.7 | - | 20.3 | 28 | 34.4 |
| Dyslipidemia, % | 58.2 | 8.9 | - | 90 | 37.6 |
| Smoking, % | 55.2 | 21.1 | 50.9 | 22 | 70.5 |
| Previous MI, % | - | 46 | - | 88 | - |
| Family history of CAD, % | | 8.5 | - | - | 9 |
| History of PCI, % | - | - | - | - | - |
| BMI, $kg/m^2$ | - | - | 26.6±2.8 | 27±8 | 26.5±2 |
| Baseline SBP, mmHg | - | 127.6±17.8 | - | - | 134.1±19 |

| | | | | | |
|---|---|---|---|---|---|
| Glucose, mg/dl | 174.4±99.6 | - | 113.51±30.63 | - | 117±36 |
| Cholesterol, mg/dl | 160.8±32.2 | - | | - | 175.9±44.5 |
| LDL-C, mg/dl | 94.8±23.8 | - | 92.7±32 | - | 107.1±36 |
| HDL-C, mg/dl | 37.71±7.64 | - | 51.35±21.62 | - | 34.3±7.5 |
| Fasting triglycerides,mg/dl | 150.29±78 | - | 159.29 | - | 151.2±79.9 |
| Creatinine, mg/dl | - | - | 0.8±0.5 | 0.93±0.32 | 0.8 [0.7-1.0] |
| WBC, $10^9$/l | - | - | 7.9±3.3 | 7.1±2 | 8.0±3.8 |
| Hemoglobin, g/l | 229±65.4 | - | - | 8.8±0.8 | 136±17 |
| Platelet count, $10^9$/l | 240.5±63.6 | - | 196.6±17.6 | 233±58 | 247.7±70.6 |
| MPV, fl | - | - | - | - | - |
| EF, % | 56.6±9.7 | 46.3±12.4 | - | - | 50.4±13.7 |
| PCI, % | - | - | - | - | 100 |
| Anterior MI, % | - | - | - | - | - |

| | | | | | |
|---|---|---|---|---|---|
| MVD, % | - | 70.9 | - | - | - |
| GPI use, % | | - | - | - | - |

Abbreviations: ACS, acute coronary syndrome; BMI, body mass index; CAD, coronary artery disease; EF, ejection fraction; GPI, glycoprotein inhibitor use; HDL-C, high-density lipoproteins; LDL-C, low-density lipoproteins; MI, myocardial infarction; MPV, mean platelet volume; MVD, multivessel disease; NST-ACS, non-ST-elevation acute coronary syndrome; NSTEMI, non-ST-elevation myocardial infarction; PCI, percutaneous coronary intervention; SBP, systolic blood pressure; STEMI, ST-elevation myocardial infarction; UA, unstable angina; WBC, white blood cells. Data are presented as mean ± standard deviation or as median (interquartile range)/ median.

**Supplementary Table 4. Leave-one out sensitivity analyses.**

| **Omitting this study - first author, year** | **ML estimation** | | | **REML estimation** | | |
|---|---|---|---|---|---|---|
| | **Estimate [95% CI]** | **$I^2$** | **$tau^2$** | **Estimate [95% CI]** | **$I^2$** | **$tau^2$** |
| **All ischemic heart disease patients** | | | | | | |
| MPV as a continuous variable for long-term mortality with HR as an effect estimate | | | | | | |
| Taglieri, 2011 | 1.30 [1.21-1.38] | 37.84 | 0.00 | 1.30 [1.21-1.39] | 47.44 | 0.00 |
| Eisen ACS subgroup, 2013 | 1.32 [1.26-1.38] | 0.01 | 0.00 | 1.32 [1.26-1.38] | 0.01 | 0.00 |
| Eisen SCAD subgroup, 2013 | 1.30 [1.21-1.39] | 38.12 | 0.00 | 1.30 [1.21-1.40] | 46.98 | 0.00 |
| Lekston, 2014 | 1.26 [1.18-1.35] | 15.32 | 0.00 | 1.27 [1.19-1.37] | 28.85 | 0.00 |
| Wasilewski, 2016 | 1.29 [1.20-1.37] | 36.23 | 0.00 | 1.29 [1.20-1.38] | 45.49 | 0.00 |
| Yu, 2017 | 1.29 [1.21-1.38] | 38.09 | 0.00 | 1.30 [1.21-1.39] | 47.88 | 0.00 |
| Tian, 2018 | 1.28 [1.21-1.35] | 29.74 | 0.00 | 1.28 [1.21-1.35] | 37.00 | 0.00 |
| Jiang, 2019 | 1.29 [1.22-1.36] | 35.07 | 0.00 | 1.29 [1.22-1.36] | 43.15 | 0.00 |
| Adali, 2022 | 1.29 [1.21-1.37] | 35.62 | 0.00 | 1.29 [1.21-1.37] | 44.17 | 0.00 |
| MPV as a categorical variable for long-term mortality with HR as an effect estimate | | | | | | |
| Taglieri, 2011 | 1.74 [1.07-2.84] | 0.00 | 0.00 | 2.95 [0.55-15.67] | 69.57 | 0.36 |
| Tekbas, 2011 | 2.66 [0.38-18.66] | 59.09 | 0.27 | 2.87 [0.41-20.13] | 72.53 | 0.49 |
| Akgul, 2013 | 1.68 [1.22-2.33] | 0.01 | 0.00 | 1.68 [1.22-2.33] | 0.00 | 0.00 |
| Jiang, 2019 | 1.70 [1.12-2.58] | 0.01 | 0.00 | 1.94 [0.50-7.44] | 81.88 | 0.16 |
| MPV as a continuous variable for long-term MACE with HR as an effect estimate | | | | | | |
| Taglieri, 2011 | 1.15 [1.04-1.28] | 94.78 | 0.02 | 1.16 [1.04-1.28] | 95.29 | 0.02 |
| Eisen ACS subgroup, 2013 | 1.16 [1.05-1.29] | 94.04 | 0.02 | 1.16 [1.05-1.29] | 94.61 | 0.02 |
| Eisen SCAD subgroup, 2013 | 1.16 [1.05-1.29] | 94.51 | 0.02 | 1.16 [1.05-1.29] | 95.04 | 0.02 |

| | | | | | | |
|---|---|---|---|---|---|---|
| Lopez-Cuenca , 2012 | 1.15 [1.04-1.27] | 94.56 | 0.02 | 1.15 [1.04-1.27] | 95.11 | 0.02 |
| Wan, 2014 | 1.16 [1.05-1.29] | 90.85 | 0.02 | 1.16 [1.05-1.29] | 91.70 | 0.02 |
| Siller-Matula, 2014 | 1.17 [1.05-1.29] | 94.61 | 0.02 | 1.17 [1.06-1.29] | 95.15 | 0.02 |
| Wasilewski, 2016 | 1.16 [1.05-1.29] | 94.80 | 0.02 | 1.16 [1.05-1.29] | 95.31 | 0.02 |
| Yu, 2017 | 1.15 [1.04-1.27] | 94.46 | 0.02 | 1.15 [1.04-1.27] | 95.03 | 0.02 |
| Tian, 2018 | 1.14 [1.03-1.25] | 93.44 | 0.02 | 1.14 [1.03-1.25] | 94.19 | 0.02 |
| Niu, 2018 | 1.12 [1.04-1.22] | 90.65 | 0.01 | 1.13 [1.04-1.22] | 91.69 | 0.01 |
| Wada, 2018 | 1.18 [1.08-1.28] | 91.15 | 0.01 | 1.18 [1.08-1.29] | 92.28 | 0.01 |
| Navarta, 2019 | 1.17 [1.06-1.29] | 91.02 | 0.02 | 1.17 [1.06-1.30] | 91.89 | 0.02 |
| Chen Xinsen, 2020 | 1.15 [1.04-1.27] | 94.50 | 0.02 | 1.15 [1.04-1.27] | 95.06 | 0.02 |
| Pedersen, 2023 | 1.17 [1.07-1.29] | 94.04 | 0.02 | 1.17 [1.07-1.29] | 94.62 | 0.02 |
| MPV as a categorized variable for long-term MACE with HR as an effect estimate | | | | | | |
| Taglieri, 2011 | 2.23 [1.36-3.65] | 86.05 | 0.00 | 2.25 [1.37-3.70] | 87.52 | 0.00 |
| Dogan, 2012 | 2.21 [1.34-3.63] | 87.51 | 0.01 | 2.23 [1.35-3.68] | 88.85 | 0.01 |
| Lopez-Cuenca, 2012 | 2.17 [1.32-3.58] | 88.25 | 0.01 | 2.20 [1.33-3.63] | 89.55 | 0.01 |
| Fabregat-Andres, 2013 | 2.05 [1.27-3.33] | 87.74 | 0.01 | 2.08 [1.27-3.38] | 89.13 | 0.01 |
| Choi, 2014 | 2.02 [1.29-3.15] | 86.97 | 0.01 | 2.03 [1.30-3.17] | 88.31 | 0.01 |
| Seo, 2014 | 1.98 [1.23-3.17] | 86.47 | 0.01 | 2.00 [1.24-3.23] | 88.06 | 0.01 |
| Navarta, 2015 | 1.84 [1.22-2.78] | 82.14 | 0.01 | 1.87 [1.23-2.83] | 84.1 | 0.01 |
| Niu, 2018 | 2.05 [1.25-3.37] | 87.19 | 0.01 | 2.07 [1.26-3.42] | 88.66 | 0.01 |
| Wada, 2018 | 2.31 [1.54-3.46] | 76.47 | 0.00 | 2.34 [1.56-3.52] | 79.34 | 0.00 |
| Chang, 2019 | 2.25 [1.38-3.67] | 84.88 | 0.00 | 2.27 [1.39-3.71] | 86.45 | 0.00 |
| Navarta , 2019 | 2.02 [1.25-3.26] | 87.27 | 0.01 | 2.04 [1.26-3.31] | 88.73 | 0.01 |
| Nozari, 2019 | 2.19 [1.33-3.60] | 88.00 | 0.01 | 2.21 [1.34-3.66] | 89.30 | 0.01 |
| MPV as a categorized variable for in-hospital mortality with OR as an effect estimate | | | | | | |
| Tekbas, 2011 | 1.48 [0.82-2.65] | 0.00 | 0.10 | 1.51 [0.77-2.97] | 8.19 | 0.12 |

| | | | | | | |
|---|---|---|---|---|---|---|
| Ranjith, 2015 | 1.74 [0.88-3.44] | 31.97 | 0.07 | 1.78 [0.84-3.75] | 58.83 | 0.08 |
| Monteiro Junior , 2018 | 1.66 [0.97-2.85] | 27.21 | 0.06 | 1.65 [0.97-2.82] | 49.86 | 0.06 |
| Satiroglu, 2019 | 2.01 [1.36-2.97] | 0.00 | 0.02 | 2.01 [1.36 2.97] | 0.00 | 0.02 |
| MPV as a categorized variable for in-hospital MACE with OR as an effect estimate | | | | | | |
| Osuna, 1998 | 1.88 [0.94-3.75] | 87.95 | 0.07 | 2.03 [0.95-4.36] | 92.64 | 0.06 |
| Vakili, 2009 | 1.55 [0.89-2.69] | 81.33 | 0.10 | 1.70 [0.87-3.32] | 90.58 | 0.10 |
| Akpek, 2011 | 1.84 [0.93-3.62] | 89.97 | 0.07 | 2.01 [0.94-4.29] | 94.17 | 0.07 |
| Liu, 2014 | 1.87 [0.93-3.78] | 87.02 | 0.07 | 2.03 [0.93-4.40] | 92.02 | 0.07 |
| Machado, 2018 | 1.60 [1.15-2.21] | 0.00 | 0.01 | 1.83 [1.10-3.06] | 58.59 | 0.03 |
| Garlobo, 2019 | 1.50 [1.04-2.17] | 77.68 | 0.04 | 1.53 [1.05-2.24] | 82.10 | 0.03 |
| Vogiatzis, 2019 | 1.56 [0.95-2.55] | 81.68 | 0.07 | 1.61 [0.95-2.75] | 86.71 | 0.07 |
| Chen, 2020 | 1.63 [0.87-3.04] | 85.38 | 0.10 | 1.85 [0.87-3.95] | 93.52 | 0.09 |
| Patients treated with percutaneous coronary intervention | | | | | | |
| MPV as a continuous variable for long-term mortality with HR as an effect estimate | | | | | | |
| Eisen ACS, 2013 | 1.33 [1.26 1.40] | 0.11 | 0.00 | 1.33 [1.26 1.40] | 0.03 | 0.00 |
| Eisen SCAD, 2013 | 1.30 [1.18 1.44] | 48.25 | 0.00 | 1.31 [1.18 1.45] | 59.48 | 0.00 |
| Lekston, 2014 | 1.27 [1.16 1.39] | 25.29 | 0.00 | 1.28 [1.16 1.42] | 44.55 | 0.00 |
| Wasilewski, 2016 | 1.29 [1.18 1.41] | 46.11 | 0.00 | 1.29 [1.18 1.42] | 58.16 | 0.00 |
| Yu, 2017 | 1.30 [1.18 1.42] | 48.64 | 0.00 | 1.30 [1.18 1.44] | 61.16 | 0.00 |
| Tian, 2018 | 1.28 [1.19 1.37] | 37.58 | 0.00 | 1.28 [1.19 1.37] | 47.02 | 0.00 |
| Jiang, 2019 | 1.29 [1.20 1.39] | 44.35 | 0.00 | 1.29 [1.20 1.39] | 54.46 | 0.00 |
| Patients with acute coronary syndrome | | | | | | |
| MPV as a continuous variable for long-term mortality with HR as an effect estimate | | | | | | |
| Taglieri, 2011 | 1.30 [1.17 1.43] | 51.97 | 0.00 | 1.30 [1.17 1.44] | 62.86 | 0.00 |
| Eisen ACS, 2013 | 1.33 [1.26 1.41] | 0.11 | 0.00 | 1.33 [1.26 1.41] | 0.11 | 0.00 |
| Lekston, 2014 | 1.26 [1.13 1.40] | 24.81 | 0.00 | 1.27 [1.14 1.42] | 42.72 | 0.00 |

| | | | | | | |
|---|---|---|---|---|---|---|
| Wasilewski, 2016 | 1.28 [1.16 1.42] | 50.70 | 0.00 | 1.29 [1.16 1.43] | 61.76 | 0.00 |
| Yu, 2017 | 1.29 [1.16 1.43] | 52.61 | 0.00 | 1.30 [1.16 1.44] | 63.82 | 0.00 |
| Tian, 2018 | 1.27 [1.18 1.37] | 42.12 | 0.00 | 1.27 [1.18 1.36] | 51.18 | 0.00 |
| MPV as a continuous variable for long-term MACE with HR as an effect estimate | | | | | | |
| Taglieri, 2011 | 1.22 [1.10 1.37] | 93.96 | 0.00 | 1.23 [1.10 1.37] | 94.77 | 0.00 |
| Eisen ACS, 2013 | 1.24 [1.11 1.37] | 92.30 | 0.00 | 1.24 [1.12 1.38] | 93.29 | 0.00 |
| Wasilewski, 2016 | 1.23 [1.11 1.37] | 93.79 | 0.00 | 1.24 [1.11 1.37] | 94.60 | 0.00 |
| Yu, 2017 | 1.21 [1.09 1.35] | 93.50 | 0.00 | 1.21 [1.09 1.35] | 94.44 | 0.00 |
| Tian, 2018 | 1.19 [1.08 1.32] | 91.71 | 0.00 | 1.20 [1.08 1.32] | 93.14 | 0.00 |
| Lopez-Cuenca , 2012 | 1.22 [1.09 1.35] | 93.52 | 0.00 | 1.22 [1.10 1.35] | 94.43 | 0.00 |
| Wan, 2014 | 1.23 [1.11 1.37] | 87.26 | 0.00 | 1.24 [1.11 1.38] | 88.78 | 0.00 |
| Niu, 2018 | 1.17 [1.09 1.26] | 86.25 | 0.00 | 1.17 [1.09 1.26] | 88.34 | 0.00 |
| Navarta , 2019 | 1.24 [1.13 1.37] | 84.01 | 0.00 | 1.25 [1.13 1.37] | 86.28 | 0.00 |
| Liang, 2023 | 1.22 [1.10 1.36] | 93.87 | 0.00 | 1.22 [1.10 1.36] | 94.73 | 0.00 |
| Chen Xinsen, 2020 | 1.21 [1.09 1.35] | 93.46 | 0.00 | 1.22 [1.09 1.35] | 94.39 | 0.00 |
| MPV as a categorical variable for long-term MACE with HR as an effect estimate | | | | | | |
| Taglieri, 2011 | 2.35 [1.46 3.78] | 73.09 | 0.00 | 2.39 [1.47 3.88] | 77.12 | 0.00 |
| Lopez-Cuenca , 2012 | 2.27 [1.39 3.70] | 79.39 | 0.00 | 2.32 [1.41 3.81] | 82.70 | 0.00 |
| Niu, 2018 | 2.09 [1.28 3.40] | 75.92 | 0.01 | 2.14 [1.30 3.53] | 80.36 | 0.01 |
| Navarta , 2019 | 2.06 [1.30 3.26] | 75.62 | 0.01 | 2.10 [1.32 3.36] | 80.00 | 0.01 |
| Dogan, 2012 | 2.32 [1.43 3.77] | 77.30 | 0.00 | 2.36 [1.44 3.87] | 80.77 | 0.00 |
| Fabregat-Andres, 2013 | 2.11 [1.32 3.37] | 77.40 | 0.01 | 2.15 [1.33 3.48] | 81.33 | 0.01 |
| Choi, 2014 | 2.08 [1.37 3.18] | 75.80 | 0.00 | 2.11 [1.38 3.24] | 79.56 | 0.00 |
| Navarta, 2015 | 1.79 [1.30 2.47] | 49.32 | 0.00 | 1.83 [1.31 2.54] | 57.55 | 0.00 |
| Chang, 2019 | 2.36 [1.48 3.76] | 69.35 | 0.00 | 2.41 [1.50 3.86] | 74.06 | 0.00 |

| | | | | | | |
|---|---|---|---|---|---|---|
| Nozari, 2019 | 2.29 [1.41 3.74] | 78.74 | 0.00 | 2.34 [1.42 3.84] | 82.07 | 0.00 |
| MPV as a categorized variable for in-hospital mortality with OR as an effect estimate | | | | | | |
| Tekbas, 2011 | 1.48 [0.82 2.65] | 0.00 | 0.10 | 1.51 [0.77 2.97] | 8.19 | 0.12 |
| Ranjith, 2015 | 1.74 [0.88 3.44] | 31.97 | 0.07 | 1.78 [0.84 3.75] | 58.83 | 0.08 |
| Monteiro Junior , 2018 | 1.66 [0.97 2.85] | 27.21 | 0.06 | 1.65 [0.97 2.82] | 49.86 | 0.06 |
| Satiroglu, 2019 | 2.01 [1.36 2.97] | 0.00 | 0.02 | 2.01 [1.36 2.97] | 0.00 | 0.02 |
| MPV as a categorized variable for in-hospital MACE with OR as an effect estimate | | | | | | |
| Osuna, 1998 | 1.88 [0.94 3.75] | 87.95 | 0.07 | 2.03 [0.95 4.36] | 92.6 | 0.06 |
| Vakili, 2009 | 1.55 [0.89 2.69] | 81.33 | 0.10 | 1.70 [0.87 3.32] | 90.58 | 0.10 |
| Akpek, 2011 | 1.84 [0.93 3.62] | 89.97 | 0.07 | 2.01 [0.94 4.29] | 94.17 | 0.07 |
| Liu, 2014 | 1.87 [0.93 3.78] | 87.02 | 0.07 | 2.03 [0.93 4.40] | 92.02 | 0.07 |
| Machado, 2018 | 1.60 [1.15 2.21] | 0.00 | 0.01 | 1.83 [1.10 3.06] | 0.07 | 0.03 |
| Garlobo, 2019 | 1.50 [1.04 2.17] | 77.68 | 0.04 | 1.53 [1.05 2.24] | 82.10 | 0.03 |
| Vogiatzis, 2019 | 1.56 [0.95 2.55] | 81.68 | 0.07 | 1.61 [0.95 2.75] | 86.71 | 0.07 |
| Chen, 2020 | 1.63 [0.87 3.04] | 85.38 | 0.10 | 1.85 [0.87 3.95] | 93.52 | 0.09 |
| Patients with stable coronary artery disease | | | | | | |
| MPV as a continuous variable for long-term mortality with HR as an effect estimate | | | | | | |
| Eisen SCAD, 2013 | 1.52 [0.23 10.09] | 0.01 | 0.22 | 1.53 [0.21 11.35] | 8.47 | 0.23 |
| Jiang, 2019 | 1.28 [0.92 1.77] | 0.00 | 0.07 | 1.28 [0.92 1.77] | 0.00 | 0.07 |
| Adali, 2022 | 1.28 [0.61 2.67] | 0.00 | 0.15 | 1.45 [0.09 24.3] | 52.96 | 0.34 |

Abbreviations: ML, maximum likelihood; REML, restricted  maximum likelihood; CI, confidence interval; MPV, mean platelet volume; HR, hazard ratio; MACE, major adverse cardiovascular events.

**Supplementary Table 5. Subgroup analyses.**

| **Subgroups** | **Estimate** | **P value for subgroup difference** |
|---|---|---|
| **All ischemic heart disease patients** | | |

| | | |
|---|---|---|
| **MPV as a continuous variable for long-term mortality with HR as an effect estimate** | | |
| SCAD | 1.29 [1.07-1.55] - 3 studies | 0.985 |
| ACS | 1.29 [1.19-1.39] - 9 studies | |
| **MPV as a continuous variable for long-term MACE with HR as an effect estimate** | | |
| SCAD | 0.98 [0.66-1.45] - 3 studies | 0.030 |
| ACS | 1.22 [1.11-1.34] - 11 studies | |
| **MPV as a categorical variable for long-term MACE with HR as an effect estimate** | | |
| SCAD | 0.70 [0.54-0.91] - 1 study | <.001 |
| ACS | 2.17 [1.41-3.32] - 10 study | |
| **All percutaneous coronary intervention patients** | | |
| **MPV as a continuous variable for long-term mortality with HR as an effect estimate** | | |
| SCAD | 1.28 [0.61-2.67] - 2 studies | 0.833 |
| ACS | 1.30 [1.17-1.43] - 5 studies | |
| **MPV as a continuous variable for long-term MACE with HR as an effect estimate** | | |
| SCAD | 1.00 [0.17-5.88] - 2 studies | 0.177 |
| ACS | 1.22 [1.05-1.42] - 5 studies | |
| **MPV as a categorical variable for long-term MACE with HR as an effect estimate** | | |
| SCAD | 0.70 [0.54-0.91] - 1 study | <.001 |
| ACS | 2.81 [1.03-7.67] - 4 studies | |
| Patients with acute coronary syndrome | | |
| **MPV as a continuous variable for long-term mortality with HR as an effect estimate** | | |
| STEMI | 1.35 [0.90-2.04] - 2 studies | 0.127 |
| NST-ACS | 1.27 [1.15-1.41] - 3 studies | |
| **MPV as a continuous variable for long-term MACE with HR as an effect estimate** | | |
| STEMI | 1.41 [1.00-1.97] - 2 studies | <.001 |
| NST-ACS | 1.19 [1.11-1.28] - 5 studies | |
| **MPV as a continuous variable for long-term MACE with HR as an effect estimate** | | |
| SCAD | 3.35 [1.12-10.05] - 1 study | 0.152 |
| ACS | 1.50 [1.32-1.69] - 4 studies | |

**Supplementary Table 6. Meta-regression analysis.**

| | **All IHD: HR, long-term mortality, continuous** | | | **All IHD: HR, long-term MACE, continuous** | | | **All IHD: HR, long-term MACE, categorical** | | | **ACS: HR, long-term MACE, continuous** | | | **ACS: HR, long-term MACE, categorical** | | |
|---|---|---|---|---|---|---|---|---|---|---|---|---|---|---|---|
| | N | estimate | p value | N | estimate | p value | N | estimate | p value | N | estimate | p value | N | estimate | p value |
| Analyser | 9 | -0.11, –0.01 | 0.49-0.91 | 15 | | 0.27-0.10 | 12 | | 0.39, 0.53 | 11 | | 0,37-0.99 | ***10*** | ***-0.51*** | ***0.0087 (for Advia)*** |
| Publication year | 9 | 0.02 | 0.002 | 15 | -0.0025 | 0.83 | 12 | -0.0231 | 0.7071 | 11 | 0.0088 | 0.4674 | 10 | 0.0198 | 0.693 |
| Study design | 9 | -0.49 | 0.16 | ***15*** | ***0.16*** | ***0.02*** | 12 | 0.3418 | 0.3552 | 11 | 0.1334 | 0.0717 | 10 | 0.0007 | 0.9983 |
| Sample | 9 | -0.00 | 0.003 | 15 | -0.0000 | 0.83 | 12 | -0.0002 | 0.0831 | 11 | 0.0000 | 0.8063 | 10 | -0.0001 | 0.4456 |
| ACS | 9 | -0.0001 | 0.87 | ***15*** | ***0.0022*** | ***0.0079*** | 12 | 0.0096 | 0.0567 | - | - | - | - | - | - |
| STEMI | 8 | 0.0005 | 0.16 | ***12*** | ***0.0028*** | ***0.02*** | 10 | 0.0121 | 0.0549 | 9 | 0.0020 | 0.0923 | 9 | 0.0092 | 0.0973 |
| NST-ACS | 8 | -0.0005 | 0.37 | 12 | 0.0008 | 0.49 | 10 | 0.0029 | 0.5719 | 9 | -0.0020 | 0.0924 | 9 | -0.0092 | 0.0973 |
| SCAD | 9 | 0.0001 | 0.87 | ***15*** | ***-0.0022*** | ***0.0079*** | 12 | -0.0096 | 0.0567 | - | - | - | - | - | - |
| PCI | 8 | 0.0015 | 0.58 | 9 | -0.0014 | 0.75 | 8 | -0.0085 | 0.5105 | 6 | 0.0007 | 0.8410 | 6 | -0.0051 | 0.691 |
| Age | 9 | -0.01 | 0.03 | 15 | -0.0121 | 0.11 | 12 | 0.0081 | 0.8161 | 11 | -0.0110 | 0.0753 | 10 | 0.0087 | 0.7568 |
| Males | 9 | -0.0076 | 0.05 | 15 | -0.001 | 0.84 | ***12*** | ***-0.0553*** | ***0.010*** | 11 | 0.0089 | 0.0790 | 10 | -0.0352 | 0.1199 |

| | | | | | | | | | | | | | | | |
|---|---|---|---|---|---|---|---|---|---|---|---|---|---|---|---|
| hypertension | 8 | -0.0048 | 0.06 | ***14*** | ***-0.006*** | ***0.0161*** | 12 | -0.005 | 0.7575 | 11 | -0.0047 | 0.0702 | 10 | 0.0055 | 0.6875 |
| diabetes | 8 | 0.0012 | 0.71 | 15 | -0.005 | 0.2624 | *12* | *-0.0392* | *0.0143* | 11 | -0.0002 | 0.9672 | 10 | -0.0253 | 0.0763 |
| dyslipidemia | 6 | -0.0002 | 0.92 | 11 | -0.0027 | 0.4994 | 12 | -0.022 | 0.0649 | 8 | 0.0072 | 0.0623 | 10 | -0.0054 | 0.7408 |
| smoking | 8 | 0.001 | 0.72 | ***15*** | ***0.0078*** | ***0.0002*** | 12 | 0.0106 | 0.4510 | ***11*** | ***0.0068*** | ***0.0017*** | 10 | -0.0038 | 0.7738 |
| prior MI | 7 | -0.0017 | 0.48 | 9 | -0.005 | 0.0213 | 5 | -0.0281 | 0.0573 | 7 | -0.0050 | 0.2637 | 5 | -0.0281 | 0.0573 |
| creatinine | 7 | 0.29 | 0.11 | 11 | -0.1824 | 0.6245 | 11 | -0.9910 | 0.4910 | 9 | -0.2979 | 0.4387 | 10 | -1.7902 | 0.142 |
| WBC | 6 | 0.006 | 0.67 | 10 | 0.0946 | 0.0577 | 8 | 0.1632 | 0.3182 | 8 | -0.0461 | 0.5801 | 7 | -0.1669 | 0.3801 |
| platelet | 6 | -0.003 | 0.003 | 13 | -0.0036 | 0.1328 | 10 | -0.0025 | 0.7711 | 9 | -0.0037 | 0.0782 | 9 | -0.0078 | 0.2122 |
| BMI | 3 | 0.0005 | 0.99 | 6 | -0.0315 | 0.5519 | 5 | -0.0503 | 0.7893 | 3 | -0.0887 | 0.0005 | 3 | -0.0873 | 0.4793 |
| SBP | 6 | -0.006 | 0.25 | 6 | -0.0047 | 0.3554 | 4 | 0.0134 | 0.7199 | 6 | -0.0047 | 0.3554 | 3 | 0.0168 | 0.6771 |
| Glucosae | 3 | -0.0004 | 0.87 | 7 | 0.0021 | 0.553 | 6 | 0.0559 | 0.0446 | 6 | 0.0005 | 0.8719 | 4 | 0.0365 | 0.0559 |
| hemoglobine | 8 | 0.005 | 0.11 | 12 | 0.0029 | 0.1158 | 9 | 0.0503 | 0.0116 | 8 | 0.0012 | 0.5558 | 8 | 0.0368 | 0.0024 |
| EF | 6 | 0.002 | 0.67 | 7 | -0.008 | 0.5613 | 7 | -0.0300 | 0.6268 | 6 | 0.0058 | 0.5963 | 5 | 0.1295 | 0.0512 |
| MVD | 7 | -0.002 | 0.02 | 5 | 0.0029 | 0.5911 | 3 | -0.0472 | 0.0191 | 3 | -0.0037 | 0.2075 | - | - | - |

**Supplementary Figures.**

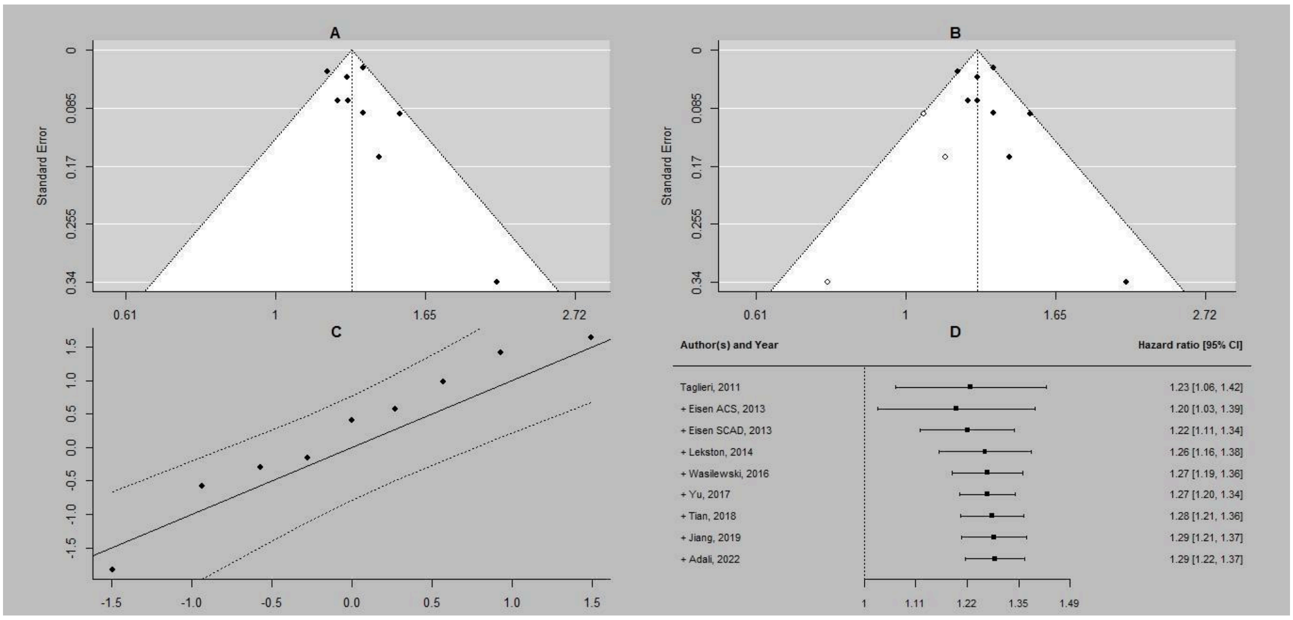

**Supplementary Figure S1. All ischemic heart disease patients. Additional analyses for long-term mortality with MPV as a continuous variable and hazard ratios as effect estimates. A, funnel plot for publication bias assessment; B, a trim and fill method for publication bias assessment; C, a Q-Q plot for ML estimation; D, cumulative meta-analysis for ML estimation.** Abbreviations: CI, confidence interval; ML, maximum likelihood; MPV, mean platelet volume.

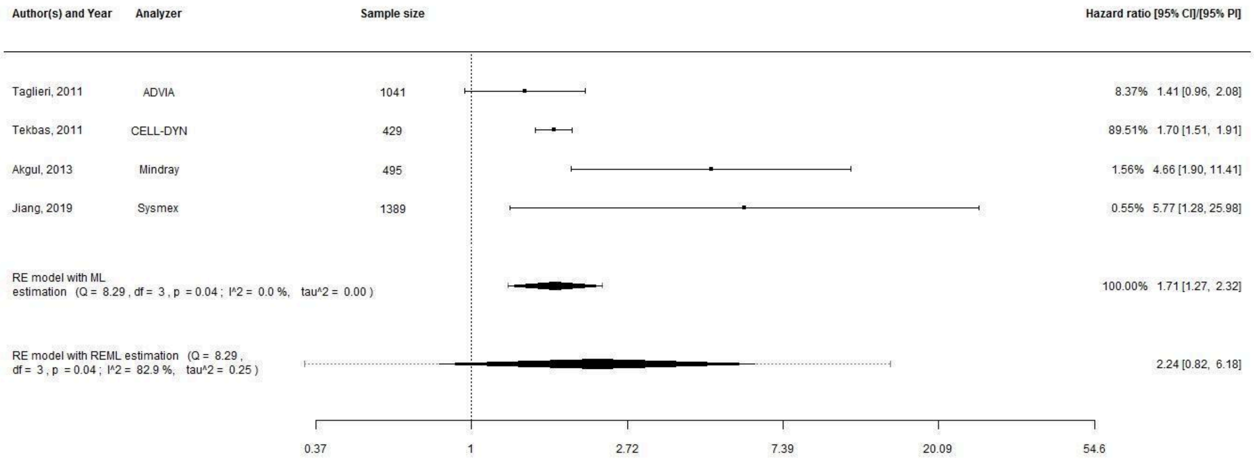

**Supplementary Figure S2. All ischemic heart disease patients. Analyses for long-term mortality with MPV treated as a categorical variable and hazard ratios as effect estimates.** Abbreviations: CI, confidence interval; PI, prediction interval; RE, random effects; ML, maximum likelihood; REML, restricted maximum likelihood; MPV, mean platelet volume.

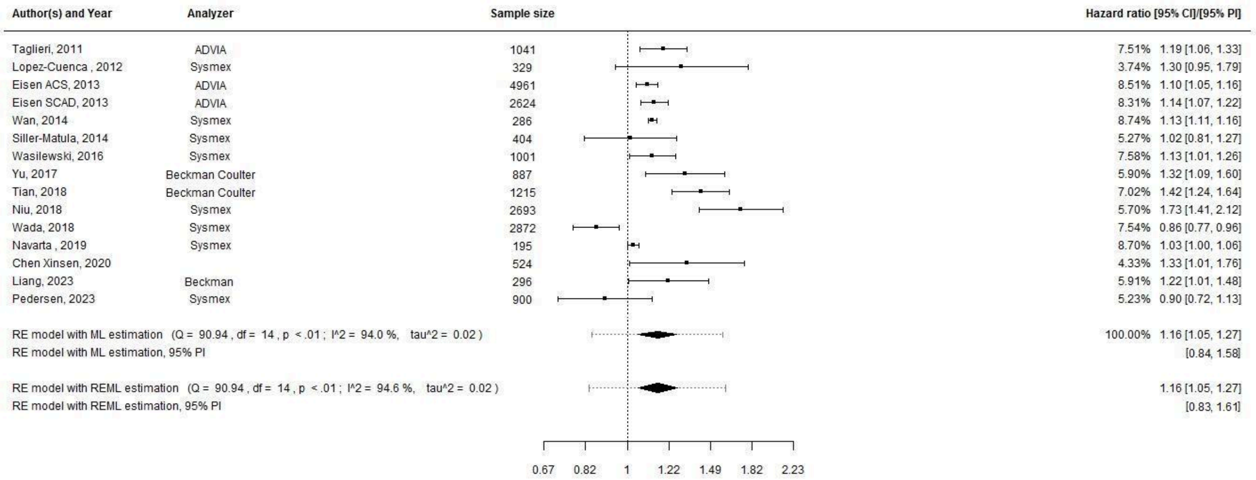

**Supplementary Figure S3. All ischemic heart disease patients. Analyses for long-term major cardiovascular events with MPV treated as a continuous variable and hazard ratios as effect estimates. Abbreviations: CI, confidence interval; PI, prediction interval; RE, random effects; ML, maximum likelihood; REML, restricted maximum likelihood; MPV, mean platelet volume.**

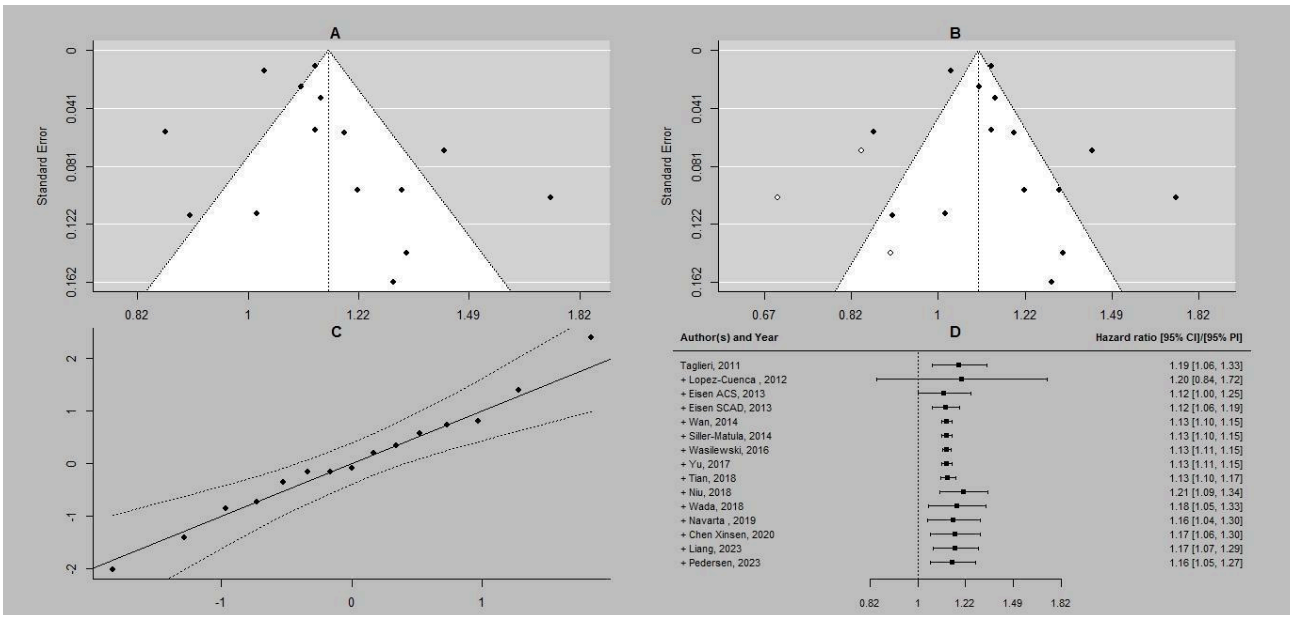

**Supplementary Figure S4. All ischemic heart disease patients. Additional analyses for long-term major cardiovascular events with MPV treated as a continuous variable and hazard ratios as effect estimates. A, funnel plot for publication bias assessment; B, a trim and fill method for publication bias assessment; C, a Q-Q plot for ML estimation; D, cumulative meta-analysis for ML estimation.**

Abbreviations: CI, confidence interval; ML, maximum likelihood; MPV, mean platelet volume.

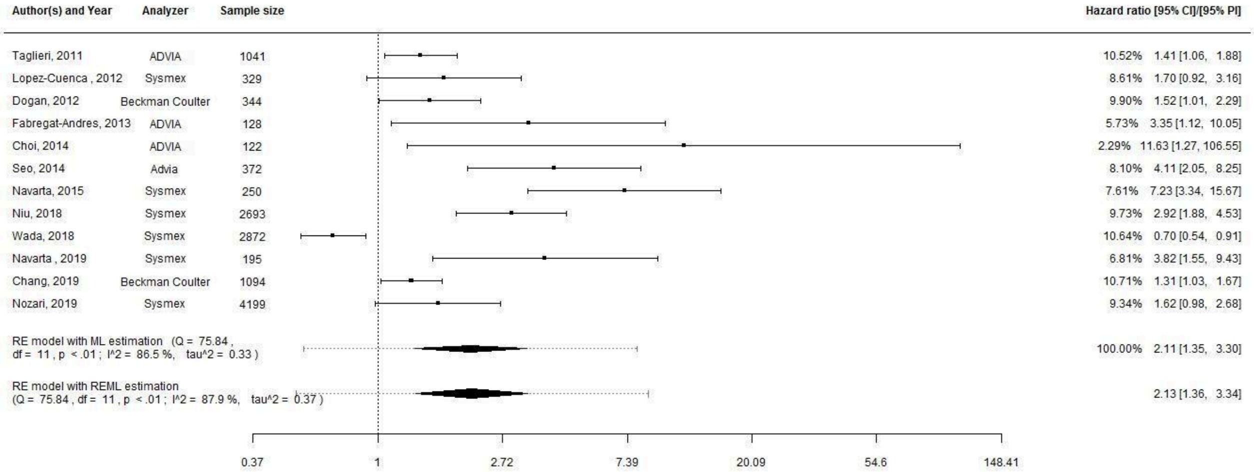

**Supplementary Figure S5. All ischemic heart disease patients. Analyses for long-term major cardiovascular events with MPV treated as a categorical variable and hazard ratios as effect estimates.**
Abbreviations: CI, confidence interval; PI, prediction interval; RE, random effects; ML, maximum likelihood; REML, restricted maximum likelihood; MPV, mean platelet volume.

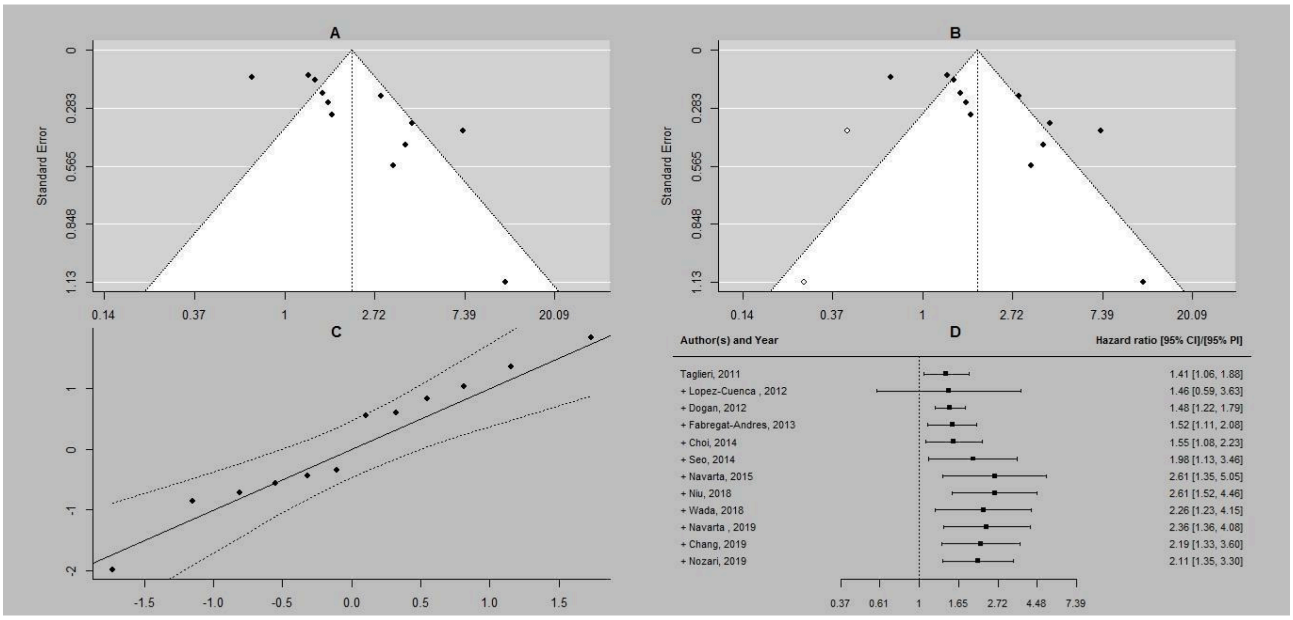

**Supplementary Figure S6. All ischemic heart disease patients. Additional analyses for long-term major cardiovascular events with MPV treated as a categorical variable and hazard ratios as effect estimates. A, funnel plot for publication bias assessment; B, a trim and fill method for publication bias assessment; C, a Q-Q plot for ML estimation; D, cumulative meta-analysis for ML estimation.**
Abbreviations: CI, confidence interval; ML, maximum likelihood; MPV, mean platelet volume.

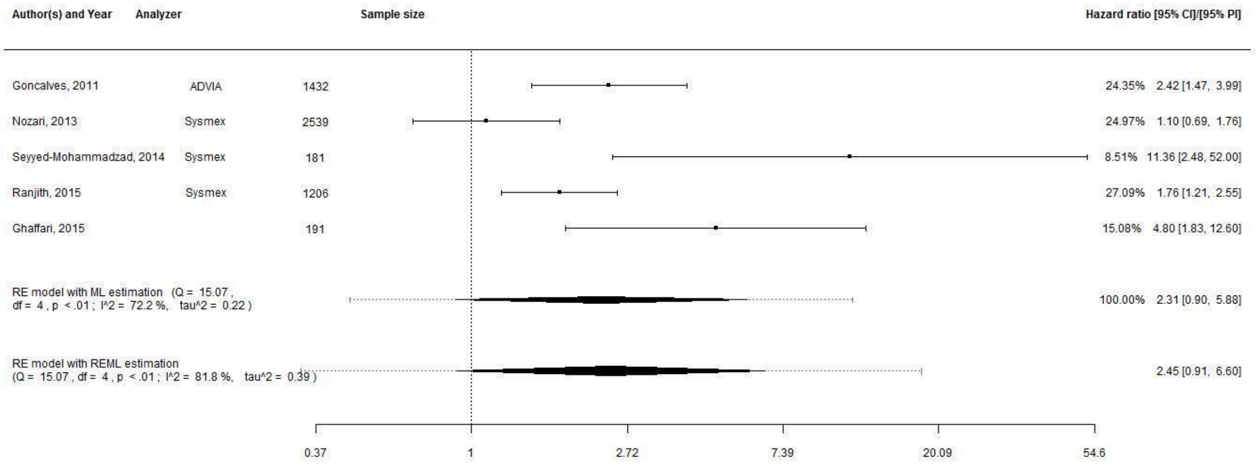

**Supplementary Figure S7. All ischemic heart disease patients. Analyses for long-term major cardiovascular events with MPV treated as a categorical variable and odds ratios as effect estimates.**
Abbreviations: CI, confidence interval; PI, prediction interval; RE, random effects; ML, maximum likelihood; REML, restricted maximum likelihood; MPV, mean platelet volume.

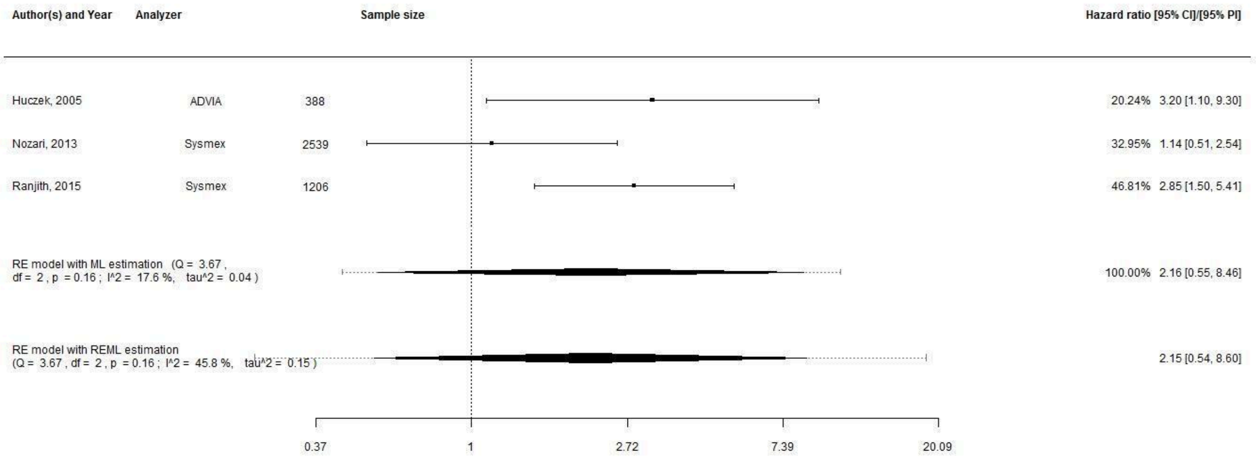

**Supplementary Figure S8. All ischemic heart disease patients. Analyses for long-term mortality with MPV treated as a categorical variable and odds ratios as effect estimates.**
Abbreviations: CI, confidence interval; PI, prediction interval; RE, random effects; ML, maximum likelihood; REML, restricted maximum likelihood; MPV, mean platelet volume.

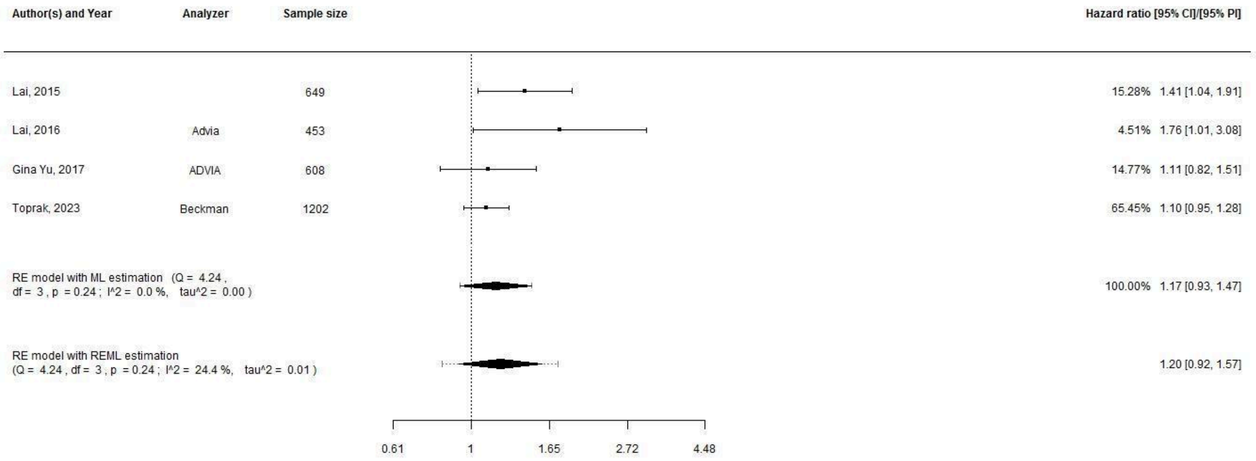

**Supplementary Figure S9. All ischemic heart disease patients. Analyses for one-month mortality with MPV treated as a continuous variable and hazard ratios as effect estimates.** Abbreviations: CI, confidence interval; PI, prediction interval; RE, random effects; ML, maximum likelihood; REML, restricted maximum likelihood; MPV, mean platelet volume.

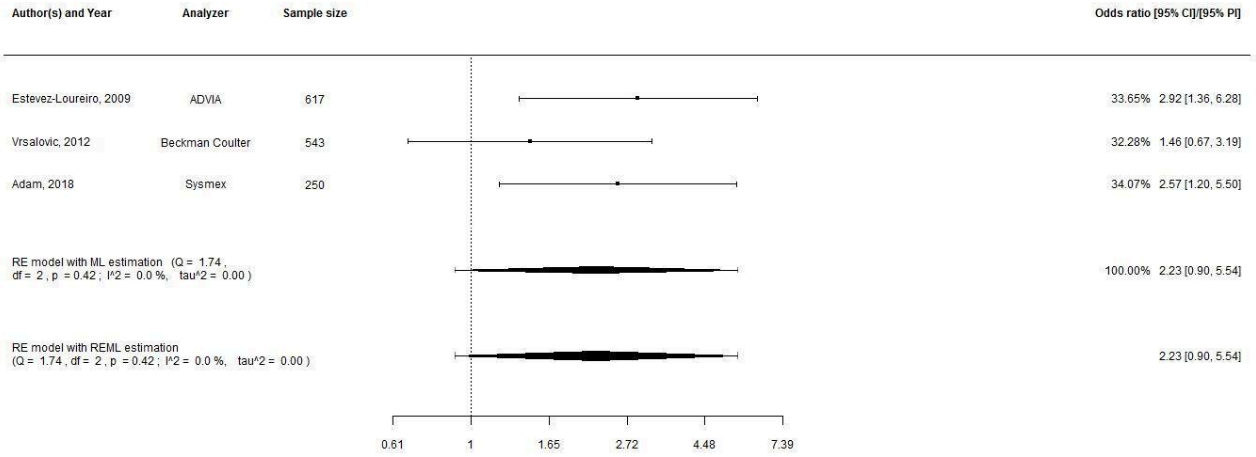

**Supplementary Figure S10. All ischemic heart disease patients. Analyses for one-month mortality with MPV treated as a categorical variable and odds ratios as effect estimates.** Abbreviations: CI, confidence interval; PI, prediction interval; RE, random effects; ML, maximum likelihood; REML, restricted maximum likelihood; MPV, mean platelet volume.

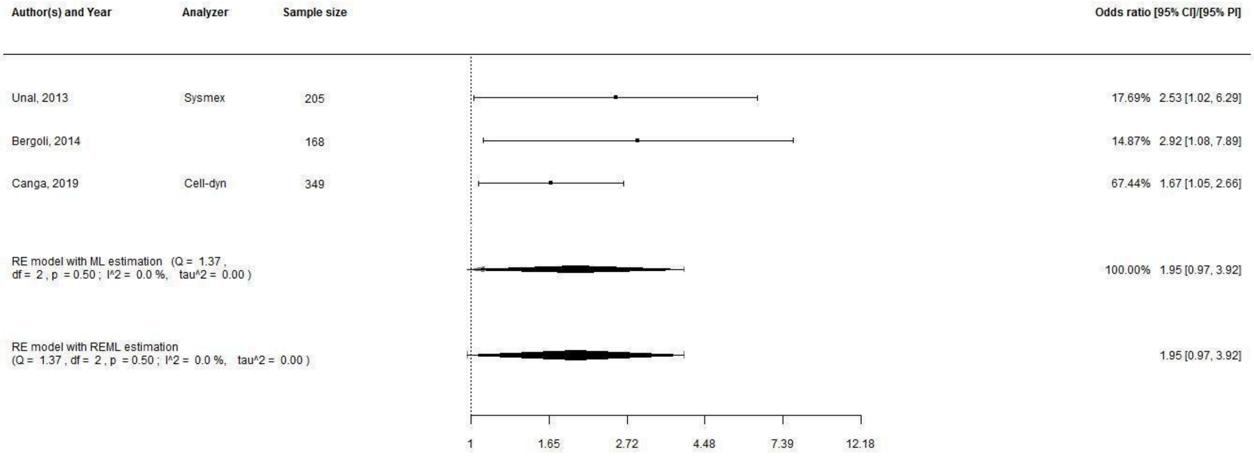

**Supplementary Figure S11. All ischemic heart disease patients. Analyses for one-month major adverse cardiovascular events with MPV treated as a categorical variable and odds ratios as effect estimates.**
Abbreviations: CI, confidence interval; PI, prediction interval; RE, random effects; ML, maximum likelihood; REML, restricted maximum likelihood; MPV, mean platelet volume.

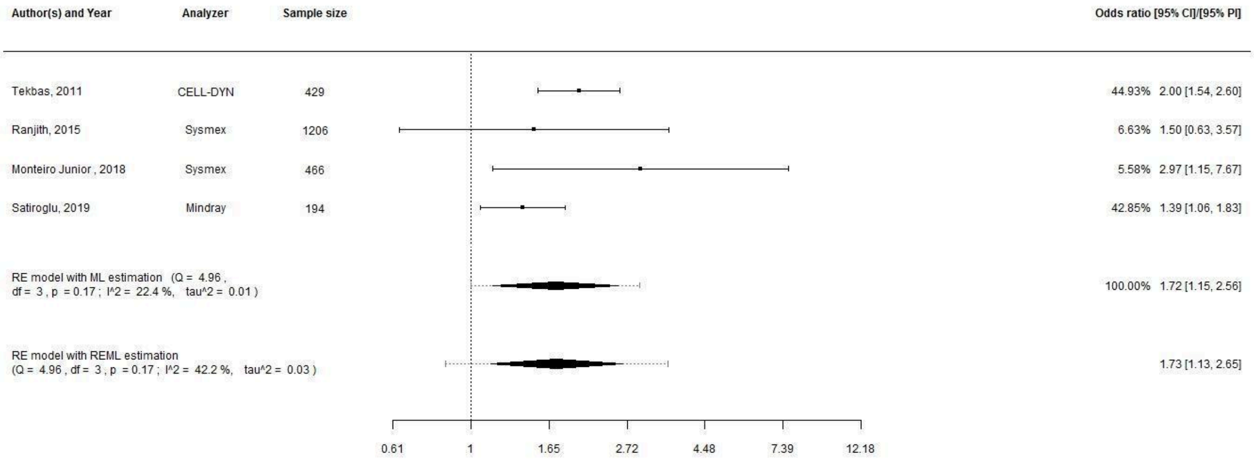

**Supplementary Figure S12. All ischemic heart disease patients. Analyses for inhospital mortality with MPV treated as a categorical variable and odds ratios as effect estimates.**
Abbreviations: CI, confidence interval; PI, prediction interval; RE, random effects; ML, maximum likelihood; REML, restricted maximum likelihood; MPV, mean platelet volume.

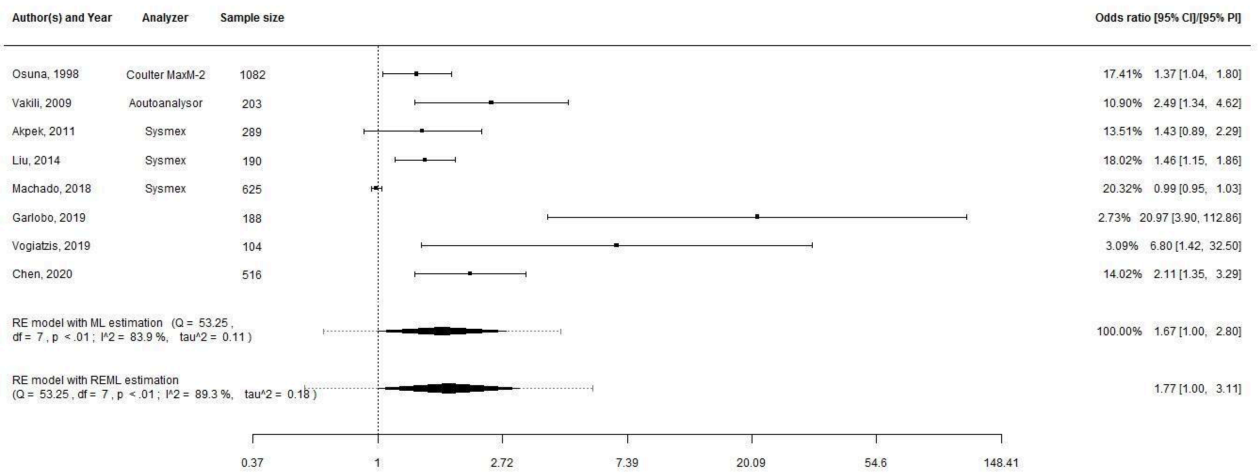

**Supplementary Figure S13. All ischemic heart disease patients. Analyses for inhospital major adverse cardiovascular events with MPV treated as a categorical variable and odds ratios as effect estimates.**

Abbreviations: CI, confidence interval; PI, prediction interval; RE, random effects; ML, maximum likelihood; REML, restricted maximum likelihood; MPV, mean platelet volume.

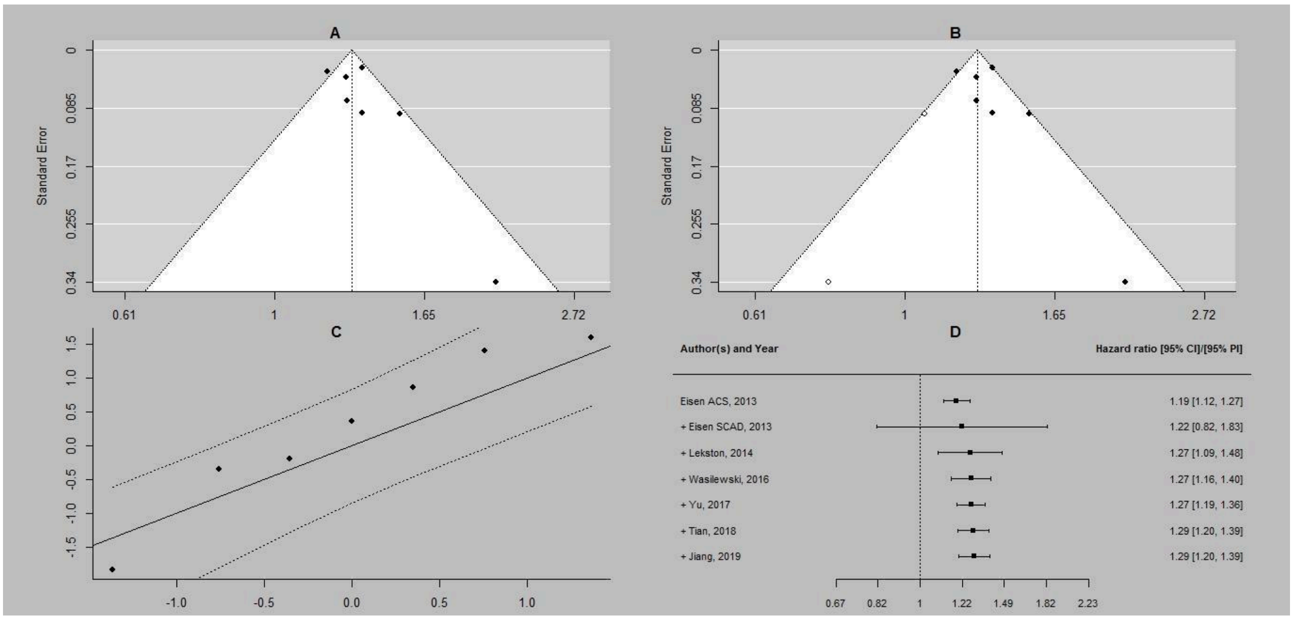

**Supplementary Figure S14. Patients treated with percutaneous coronary intervention. Additional analyses for long-term mortality with MPV treated as a continuous variable and hazard ratios as effect estimates. A, funnel plot for publication bias assessment; B, a trim and fill method for publication bias assessment; C, a Q-Q plot for ML estimation; D, cumulative meta-analysis for ML estimation.**

Abbreviations: CI, confidence interval; ML, maximum likelihood; MPV, mean platelet volume.

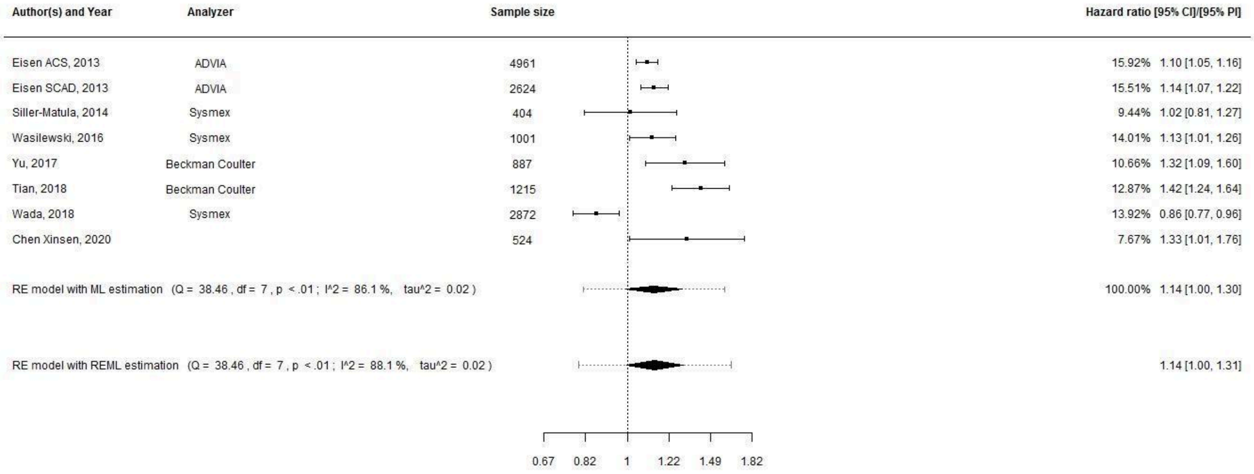

**Supplementary Figure S15. Patients treated with percutaneous coronary intervention. Analyses for long-term major adverse cardiovascular events with MPV treated as a continuous variable and hazard ratios as effect estimates.**
Abbreviations: CI, confidence interval; PI, prediction interval; RE, random effects; ML, maximum likelihood; REML, restricted maximum likelihood; MPV, mean platelet volume.

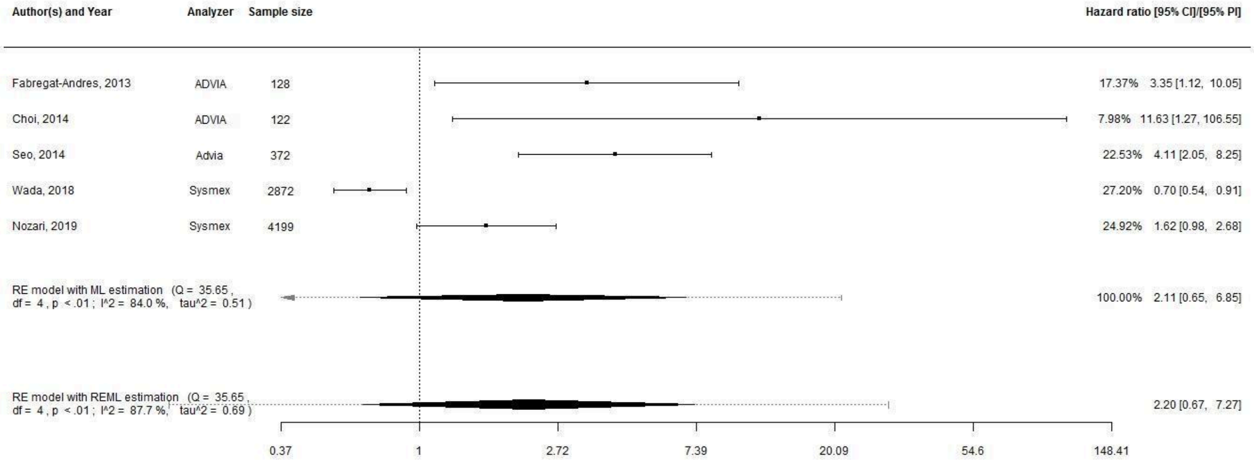

**Supplementary Figure S16. Patients treated with percutaneous coronary intervention. Analyses for long-term major adverse cardiovascular events with MPV treated as a categorical variable and hazard ratios as effect estimates.**
Abbreviations: CI, confidence interval; PI, prediction interval; RE, random effects; ML, maximum likelihood; REML, restricted maximum likelihood; MPV, mean platelet volume.

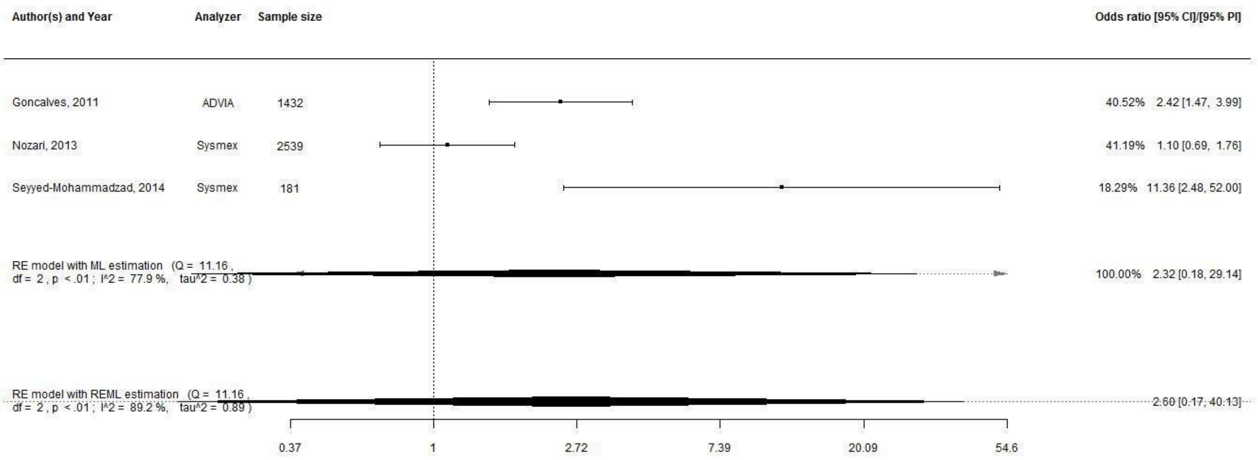

**Supplementary Figure S17. Patients treated with percutaneous coronary intervention. Analyses for long-term major adverse cardiovascular events with MPV treated as a categorical variable and odds ratios as effect estimates.**
Abbreviations: CI, confidence interval; PI, prediction interval; RE, random effects; ML, maximum likelihood; REML, restricted maximum likelihood; MPV, mean platelet volume.

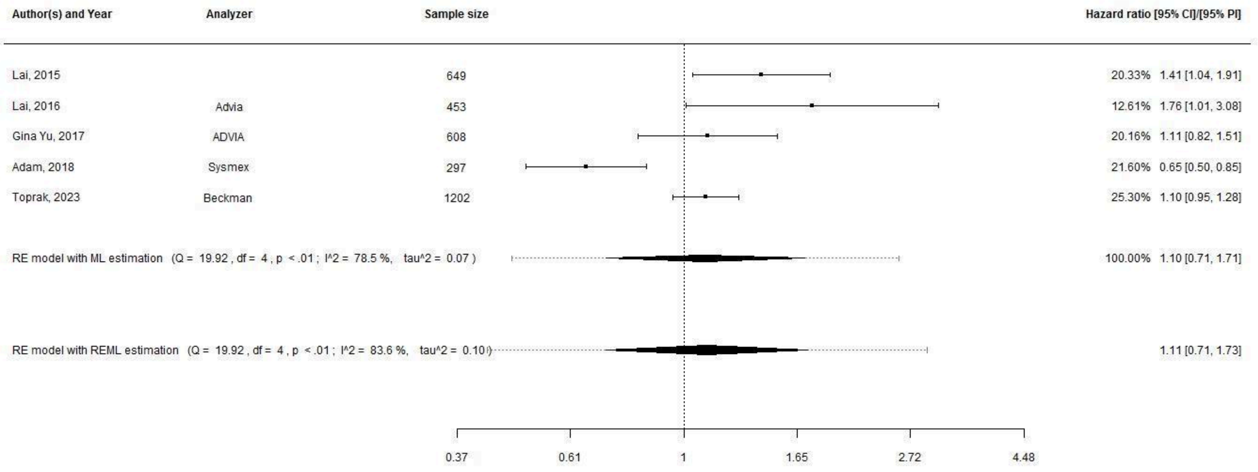

**Supplementary Figure S18. Patients treated with percutaneous coronary intervention. Analyses for one-month mortality with MPV treated as a continuous variable and hazard ratios as effect estimates.**
Abbreviations: CI, confidence interval; PI, prediction interval; RE, random effects; ML, maximum likelihood; REML, restricted maximum likelihood; MPV, mean platelet volume.

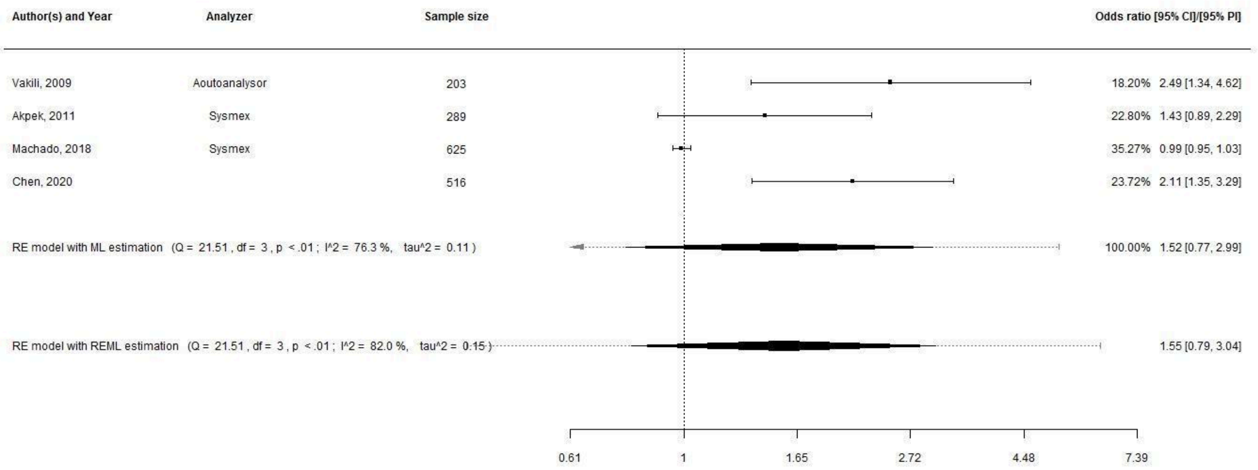

**Supplementary Figure S19. Patients treated with percutaneous coronary intervention. Analyses for inhospital major adverse cardiovascular events with MPV treated as a categorical variable and odds ratios as effect estimates.**

Abbreviations: CI, confidence interval; PI, prediction interval; RE, random effects; ML, maximum likelihood; REML, restricted maximum likelihood; MPV, mean platelet volume.

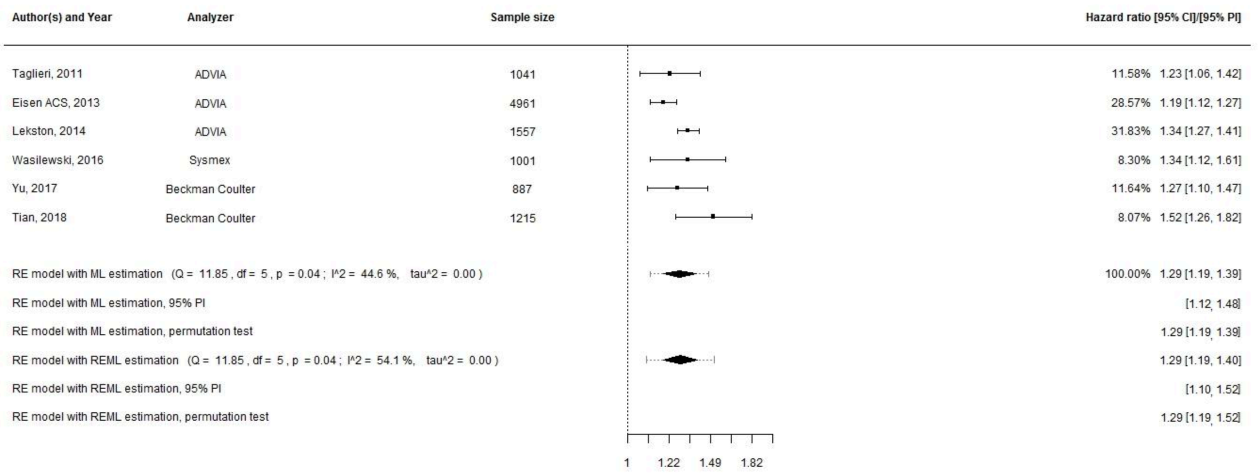

**Supplementary Figure S20. Patients with acute coronary syndrome. Analyses for long-term mortality with MPV treated as a continuous variable and hazard ratios as effect estimates.**

Abbreviations: CI, confidence interval; PI, prediction interval; RE, random effects; ML, maximum likelihood; REML, restricted maximum likelihood; MPV, mean platelet volume.

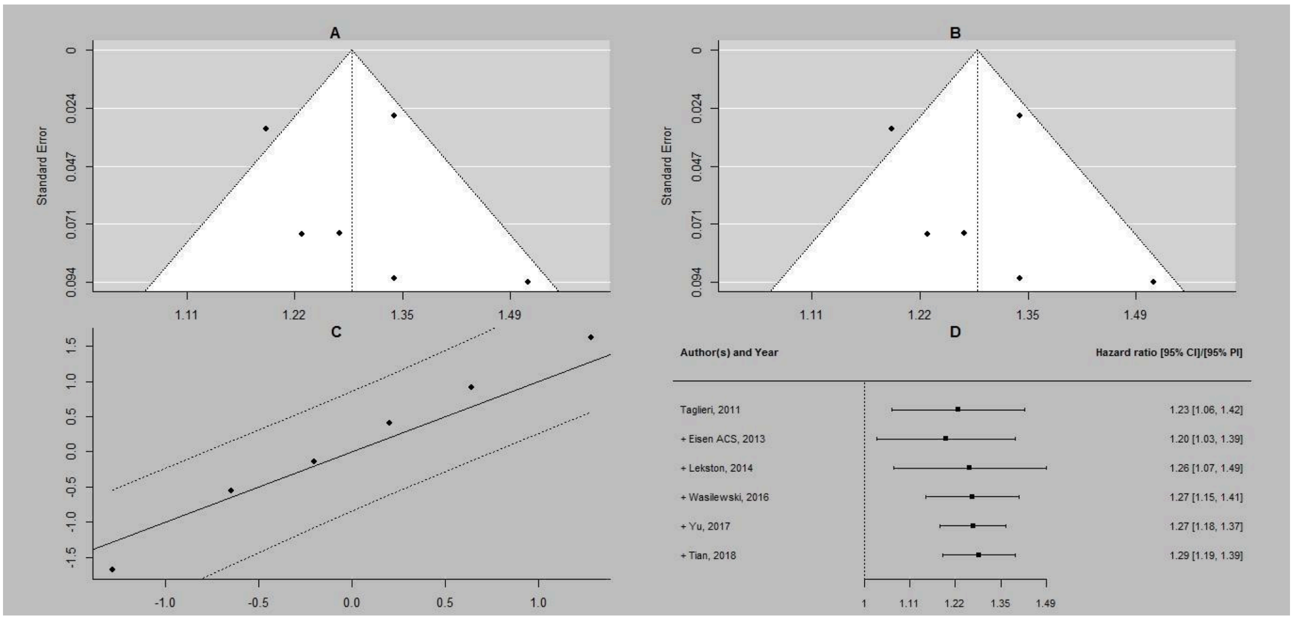

**Supplementary Figure S21. Patients with acute coronary syndrome. Additional analyses for long-term mortality with MPV treated as a continuous variable and hazard ratios as effect estimates. A, funnel plot for publication bias assessment; B, a trim and fill method for publication bias assessment; C, a Q-Q plot for ML estimation; D, cumulative meta-analysis for ML estimation.**

Abbreviations: CI, confidence interval; ML, maximum likelihood; MPV, mean platelet volume.

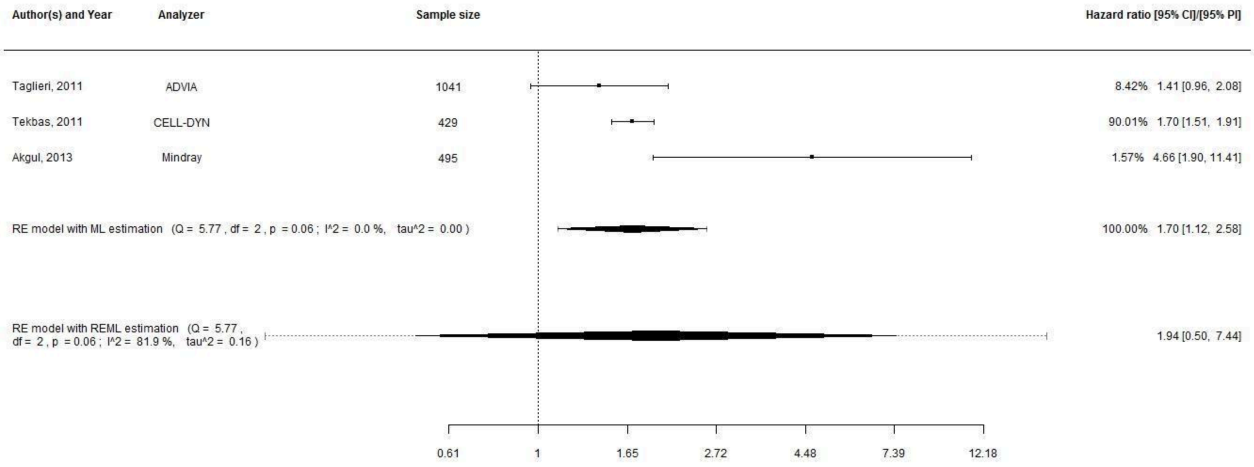

**Supplementary Figure S22. Patients with acute coronary syndrome. Analyses for long-term mortality with MPV treated as a categorical variable and hazard ratios as effect estimates.**

Abbreviations: CI, confidence interval; PI, prediction interval; RE, random effects; ML, maximum likelihood; REML, restricted maximum likelihood; MPV, mean platelet volume.

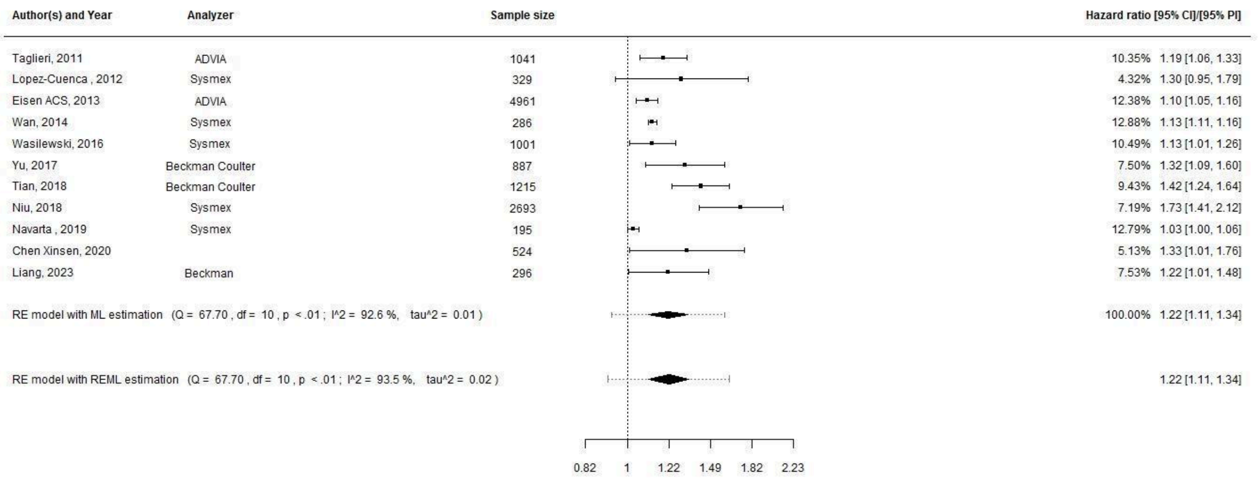

**Supplementary Figure S23. Patients with acute coronary syndrome. Analyses for long-term major adverse cardiovascular events with MPV treated as a continuous variable and hazard ratios as effect estimates. Abbreviations: CI, confidence interval; PI, prediction interval; RE, random effects; ML, maximum likelihood; REML, restricted maximum likelihood; MPV, mean platelet volume.**

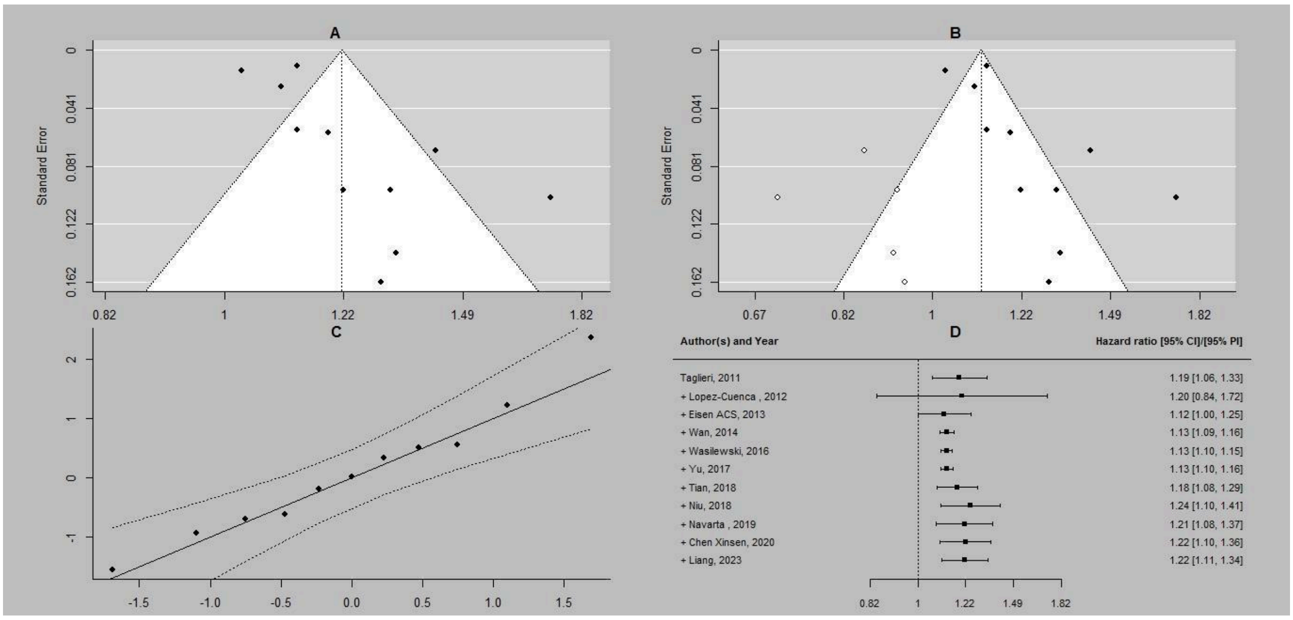

**Supplementary Figure S24. Patients with acute coronary syndrome. Additional analyses for long-term major adverse cardiovascular events with MPV treated as a continuous variable and hazard ratios as effect estimates. A, funnel plot for publication bias assessment; B, a trim and fill method for publication bias assessment; C, a Q-Q plot for ML estimation; D, cumulative meta-analysis for ML estimation. Abbreviations: CI, confidence interval; ML, maximum likelihood; MPV, mean platelet volume.**

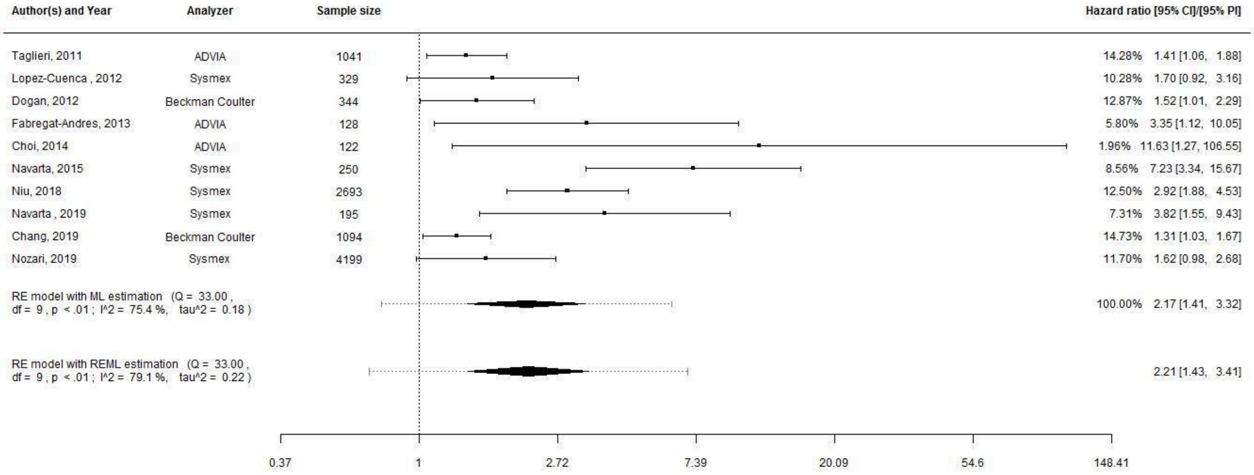

**Supplementary Figure S25. Patients with acute coronary syndrome. Analyses for long-term major adverse cardiovascular events with MPV treated as a categorical variable and hazard ratios as effect estimates. Abbreviations: CI, confidence interval; PI, prediction interval; RE, random effects; ML, maximum likelihood; REML, restricted maximum likelihood; MPV, mean platelet volume.**

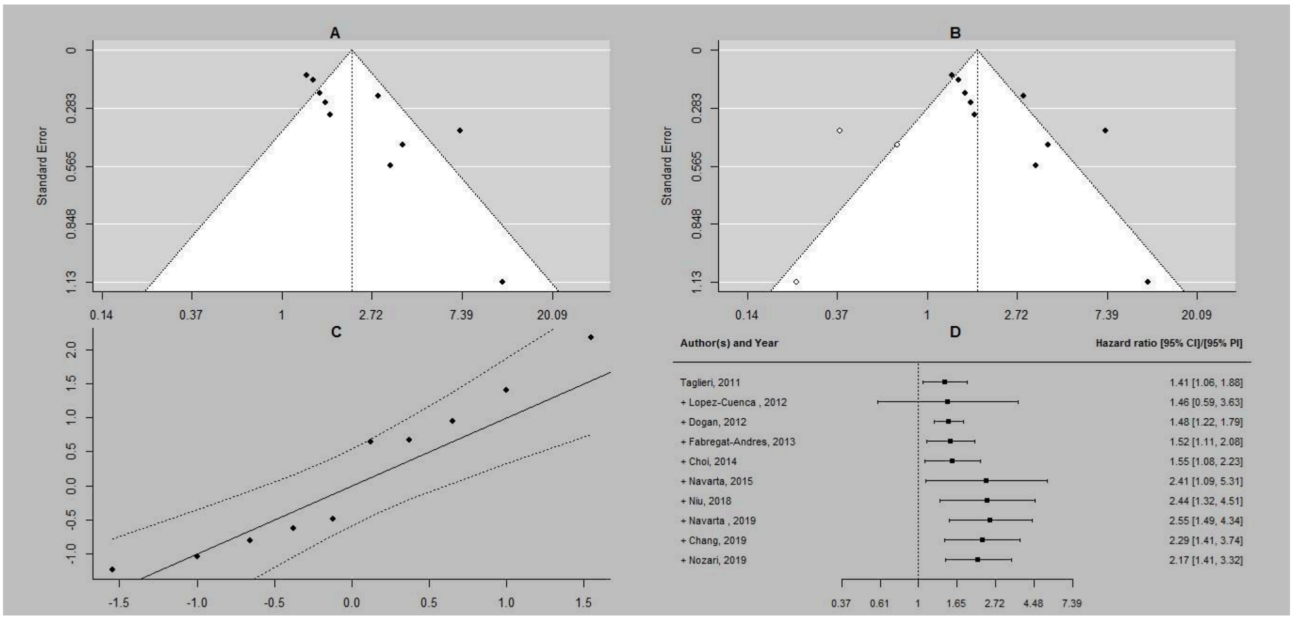

**Supplementary Figure S26. Patients with acute coronary syndrome. Additional analyses for long-term major adverse cardiovascular events with MPV treated as a categorical variable and hazard ratios as effect estimates. A, funnel plot for publication bias assessment; B, a trim and fill method for publication bias assessment; C, a Q-Q plot for ML estimation; D, cumulative meta-analysis for ML estimation. Abbreviations: CI, confidence interval; ML, maximum likelihood; MPV, mean platelet volume.**

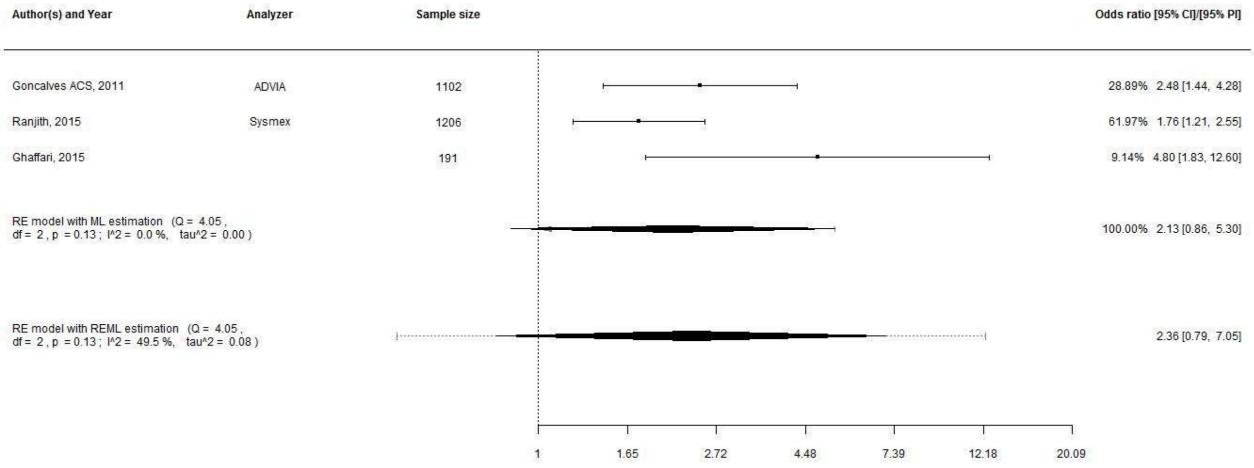

**Supplementary Figure S27. Patients with acute coronary syndrome. Analyses for long-term major adverse cardiovascular events with MPV treated as a categorical variable and odds ratios as effect estimates. Abbreviations: CI, confidence interval; PI, prediction interval; RE, random effects; ML, maximum likelihood; REML, restricted maximum likelihood; MPV, mean platelet volume.**

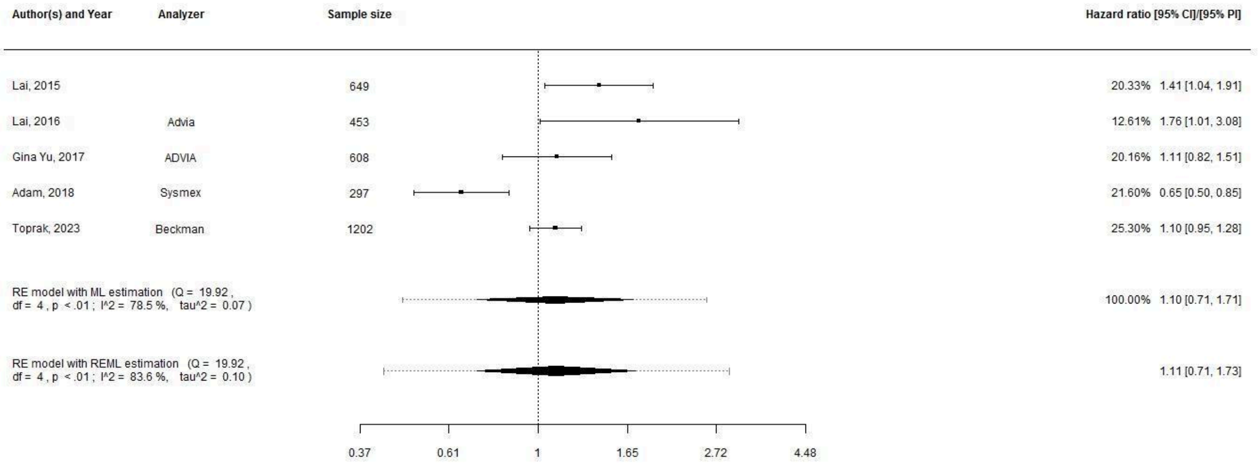

**Supplementary Figure S28. Patients with acute coronary syndrome. Analyses for one-month mortality with MPV treated as a continuous variable and hazard ratios as effect estimates. Abbreviations: CI, confidence interval; PI, prediction interval; RE, random effects; ML, maximum likelihood; REML, restricted maximum likelihood; MPV, mean platelet volume.**

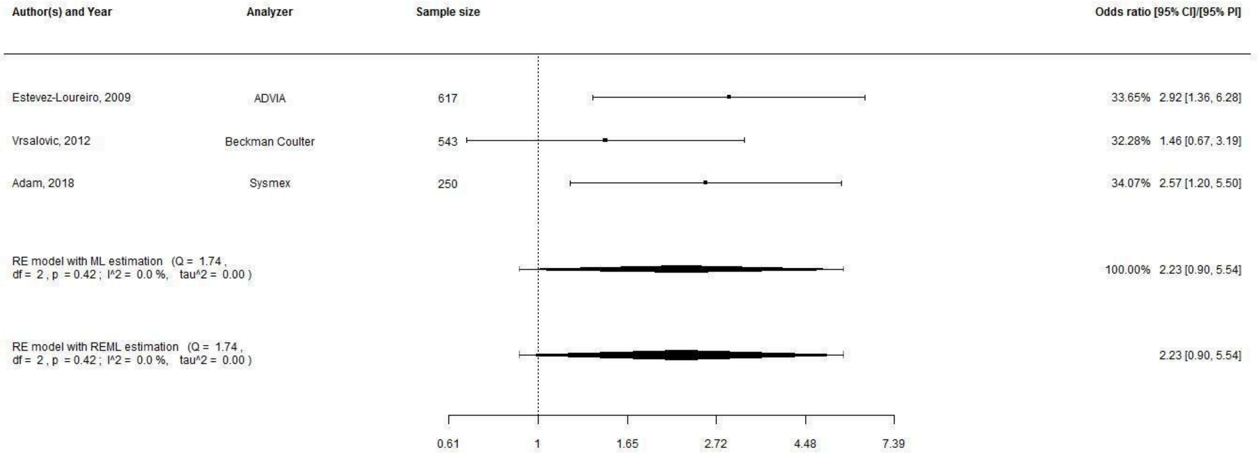

**Supplementary Figure S29. Patients with acute coronary syndrome. Analyses for one-month mortality with MPV treated as a categorical variable and odds ratios as effect estimates. Abbreviations: CI, confidence interval; PI, prediction interval; RE, random effects; ML, maximum likelihood; REML, restricted maximum likelihood; MPV, mean platelet volume.**

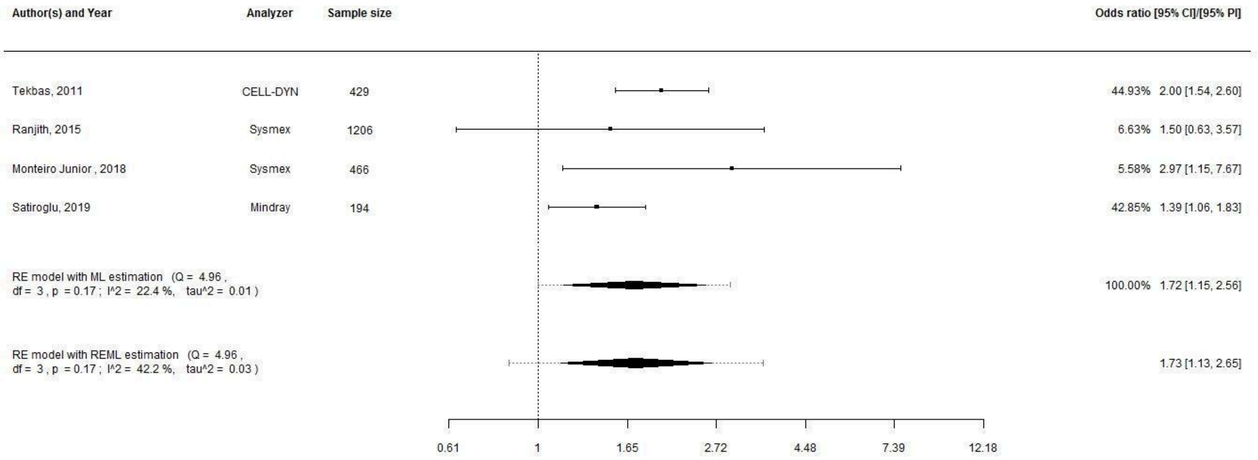

**Supplementary Figure S30. Patients with acute coronary syndrome. Analyses for inhospital mortality with MPV treated as a categorical variable and odds ratios as effect estimates. Abbreviations: CI, confidence interval; PI, prediction interval; RE, random effects; ML, maximum likelihood; REML, restricted maximum likelihood; MPV, mean platelet volume.**

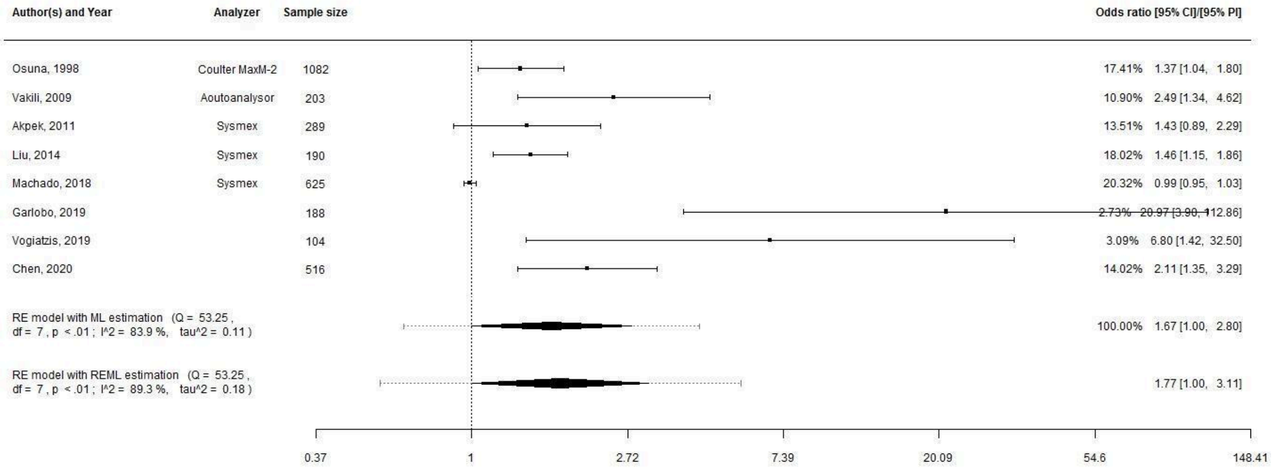

**Supplementary Figure S31. Patients with acute coronary syndrome. Analyses for inhospital major adverse cardiovascular events with MPV treated as a categorical variable and odds ratios as effect estimates. Abbreviations: CI, confidence interval; PI, prediction interval; RE, random effects; ML, maximum likelihood; REML, restricted maximum likelihood; MPV, mean platelet volume.**

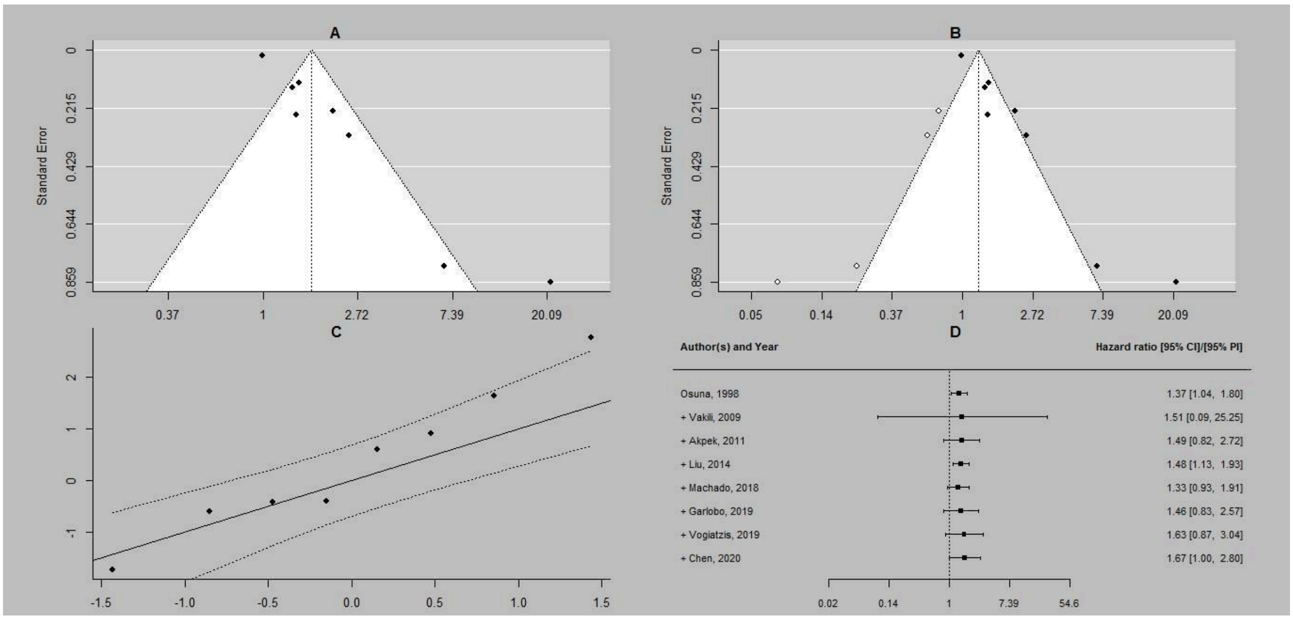

**Supplementary Figure S32. Patients with acute coronary syndrome. Additional analyses for inhospital major adverse cardiovascular events with MPV treated as a categorical variable and odds ratios as effect estimates. A, funnel plot for publication bias assessment; B, a trim and fill method for publication bias assessment; C, a Q-Q plot for ML estimation; D, cumulative meta-analysis for ML estimation. Abbreviations: CI, confidence interval; ML, maximum likelihood; MPV, mean platelet volume.**

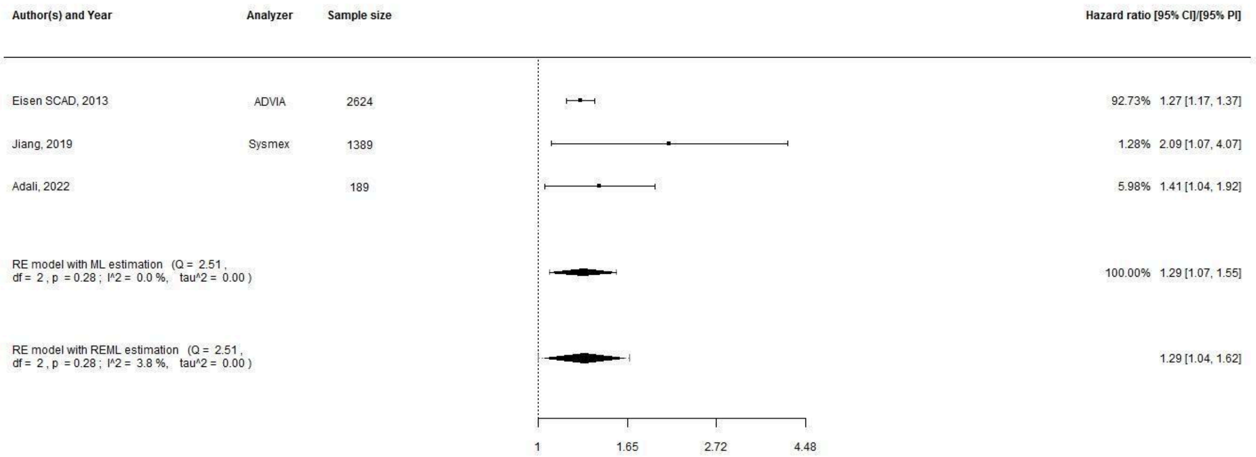

**Supplementary Figure S33. Patients with stable coronary artery disease. Analyses for long-term mortality with MPV treated as a continuous variable and hazard ratios as effect estimates. Abbreviations: CI, confidence interval; PI, prediction interval; RE, random effects; ML, maximum likelihood; REML, restricted maximum likelihood; MPV, mean platelet volume.**

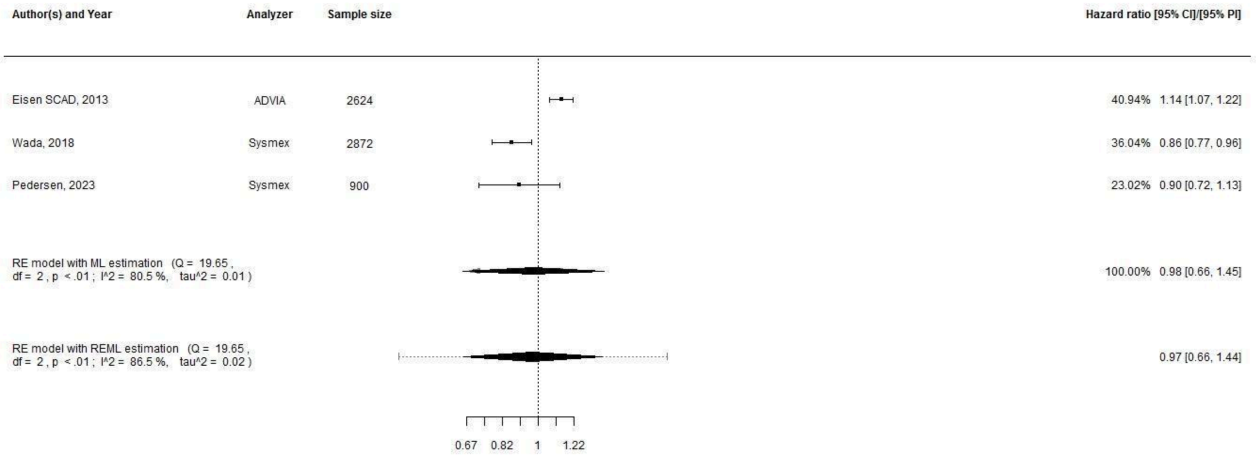

**Supplementary Figure S34. Patients with stable coronary artery disease. Analyses for long-term major adverse cardiovascular events with MPV treated as a continuous variable and hazard ratios as effect estimates. Abbreviations: CI, confidence interval; PI, prediction interval; RE, random effects; ML, maximum likelihood; REML, restricted maximum likelihood; MPV, mean platelet volume.**

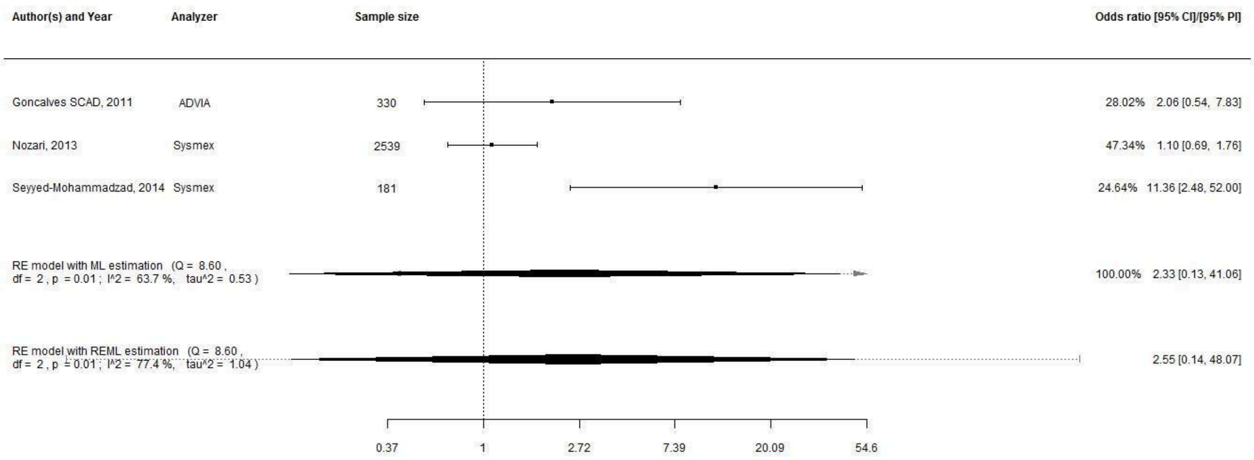

**Supplementary Figure S35. Patients with stable coronary artery disease. Analyses for long-term major adverse cardiovascular events with MPV treated as a categorical variable and odds ratios as effect estimates. Abbreviations: CI, confidence interval; PI, prediction interval; RE, random effects; ML, maximum likelihood; REML, restricted maximum likelihood; MPV, mean platelet volume.**
